# A rapid review of physical health risks associated with special procedures (tattooing, body piercing, acupuncture, electrolysis)

**DOI:** 10.1101/2023.12.11.23299649

**Authors:** Meg Kiseleva, Judit Csontos, Deborah Edwards, Elizabeth Gillen, Mala Mann, Lydia Searchfield, Abubakar Sha’aban, Ruth Lewis, Alison Cooper, Adrian Edwards

**Affiliations:** Specialist Unit for Review Evidence, Cardiff University, United Kingdom; Wales Centre for Evidence Based Care, Cardiff University, United Kingdom; Health and Care Research Wales Evidence Centre, Cardiff University, United Kingdom; Health and Care Research Wales Evidence Centre, Bangor University, United Kingdom

## Abstract

Special procedures, such as tattooing (including semi-permanent make-up), body piercing, acupuncture and electrolysis involve piercing the skin and carry a potential risk of infection and other complications. This review provides an up-to-date evidence base on the main physical health risks associated with these special procedures.

Searches were conducted for research published between 2015 and 2023. Where possible, existing reviews were used. For tattooing, one systematic review including 55 studies was identified. For semi-permanent make-up, 37 cases presented in 31 case reports were identified. For body piercing, four systematic reviews were identified, which covered 174 studies. For acupuncture, one evidence map of 535 systematic reviews without age limits and one overview of 24 systematic reviews focused on children were included. For electrolysis, one case report was identified.

Complications and adverse events resulting from special procedures include fungal infections, bacterial infections, viral infections, blood-borne infections, allergic reactions, malignant growths, benign growths, sarcoidosis-related reactions, and a range of other skin and eye-related adverse reactions and dental issues.

**Research Implications and Evidence Gaps:** Most of the primary evidence for the physical health risks associated with the special procedures came from case reports and case series, which do not allow causal inferences or provide information on the prevalence of adverse events. Future primary research should employ more robust designs to gather evidence about the causal links between special procedures and adverse physical health events as well as about the prevalence of such events in people receiving special procedures. More evidence is needed to identify physical health risks associated with electrolysis.

**Policy and Practice Implications:** The evidence contained in this review will be used to inform the training of local authority enforcement officers and special procedures practitioners and to educate members of the public who seek to use these services. Due to the nature of the special procedures which involves piercing the skin and poses the risk of introducing infections to the body, high standards of hygiene may reduce the rate of infection.

**Funding statement:** The Wales Centre for Evidence Based Care, and the Specialist Unit for Review Evidence, were funded for this work by the Health and Care Research Wales Evidence Centre, itself funded by Health and Care Research Wales on behalf of Welsh Government.

## Definitions

***Acupuncture*** is the insertion of one or several needles into the skin at specific sites or acupuncture points for therapeutic purposes (Ernst 2010).

***Acupoint catgut embedding*** is the implantation of absorbable catgut sutures at acupoints (Inprasit et al. 2020).

***Auricular acupuncture*** is acupuncture of the ear (Kay Garcia & Chiang 2007).

***Electroacupuncture*** is a type of acupuncture in which a weak electric current is passed through acupuncture needles into the skin (Cai & Yang 2019).

***Dry needling therapy*** is the insertion of thin needles into the body (Dunning et al. 2014).

***Electrolysis*** is a permanent method of hair removal and involves inserting a small needle or probe into the hair follicle and passing an electric current through it, aiming to destroy the follicle to prevent hair regrowth (Fernandez et al. 2013).

***Microblading*** is a semi-permanent cosmetic procedure used to enhance the appearance of eyebrows. It is a form of eyebrow tattooing where pigment is introduced into the skin using fine needles to create hair-like strokes on the skin (Wong & Heacock 2018).

***Micropigmentation*** is a cosmetic procedure that involves the application of pigments to the skin to create semi-permanent or permanent make-up (De Cuyper 2008). The difference between tattooing and semi-permanent skin colouring is based on the depth at which ink is placed within the skin.

## 1. BACKGROUND

### 1.1 Who is this review for?

This Rapid Review was conducted as part of the Health and Care Research Wales Evidence Centre Work Programme. The above question was suggested by the Public Health Directorate of Welsh Government in light of the implementation of Part 4: Special Procedures of the Public Health (Wales) Act 2017. The Act creates a mandatory licensing scheme for carrying out special procedures, which include tattooing, body piercing, acupuncture and electrolysis.

Given the licensing scheme is due to come into force in 2024, the requested review updates the evidence base for the regulations to provide assurance that the risks associated with these procedures remain relevant to the proposed requirements of the licensing scheme and to inform how the scheme will work in practice.

### 1.2 Background and purpose of this review

The prevalence of individuals in the UK receiving tattoos, including semi-permanent skin make-up such as microblading and micropigmentation, as well as cosmetic piercings, electrolysis and acupuncture, collectively known as “special procedures”, is on the rise. Each of these special procedures involves piercing the skin and as a consequence, all of these procedures carry a potential risk of infection (Royal Society for Public Health 2019).

Special procedures can lead to infection from endogenous and exogenous agents (Patel & Cobbs 2015). Endogenous agents are microorganisms that naturally reside on the skin and these do not normally cause problems. However, if the skin barrier is broken or disrupted during the procedure, they can enter the skin and cause infections at the site where the skin is pierced. Common causes of endogenous infection include streptococci, staphylococci and pseudomonas (Patel & Cobbs 2015). The second cause of infection arises from exogenous agents, which are microorganisms not naturally present on the skin. These agents are introduced during the procedure through contaminated tools or equipment, such as a dirty needle, and can lead to the transmission of blood-borne viruses, for example, viral hepatitis, tuberculosis, syphilis and HIV (Patel & Cobbs 2015, Royal Society for Public Health 2019).

Many complications related to tattooing or the application of semi-permanent make-up are caused by acute allergies and delayed hypersensitivity reactions, primarily triggered by the ingredients present in the tattoo inks or trauma during tattooing or removal procedures. These non-specific reactions, often worsened by exposure to sunlight, are unpredictable and may occasionally manifest after a long period of time (even decades), leading to chronic complications associated with underlying auto-immune conditions (Piccinini et al. 2016).

Consequently, it is crucial for practitioners to adhere to safe working practices and have good infection control measures in place so that both clients and practitioners can be safeguarded and protected (Chartered Institute of Environmental Health 2013). Additionally, the increasing prevalence of antimicrobial-resistant infections, responsible for a significant number of fatalities each year in Europe and the USA alone (estimated to be at least 50,000 lives), underscores the critical importance of minimising infections associated with special procedures (O’Neill 2014).

The regulations and laws concerning special procedures vary across the different regions of the UK. In Wales in 2017, the Public Health (Wales) Act was enacted (Welsh Government 2017) and under Part 4 of the Act, special procedures were categorised as follows: tattooing, which includes both traditional tattooing and semi-permanent make-up such as microblading and micropigmentation; piercing, which covers various body piercing practices; electrolysis, involving the removal of hair using electric currents; and acupuncture, which includes dry needling techniques (National Assembly for Wales 2017). The legislation ensures public safety by establishing clear guidelines for practitioners, thus minimising health risks associated with the special procedures.

As part of the final phase of the changes introduced under the Public Health (Wales) Act 2017, Wales is set to become the first UK nation to introduce a mandatory national licensing scheme for the providers of special procedures (Welsh Government 2023). In preparation for this process, this Rapid Review provides an up-to-date evidence base on the main physical health risks that may be associated with tattooing, body piercing, acupuncture and electrolysis. As such, it answers the following questions:

Q1a. What are the physical health risks associated with tattooing (including semi-permanent make-up)?
Q1b. What are the physical health risks associated with semi-permanent make-up (i.e., microblading and micropigmentation)?
Q2. What are the physical health risks associated with body piercing?
Q3. What are the physical health risks associated with acupuncture?
Q4. What are the physical health risks associated with electrolysis?

Preliminary searches identified a large volume of existing systematic reviews in this field. The current review therefore utilised, where possible, existing systematic reviews or overviews or maps of existing reviews. Where there was no existing review, a review of new research (primary studies) was conducted.

## 2. RESULTS

This section details the extent of the evidence base and provides an overview of the included research reporting adverse events or complications experienced by individuals who have received a tattoo, semi-permanent make-up, body piercing, acupuncture or electrolysis as identified in the literature. The research evidence is summarised separately for each of the special procedures: tattooing, semi-permanent make-up, body piercing, acupuncture and electrolysis.

The overview of the evidence base provides a description of the characteristics of the available evidence, including, where such information was available, study designs, the dates of the studies, demographic information of the participants, the puncture site, the type of practitioner undertaking the procedure, the countries where the procedures were performed, the outcomes of interest and the time frame of when an adverse reaction or complication was experienced or reported.

Summaries of the reported clinical presentations (signs and symptoms of a medical problem), microbiological complications and non-microbiological complications are presented in Tables 1–3. A glossary of the terms associated with the adverse events is available in Appendix 1.

Microbiological complications in the context of special procedures refer to adverse reactions related to the presence of growth of microorganisms (such as bacteria, viruses or fungi) in or around the puncture site. Non-microbiological complications related to special procedures are health issues or adverse reactions that can arise from the special procedure processes themselves or the tattooing ink but that do not involve microorganisms.

At the end of each section dedicated to a particular special procedure, the quality of the included evidence based on the relevant quality appraisal checklists is summarised. Details of how quality was assessed are provided in Section 5.5. Quality appraisal tables are reported in Section 6.3, with Table 16 providing a summary of the critical appraisal scores of the included SRs, evidence map and overview of reviews and Table 17 of the case reports.

The detailed eligibility criteria for this review are presented in Tables 4–8 in Section 5.1. The results of the searches are reported in Appendix 2, and the study selection process is presented in Table 9 in Section 6.1.

### 2.1 Tattooing

#### 2.1.1 Overview of the evidence base on tattooing

The searches identified one SR (Sindoni et al. 2021) that focused on tattooing, which included 55 studies (see Table 10). The study designs included 32 case reports, 13 case series, seven cross-sectional studies, two prospective cohort studies and one retrospective cohort study, which were published between 1991 and 2020. From 2015 to 2020 there were 15 case reports, eight case series and one cohort study. The locations of the tattoo on the body that were reported include the torso (seven studies), arm (nine studies), leg (six studies), face (three studies), buttock (one study), penis (one study) and multiple sites (ten studies). A further 18 studies did not report the location of the tattoo. The type of practitioner undertaking the procedures was not reported. The countries where the procedures took place were not reported. The outcomes were microbiological and non-microbiological adverse reactions as a result of tattoos in mainly adult populations (three studies included children under 18 years of age). The time frame for reporting or experiencing an adverse effect or complication, where reported, ranged from six days to two months for the studies that reported microbiological complications and from two weeks to over 10 years for the studies that reported non-microbiological complications.

The sex of participants across the case reports reporting single cases was split between males (16 studies) and females (14 studies). Two further case reports of three participants had a mix of both males and females. The sex of participants across the remaining study types (23 studies) were males (two studies), females (one study), a mix (seven studies) and 13 studies did not provide this detail.

#### 2.1.2 Physical health risks associated with tattooing

In the one included SR by Sindoni et al. (2021), the clinical presentations commonly reported after receiving a tattoo include the following: skin irritation, swollen glands near the site, bleeding, oedema (swelling), pruritus (itching), suppuration (production of pus), blistering, scabs, pain, fever, nausea and vomiting (see Table 1).

Across 10 of the included studies (five case reports and five case series) reporting **microbiological complications** in the SR (seven good quality, two poor quality and one fair quality), the most commonly reported bacterial infections included non-tuberculous *Mycobacterial* infections (60 cases), *Mycobacterium chelonae* infections (14 cases) and cutaneous *Mycobacterial* infections (four cases). Less commonly reported bacterial infections included life-threatening cellulitis (two cases), cutaneous diphtheria (two cases) and *staphylococcal* scalded skin syndrome infection (one case). Additionally, one study (case series) described the presence of warts. The review authors concluded that it is likely that these outbreaks can be linked to poor infection control practices and contaminated or diluted inks. Additionally, one instance of the blood-borne infection Hepatitis C Virus transmitted through the use of contaminated tattoo equipment was reported.

The SR identified two case series (good quality) and one case report (fair quality) that were published from 2015 to 2020 that reported bacterial and viral infections. These were non-tuberculous *Mycobacterial* infections (two cases), *Staphylococcal* infection (one case) and warts (one case).

Thirty-seven of the included studies (23 good quality case reports, four fair quality case reports, eight good quality case series, one fair quality cross sectional and one good quality cohort study) reported a wide variety of **non-microbiological complications**. The most frequently reported were allergic reactions such as contact dermatitis to the pigments or components in tattoo ink. Other non-microbiological complications that were described include cancerous growths, benign growth conditions, sarcoidosis-related reactions (including granulomatous reactions), skin-related adverse reactions, eye-related adverse reactions and other adverse reactions.

Findings from the case reports and case series have reported a number of different types of cancers occurring in some instances up to 16 years after a person has had a tattoo and include melanoma, squamous cell carcinoma, basal cell carcinoma, dermatofibrosarcoma protuberans and leiomyosarcoma. The benign growth conditions that have been described include haemangioma, epithelioid osteoblastoma and dermatofibroma. The sarcoidosis-related reactions that were described include sarcoidosis, sarcoid uveitis, cutaneous and pulmonary sarcoidosis, granulomatous infiltrate and granulomatous dermatitis.

A wide range of other skin-related adverse reactions have been described and include dermatitis (eczema), lichenoid reactions, inflammatory reactions, keratoacanthoma, nodular dermal lymphohistiocytic infiltration, pseudoepitheliomatous hyperplasia, pyoderma gangrenosum, Koebner phenomenon, scar formation, papulo-nodular reactions and skin thickening. Additionally, a diverse range of other adverse reactions described within single cases have also been reported, such as priapism linked to a tattoo of the penis, cutaneous and pulmonary sarcoidosis, lymphopathy and photosensitivity. Eye-related adverse effects that have occurred after receiving a lid-eye tattoo include short tear film break-up time, corneal erosion and meibomian gland loss, increased tear film instability and exacerbated signs and symptoms of ocular surface disease.

#### 2.1.3 The quality of the SR of tattooing

The included SR of tattooing (Sindoni et al. 2021) received eight out of 11 points on the relevant critical appraisal checklist. While the SR had no critical flaws (the search strategy, the criteria for appraising studies and the methods used to combine studies were appropriate, and the sources and resources used to search for studies were adequate), it had some weaknesses deemed non-critical for the purposes of the present review, such as the lack of assessment of the likelihood of publication bias. The SR was therefore considered to be of moderate quality. Information on how quality was derived is provided in Section 5.5. More detail on the quality of the included SR can be found in Table 16 in Section 6.3.

#### 2.1.4 Bottom line summary for tattooing

Evidence from one moderate quality SR (Sindoni et al. 2021) described a wide range of complications and adverse reactions experienced by individuals after getting a tattoo. These were classified as microbiological complications – bacterial infections, viral infections and blood-borne infections, and non-microbiological complications – allergic reactions, malignant growths, benign growths, sarcoidosis-related reactions (including granulomatous reactions), along with a wide range of other skin-related adverse reactions. The countries where the procedures took place were not reported.

Bacterial and viral infections reported in studies published since 2015 included non-tuberculous *mycobacterial* skin infections, *staphylococcal* scalded skin syndrome and warts.

### 2.2 Semi-permanent make-up

#### 2.2.1 Overview of the evidence base on semi-permanent make-up

The searches identified one SR (Motoki et al. 2020), see Table 11, which sought to retrieve primary research reporting pathological scaring after eyebrow micropigmentation. However, this SR did not find any such primary research.

Due to the lack of existing secondary research, a search for primary studies was conducted as part of the present review. It identified 31 case reports (see Table 12), which were published between 2015 and 2023 that reported a variety of adverse reactions. The sample size for each case report varied from one participant to five participants, with 28 case reports (90%) involving a single participant. In total, there were 37 individual cases presented. In the majority of cases (n=26; 84%), the specific practitioner responsible for the semi-permanent make-up procedure was not explicitly specified. Among the cases (six case reports) where the practitioner information was provided, two involved non-medical professionals, one mentioned an aesthetician, one identified a medical practitioner, and for one case, it was unclear whether the practitioner was a medical or non-medical professional. Across the cases, the majority (n=33, 89.2%) of the permanent make-up procedures were applied to eyebrows, two cases (5.4%) were confined to the lips, while one case (2.7%) was limited to the eyelids and a single case (2.7%) involved both the lips and eyebrows.

The timeframe between the semi-permanent make-up procedure and a person presenting at the hospital for a microbiological reaction across five cases ranged from the same day to one month. The time frame for the non-microbiological reactions varied significantly. In two cases (5.4%), reactions occurred within one to two days after the semi-permanent make-up procedure. In 15 cases (40.5%), reactions appeared within a range of one to 21 months. Additionally, five cases (13.5%) reported reactions occurring two to five years later, while four cases (10.8%) documented reactions emerging eight to 16 years after the procedure. In six cases (16.2%), the timeframe was not reported.

Participants across the case reports were all women, with varying ages ranging from 25 to 75 years old. Among the 31 case reports examined, only seven of them explicitly mentioned the ethnicity of the individuals, specifically Hispanic (three cases), Caucasian (one case), Chinese (one case), Japanese (one case) and Lithuanian (one case).

The majority (n=26, 84%) of the procedures occurred in what was assumed to be a commercial setting, although five case reports (16%) did not specify the setting. The case reports originated from a total of 18 countries and included Iran (n=4), the USA (n=4), two case reports each from Brazil, China, Japan, Germany, and Spain and one case report each from Canada, Colombia, Israel, Italy, Mexico, Republic of Ireland, Syria, the Netherlands, Tunisia, Turkey and the UK. The country of origin for two reports, specifically Hinojosa et al. (2017; Mexico or the USA) and Vera et al. (2018; Venezuela or Germany), was not clear.

#### 2.2.2 Physical health risks associated with semi-permanent make-up

Across the 37 cases presented in 31 case reports, the clinical presentations commonly reported after following the application of semi-permanent make-up are reported in Table 1 and include **erythema or redness** (Akoh et al. 2021, Bashinskaya et al. 2022, Gilhooley et al. 2020, Goldberg et al. 2018, Maier et al. 2015, Mirzaei et al. 2017, Naeini et al. 2017, Sano et al. 2021, Suleman et al. 2023, Tittelbach et al. 2018, Vera et al. 2018); **oedema or swelling** (Akoh et al. 2021, Gilhooley et al. 2020, Lahouel et al. 2022, Sano et al. 2021, Soltany et al. 2023), **pain** (Motoki et al. 2020) and **pruritus** (Abtahi-Naeini et al. 2019, Ibraheim et al. 2023, Lahouel et al. 2022, Motoki et al. 2020, Tittelbach et al. 2018, Valbuena et al. 2017, Vera et al. 2018).

Five case reports (Akoh et al. 2021, Castaño-Fernández & Grau-Pérez 2023, Marcelino et al. 2021, Sano et al. 2021, Soltany et al. 2023) reported **microbiological complications** in the context of semi-permanent make-up, which included both viral and bacterial infections. The viral infections reported were **monkeypox (mpox)** (Castaño-Fernández & Grau-Pérez 2023) and *Molluscum contagiosum* (Marcelino et al. 2021) and the bacterial infections reported were **orbital cellulitis** (Soltany et al. 2023), three cases of **periorbital cellulitis** (Akoh et al. 2021, Soltany et al. 2023, Sano et al. 2021) and **necrotising fasciitis** (Soltany et al. 2023).

The **non-microbiological adverse reactions** that have been described in the context of semi-permanent make-up included 25 cases (across 19 case reports) of **sarcoidosis-related reactions** (Bashinskaya et al. 2022, Bombonato et al. 2015, Ebrahimiadib et al. 2021, Gilhooley et al. 2020, Hirai et al. 2022, Huisman et al. 2019, Ibraheim et al. 2023, Iwayama et al. 2017, Mirzaei et al. 2017, Naeini et al. 2017, Nie et al. 2022, Tukenmez Demirci et al. 2016, Valbuena et al. 2017, Vera et al. 2018, Tittelbach et al. 2018); 17 cases of **granulomatous reactions, including papules** (Bashinskaya et al. 2022, Ebrahimiadib et al. 2021, Hinojosa et al. 2017, Hirai et al. 2022, Huisman et al. 2019, Ibraheim et al. 2023, Iwayama et al. 2017, Leight-Dunn et al. 2022, Maier et al. 2015, Naeini et al. 2017, Nie et al. 2022, Suleman et al. 2023, Sun & Lao 2023, Tittelbach et al. 2018, Tukenmez Demirci et al. 2016, Valbuena et al. 2017); three cases of **allergic reactions** (Lahouel et al. 2022, Ibraheim et al. 2023, Motoki et al. 2020), 19 cases of **skin-related adverse reactions** (Abtahi-Naeini et al. 2019, Akoh et al. 2021, Bashinskaya et al. 2022, Ebrahimiadib et al. 2021, Gilhooley et al. 2020, Hinojosa et al. 2017, Goldberg et al. 2018, Hirai et al. 2022, Maier et al. 2015, Mirzaei et al. 2017, Naeini et al. 2017, Marcelino et al. 2021, Navarro-Triviño et al. 2021, Nie et al. 2022, Sano et al. 2021, Suleman et al. 2023, Tierney & Kavanagh 2021, Tittelbach et al. 2018, Vera et al. 2018) and five cases of **eye-related adverse reactions and other adverse reactions** (Sano et al. 2021, Goldberg et al. 2018, Hinojosa et al. 2017, Soltany et al. 2023, Navarro-Triviño et al. 2021).

Sarcoidosis is a multisystem inflammatory disorder characterised by the presence of granulomas in various organs and tissues throughout the body. Five cases describe **cutaneous sarcoidosis** (Bashinskaya et al. 2022, Hinojosa et al. 2017, Ibraheim et al. 2023, Naeini et al. 2017, Tukenmez Demirci et al. 2016), five cases describe **systemic sarcoidosis, i.e. affecting multiple organs systems in the body** (Bashinskaya et al. 2022, Ebrahimiadib et al. 2021, Hinojosa et al. 2017, Vera et al. 2018, Huisman et al. 2019), one case of **pulmonary sarcoidosis** (Naeini et al. 2017) and one case of **Löfgren’s syndrome**, an acute form of sarcoidosis (Mirzaei et al. 2017). A range of granulomatous reactions across the 17 cases were described using a variety of terms, for example, non-caseating granulomas, granulomatous dermatitis, chronic granulomatous inflammation and sarcoidal foreign body (granulomatous) reactions.

A number of skin-related adverse effects were described and these include **crusting** (Akoh et al. 2021, Hinojosa et al. 2017, Sano et al. 2021, Suleman et al. 2023), **cutaneous lymphoid hyperplasia** (Navarro-Triviño et al. 2021); **hyperpigmentation** (Abtahi-Naeini et al. 2019, Akoh et al. 2021), **hyperplasia** (Marcelino et al. 2021), **induration** (Maier et al. 2015), **irritation** (Goldberg et al. 2018), **lichenoid tattoo reactions** (Abtahi-Naeini et al. 2019), **pustules** (Akoh et al. 2021), **rash** (Hirai et al. 2022, Nie et al. 2022, Suleman et al. 2023), **scaling** (Ebrahimiadib et al. 2021, Maier et al. 2015, Tittelbach et al. 2018), **vitiligo arising from the koebnerisation effect of microblading** (Tierney & Kavanagh 2021).

Additionally, there were eye-related adverse reactions described which included **bilateral eye swelling, redness, and pain** (Sano et al. 2021), **conjunctival injection** (Goldberg et al. 2018, Hinojosa et al. 2017), **irritation** (Goldberg et al. 2018), **right eye proptosis and painful eye movement** (Soltany et al. 2023) **uveitis** (Navarro-Triviño et al. 2021) and **pigmented stain on the eye** (Goldberg et al. 2018).

The non-microbiological **allergic reactions** were reported in relation to the pigments or components of the semi-permanent make-up inks (for example, iron oxide in Leight-Dunn et al. 2022) or to the tattoo being performed with nickel containing needles (Lahouel et al. 2022).

#### 2.2.3 The quality of the studies on semi-permanent make-up

The quality rating for the case reports of complications associated with semi-permanent make-up ranges from four to eight points out of eight, indicating varying levels of methodological rigour and reporting quality in the included case reports.

Demographic characteristics, the clinical condition of the patient on presentation and diagnostic tests or assessment methods and their results were clearly described in all case reports. All but one case report clearly described the patient’s history and presented it as a timeline. More detail can be found in Table 17 in Section 6.3.

Seven case reports achieved the highest rating of eight out of eight on the relevant critical appraisal checklist (Akoh et al. 2021, Gilhooley et al. 2020, Goldberg et al. 2018, Hinojosa et al. 2017, Leight-Dunn et al. 2022, Mirzaei et al. 2017, Soltany et al. 2023, Sun & Lao 2023).

Seventeen case reports received a rating of seven out of eight (Abtahi-Naeini et al. 2019, Ebrahimiadib et al. 2021, Hirai et al. 2022, Huisman et al. 2019, Ibraheim et al. 2023, Iwayama et al. 2017, Lahouel et al. 2022, Maier et al. 2015, Marcelino et al. 2021, Motoki et al. 2020, Naeini et al. 2017, Navarro-Triviño et al. 2021, Nie et al. 2022).

Three case reports earned a score of six out of eight (Sano et al. 2021, Tierney & Kavanagh 2021, Valbuena et al. 2017). Three case reports were rated five out of eight (Bashinskaya et al. 2022, Castaño-Fernández & Grau-Pérez 2023, Vera et al. 2018). Finally, a single case report was rated four out of eight (Bombonato et al. 2015).

#### 2.2.4 Bottom line summary for semi-permanent make-up

Evidence from 31 case reports reporting on 37 individual cases described a diverse spectrum of complications and adverse reactions experienced by individuals following the application of semi-permanent make-up. These can be broadly classified as microbiological complications (bacterial infections and viral infections) and non-microbiological complications (sarcoidosis-related reactions, including granulomatous reactions, allergic reactions and a range of other skin and eye-related adverse reactions).

The countries where the procedure took place were Brazil, Canada, China, Colombia, Germany, Iran, Israel, Italy, Japan, Mexico, the Republic of Ireland, Spain, Syria, the Netherlands, Tunisia, Turkey, the UK and the USA.

Bacterial and viral infections reported in studies published since 2015 included *Molluscum contagiosum,* orbital cellulitis, periorbital cellulitis, necrotising fasciitis and monkeypox.

### 2.3 Body piercing

#### 2.3.1 Overview of the evidence base on body piercing

The searches identified four SRs (Acuña-Chavez et al. 2022, Hennequin-Hoenderdos et al. 2016, Passos et al. 2022, Sindoni et al. 2022) that included 174 studies published between 1973 and 2020 focusing on body piercing (see Table 13). There was a moderate overlap across the SRs, meaning that 30 primary studies were included in more than one SR. More details about overlap can be found in Section 5.6. All of the information that follows is based on the 174 studies.

Two of the SRs included adults and children and the remaining two SRs did not provide this detail. Three SRs provided details of the sex of the participants and between 61% and 85% were female. None of the SRs reported the ethnicity of the participants. Three SRs reported the country where the body piercing took place.

The study designs included before-after design (one study), case-control (23 studies), case report (62 studies), case report in a cross-sectional study (one study), case series (15 studies), cohort (five studies), cross-sectional (65 studies, out of which 26 were prevalence studies), quasi-experimental (one study) and randomised controlled trial (one study). The SRs identified three case series, seven case reports, 11 cross-sectional studies (out of which four were prevalence studies), four case-control studies, one retrospective cohort study, and one quasi-experimental study that were published between 2015 and 2020.

The research was conducted across 31 different countries with just under a third of the studies being conducted in the USA (50 studies) and only six in the UK. Two SRs focused on oral piercing (Hennequin-Hoenderdos et al. 2016, Passos et al. 2022), one on nipple piercing (Acuña-Chavez et al. 2022) and one on piercing in general (Sindoni et al. 2022). Across all the SRs, there were 213 puncture (piercing) sites, which included the ear (22 studies), nipple (28 studies), tongue (68 studies), lip (44 studies), tongue and lip (nine studies), oral (15 studies), nose (one study), genital (one study) and navel (one study). Puncture (piercing) site was not reported in a further 24 studies. The type of practitioner undertaking the procedures was not reported.

Three of the four SRs (Sindoni et al. 2022, Acuña-Chavez et al. 2022, Passos et al. 2022) reported on the time frame for experiencing an adverse reaction or complication. Acuña-Chavez et al. (2022) and Passos et al. (2022) made no distinction when reporting microbiological and non-microbiological effects and the time frame ranged from immediate (n=11) to less than six months (n=13) and over six months (n=16), three of the 16 being over one year. The time frame wasn’t reported in a further 41 studies. Sindoni et al. (2022) made a distinction when reporting microbiological and non-microbiological effects. For the studies reporting microbiological effects, the time frame ranged from within the first month (n=19), one to six months (n=13), six to 12 months (n=3) and over one year (n=8), with one study as late as eight years after the procedure. The time frame was not reported in a further ten studies. For the studies reporting non-microbiological effects, the time frame varied between less than one year (n=1) to over one year (n=5), with a further five studies not reporting. For the studies reporting both microbiological and non-microbiological effects, the time frame ranged from less than one year (n=3) to over one year (n=5), with a further 12 studies not reporting it.

The outcomes studied were most frequently isolated bacteria, microbiological complications, non-microbiological complications and the occurrence of clinical manifestations/ complications, oral lesions related to soft tissue and mucosa, periodontal and tooth damage and alterations (speech, mastication, deglutition and taste-related, saliva and galvanic current, temporomandibular disorders, soft plaque and calculus formation and microbiology).

#### 2.3.2 Physical health risks associated with body piercing

Across the four SRs (Acuña-Chavez et al. 2022; Hennequin-Hoenderdos et al. 2016; Passos et al. 2022, Sindoni et al. 2022), the clinical presentations commonly reported after having undergone body piercing varied according to the site of the piercing. Specifically, those who had nipple piercings described breast fluid collection, breast pain or tenderness, breast swelling, deformity and discharge, as well as similar presentations to those having a piercing in other parts of the body, including erythema, fever, headache, hyperpigmentation, oedema, productive cough with bloody sputum, pruritus, swollen glands and syncope (see Table 1).

Three SRs reported **microbiological complications** in individuals who have undergone body piercing (Sindoni et al. 2022, Acuña-Chavez et al. 2022, Passos et al. 2022), which included fungal infections (*Candida dubliniensis*), viral infections (viral hepatitis and Herpes simplex hepatitis) and bacterial infections. The bacterial infections that were described included cephalic tetanus, cerebellar brain abscess, chest wall cellulitis/retroareolar cellulitis, endocarditis, glomerulonephritis, mastitis and toxic shock syndrome and infections (unspecified). The commonly occurring organisms identified as causative agents for infections were *Staphylococcus* and non-tuberculous *Mycobacterium*. Sixteen rarer types of organisms were identified as causative agents of infections in people who received a body piercing (for details, see Table 2). Information on the prevalence of infections was not available.

The three SRs included two case series, five case reports, five cross-sectional studies (out of which two were prevalence studies) and one retrospective cohort study that were published from 2015 to 2020 that reported bacterial and fungal infections. The bacterial and fungal infections that were described included retroareolar cellulitis (one case), unspecified infections (five cross-sectional studies) and *Candida dublinensis* (one case). The commonly occurring organisms identified as causative agents for infections included *Neisseria gonorrhoeae* (two cases), *Propionibacterium acnes* (two case series n=11) and *Staphylococcus epidermidis* (one case). Rarer types of organisms that were identified as causative agents in people who received a body piercing included *Pseudomonas aeruginosa* (one retrospective cohort study), *Mycobacterium fortuitum* (one case), *Staphylococcus epidermidis* alongside *Actinomyces turicensis* and *Peptoniphilus harei* (one case), Coagulase-negative *Staphylococci* (one case series n=4), *Corynebacterium amycolatum* (one case series n=4)*, Haemophilus parainfluenzae* (one case series n=4), and a rare gram-positive cocci not otherwise specified (one case series n=4).

The **non-microbiological adverse reactions** that have been described in the context of body piercing include allergic reactions, malignant growths (basal cell carcinoma), benign growths (fibroma) and a wide range of other skin-related adverse reactions (acne, cysts, eczema (dermatitis), inflammatory reactions, scarification/keloid formation, induration and skin tearing) (Passos et al. 2022, Sindoni et al. 2022). The allergic reactions included contact dermatitis and nickel sensitisation (body piercing tattoo being performed with nickel-containing needles).

Additionally, in relation to tongue and lip piercings, a number of specific adverse reactions were described, which were oral and mucosal lesions, periodontal issues, teeth damage and alterations (changes), and these are reported in Table 3. Two SRs reported relative risks (RR), odds ratios (ORs) and event rates (ERs) to the incidence of oral and teeth injuries and the association between the presence of oral piercings and gingival recessions or dental alterations (Hennequin-Hoenderdos et al. 2016, Passos et al. 2022). Based on these SRs, the event rate within a pierced population indicated that 33% of participants with an oral piercing had gingival recession (Passos et al. 2022). Compared to the population without a piercing, the incidence of gingival recession was 2.77 times more likely in people with tongue piercing (RR = 2.77, 95% CI 1.99 to 3.85; p<0 .001) and 4.14 times more likely with a lip piercing (RR = 4.14, 95% CI 1.54 to 11.13; p=0.005; Hennequin-Hoenderdos et al. 2016). This is supported by findings from Passos et al. (2022), who found that the odds of developing gingival recession were seven times higher in people with an oral piercing than in individuals without it (OR = 7.085; 95% CI 4.252 to 11.805; p<0 .001).

The event rate of dental fracture (ER = 0.338; 95% CI 0.248 to 0.440), wear or abrasion (ER 0.344; 95% CI 0.183 to 0.553) in individuals with oral piercing was 34%, followed by 27% of dental damage (lesion not reported in detail) (ER = 0.270; 95% CI 0.074 to 0.630), and 22% of tooth chipping or enamel infraction (Passos et al. 2022). Incidence of tooth injury (including chipped/cracked/broken teeth, tooth wear or fractures) compared to unpierced individuals was 2.44 times more likely with a tongue piercing (RR = 2.44, 95% CI 1.35 to 4.41; p=0.003), although it was not significantly more likely with a lip piercing (RR = 1.33, 95% CI 0.74 to 2.41; p=0.34; Hennequin-Hoenderdos et al. 2016). The odds of dental fracture were three times higher in people with an oral piercing compared to individuals without it (OR = 3.293; 95% CI: 1.868 to 5.807; p <0.001), although no significant association was detected between tooth chipping or enamel infraction and oral piercings (OR = 2.223; 95% CI 0.737 to 6.775; p=0.156; Passos et al. 2022).

A diverse range of other adverse reactions were described for people undergoing body piercing and these include adornment aspiration. Adverse effects also included nipple ring ripped out, embedded earrings, lymphadenopathy, rejection and a split tongue.

#### 2.3.3 The quality of the SRs of body piercing

The quality of the SRs of body piercing is as follows. Out of the four included SRs, one received the highest score of 11 out of 11 on the relevant critical appraisal checklist (Passos et al. 2022). It was therefore deemed to be high quality. Two scored nine out of 11 (Acuña-Chavez et al. 2022, Sindoni et al. 2022). Neither had any flaws considered critical for the purposes of the present review, but they had non-critical weaknesses, such as the lack of assessment of the likelihood of publication bias, so they were considered to be of moderate quality. Finally, one SR scored eight out of 11 (Hennequin-Hoenderdos et al. 2016). It was considered to be of low quality because one critical flaw was identified (the methods used to combine studies were not considered appropriate), as well as some non-critical weaknesses. More detail can be found in Table 16 in Section 6.3.

#### 2.3.4 Bottom line summary for body piercing

Evidence from four SRs (Acuña-Chavez et al. 2022 – moderate quality; Hennequin-Hoenderdos et al. 2016 – low quality; Passos et al. 2022 – high quality, Sindoni et al. 2022 – moderate quality) described a wide range of complications and adverse reactions experienced by individuals who had undergone body piercing. These were classified as microbiological complications (fungal, viral and bacterial infections) and non-microbiological complications (allergic reactions, malignant growths, benign growths, and other skin adverse reactions). The countries where the procedures took place were Argentina, Austria, Australia, Belgium, Brazil, Canada, Cuba, France, Germany, Greece, Ireland, Israel, Italy, Kenya, Mexico, New Zealand, Pakistan, Poland, Saudi Arabia, Slovenia, South Africa, Spain, Sudan, Sweden, Switzerland, the Netherlands, Turkey, UK, Ukraine, USA and Venezuela.

Bacterial and fungal infections reported in studies published since 2015 included retroareolar cellulitis, unspecified infections and *Candida dublinensis*. Common causative agents for infections were identified and included *Neisseria gonorrhoeae*, *Propionibacterium acnes* and *Staphylococcus epidermidis.* Rare causative agents for infections were identified and included *Pseudomonas aeruginosa, Mycobacterium fortuitum, Staphylococcus epidermidis* alongside *Actinomyces turicensis* and *Peptoniphilus harei,* Coagulase-negative *Staphylococci, Corynebacterium amycolatum, Haemophilus parainfluenzae* and rare gram-positive cocci not otherwise specified.

Following oral piercings, including tongue and lip, a range of non-microbiological complications were reported, which included soft tissue and mucosal lesions, periodontal issues, teeth damage and alterations (changes).

### 2.4 Acupuncture

#### 2.4.1 Overview of the evidence base on acupuncture

The searches identified five overviews of existing SRs (Yang et al. 2015, Chan et al. 2017, Vieira et al. 2018, Kwon et al. 2019, Xu et al. 2023) that focused on acupuncture. To avoid repetition, two overviews of SRs deemed the most exhaustive were selected for this report: one evidence map of 535 SRs with no age limit for participants (Xu et al. 2023) and one overview of 24 SRs focused on children up to 18 years old, of which six reported adverse reactions (Yang et al. 2015) – see Table 14 for detail. Among the six SRs reporting adverse reactions included in Yang et al. (2015), one was also included in Xu et al. (2023). As for the overviews of SRs identified but not reported here, Chan et al. (2017) included 17 SRs, Kwon et al. (2019) included 11 SRs, and Vieira et al. (2018) focused on auriculotherapy, including ear acupuncture, and included a total of 14 SRs. None of the three reviews were limited to children.

The evidence map of 535 SRs included both healthy people and people with 23 different disease types classified by ICD-11 (Xu et al. 2023). The included SRs were published between 1999 and 2022. It included 18 types of acupuncture, with the most popular being electroacupuncture (67 SRs), followed by manual acupuncture (47 SRs), acupoint catgut embedding (41 SRs), dry needling therapy (39 SRs), auricular acupuncture (22 SRs) and acupoint injection (14 SRs). Systematic reviews of acupuncture where there was no penetration of the skin were excluded. The age and sex of participants varied and were not reported for each included SR. The first authors of the SRs came from 18 different countries, mainly from China (336 SRs), followed by South Korea (90 SRs) and the USA (30 SRs). Twenty SRs came from the UK. The countries of origin of the primary studies included in the overview of SRs were not reported. The outcomes of interest included any adverse reactions experienced by individuals who have undergone acupuncture.

The six relevant SRs in the overview of SRs of acupuncture in children (Yang et al. 2015) included participants aged up to 18 years old with nocturnal enuresis (2 SRs), Autism (2 SRs), cerebral palsy (1 SR) and asthma (1 SR). The included SRs reported on unspecified acupuncture, needle acupuncture, tongue acupuncture, scalp acupuncture, heat-producing needling and laser acupuncture. However, one SR out of the six only reported on the adverse events experienced by individuals who have undergone with laser acupuncture and is not included here due to it being outside of the eligibility criteria. The first authors of four of the remaining SRs originated from China and one from Korea. The countries of origin of the primary studies were not reported. Any adverse reactions experienced by individuals who have undergone with acupuncture were included.

#### 2.4.2 Physical health risks associated with acupuncture

In the evidence map of SRs (Xu et al. 2023), adverse events commonly reported by individuals who have undergone acupuncture include the following: pain (144 SRs), bleeding or bruising (120 SRs), digestive system symptoms such as nausea or vomiting, loss of appetite, dry mouth, constipation, diarrhoea, dyspepsia and heartburn (46 SRs), erythema (19 SRs), tiredness (40 SRs), discomforts (31 SRs), headache (27 SRs), pruritus (23 SRs), aggravation of symptoms (14 SRs), numbness (13 SRs), fevers (10 SRs), palpitations (10 SRs), heat or sweating (8 SRs), menoxenia (5 SRs), mild oedema (swelling) (16 SRs) and blisters (3 SRs). Syncope was reported in 86 SRs. It was thought to be linked to stress and fear of acupuncture in patients as well as frail and seriously ill patients and those with excessive blood loss, a history of dizziness from acupuncture and an improper posture.

**Microbiological complications** included infections, which were reported in 19 SRs^1^. Xu et al. (2023) stated that infections occurred primarily because of unsterilised needles, repeated use of needles, or contact of the needlepoint with clothing, but no further information on the cause of infections or how the causation was derived was provided. All patients recovered after receiving treatment. According to the authors of the evidence map, the rate of infections related to acupuncture has been decreasing in recent years due to the rise of health consciousness and disinfection.

Eleven SRs that reported infections in people who had received acupuncture were identified through supplementary materials to Xu et al. (2023). They were published between 2015 and 2021. These SRs included between six and 61 primary studies (*M* = 24.64), but the publication dates of the primary studies or the search dates were not reported in the evidence map. The reported infections were classified by Xu et al. (2023) as unspecified infections (4 SRs), skin infections (4 SRs, including 1 SR reporting a local skin infection), local or systemic infection (1 SR), pulmonary infection (1 SR) and urinary system infection (1 SR, in patients with urinary retention). No further information about the infections was provided.

One SR included primary studies with children six to 14 months old with cerebral palsy. Participants’ age in the other included SRs was either reported as adults or not specified. Ten of the SRs came from China and one from the USA. All of the SRs included a control condition (e.g., other kinds of Chinese medicine, western medicine, rehabilitation training, unspecified usual care, unspecified other therapy, sham acupuncture, and waitlist control). The quality of these SRs was classified by Xu et al. (2023) as critically low (n=6), low (n=4) and high (n=1).

To gain a better understanding of what kind of infections have been reported in recent years in people who have undergone acupuncture, the SRs included in the evidence map (Xu et al. 2023) that reported infections and were published after 2015 (n=9) were retrieved at full-text and examined further, as a snapshot of the available evidence rather than a systematic report, which was not feasible due to the time constraints of this rapid review. Formal data extraction or quality appraisal of these SRs was not conducted. Two unique primary studies published since 2015 reporting infections (one reporting skin infection and one unspecified infection) were identified in those SRs, but neither of them specified the type of infection.

Regarding **non-microbiological complications** described in the evidence map of SRs (Xu et al. 2023), skin allergy reactions (14 SRs), haematomas (70 SRs), and induration (11 SRs) were reported. The evidence map also reported on adverse events such as neuromuscular disease (16 SRs) and motor disorders (2 SRs).

**Other adverse events** were reported in 373 SRs identified by the evidence map and included needle sticking, broken needles, and bent needles. No acupuncture-related adverse events were reported in 176 reviews and 120 reviews did not specify adverse events. In addition, 89 SRs stated that no adverse events were reported in the original studies.

Fifty-three SRs reported that acupuncture-related adverse events were associated with the practitioners, but no information on how the association was derived was provided in the evidence map. Severe needle-related adverse reactions were reported to be rare. The incidence of adverse events raged from 6.71%–8.6% and the incidence of serious adverse events was stated to be approximately 0.001%, but information on what was considered serious adverse events was not provided (Xu et al. 2023).

As per the five relevant SRs reporting adverse events included in the overview of acupuncture in children (Yang et al. 2015), the adverse reactions that were described for children include superficial bleeding (2 SRs) and mild pain (1 SR) as well as crying (2 SRs), painful bi-auricular stimulation (1 SR), eczema (1 SR) and heat or swelling while pressing (1 SR). The frequency of these adverse events was unclear. In four SRs, the relevance between adverse events and acupuncture was unclear, and one found no relevance.

#### 2.4.3 The quality of the evidence map and overview of SRs acupuncture

The evidence map of SRs (Xu et al. 2023) scored 11 out of 11 on the relevant critical appraisal checklist and was therefore deemed to be of high quality. The overview of SRs (Yang et al. 2015) scored 10 out of 11, and the identified weakness was not judged to be critical for the purposes of the present review, so the overall quality was considered to be high. More detail can be found in Table 16 in Section 6.3.

#### 2.4.4 Bottom line summary for acupuncture

Evidence from one high quality evidence map of 535 SRs (Xu et al. 2023) described a wide range of complications and adverse reactions experienced by individuals (mainly adults) who had undergone acupuncture. Microbiological complications included skin infections, local or systemic infections, pulmonary infections, urinary system infections, and otherwise unspecified infections. The most commonly described non-microbiological complications were skin-related adverse effects such as skin allergy reactions, haematoma, and induration. The incidence of serious adverse events was low at 0.001%. The countries where the procedures took place were not reported.

Evidence from one high quality overview of reviews (Yang et al. 2015) described the commonly reported clinical presentations and adverse events experienced by children who have undergone acupuncture and included instances of pain and bleeding as well as eczema, heat, and swelling. The countries where the procedures took place were not reported.

Infections reported in primary studies published since 2015 included skin infection and unspecified infection. The types of infection were not stated.

### 2.5 Electrolysis

#### 2.5.1 Overview of the evidence base on electrolysis

The searches did not identify any SRs that focused on electrolysis. Additional searches for primary research identified one case report (Morand et al. 2015) – see Table 15. The case was of a 35-year-old female from Canada. No further details were reported.

#### 2.5.2 Physical health risks associated with electrolysis

A diagnosis of cutaneous sarcoidosis (non-microbiological reaction) was described in the case report by Morand et al. (2015) of a patient who had been receiving electrolysis for many years.

#### 2.5.3 The quality of the studies on electrolysis

The single identified case report of electrolysis was rated six out of eight on the JBI critical appraisal checklist for case reports (Morand et al. 2015). It lost points because the patient’s history was not clearly described and presented as a timeline and because adverse events or unanticipated events during treatment were not identified and described. More detail can be found in Table 17 in Section 6.3.

#### 2.5.4 Bottom line summary for electrolysis

One case report described a non-microbiological complication (cutaneous sarcoidosis). The country where the procedure took place was Canada.

**Table 1.**
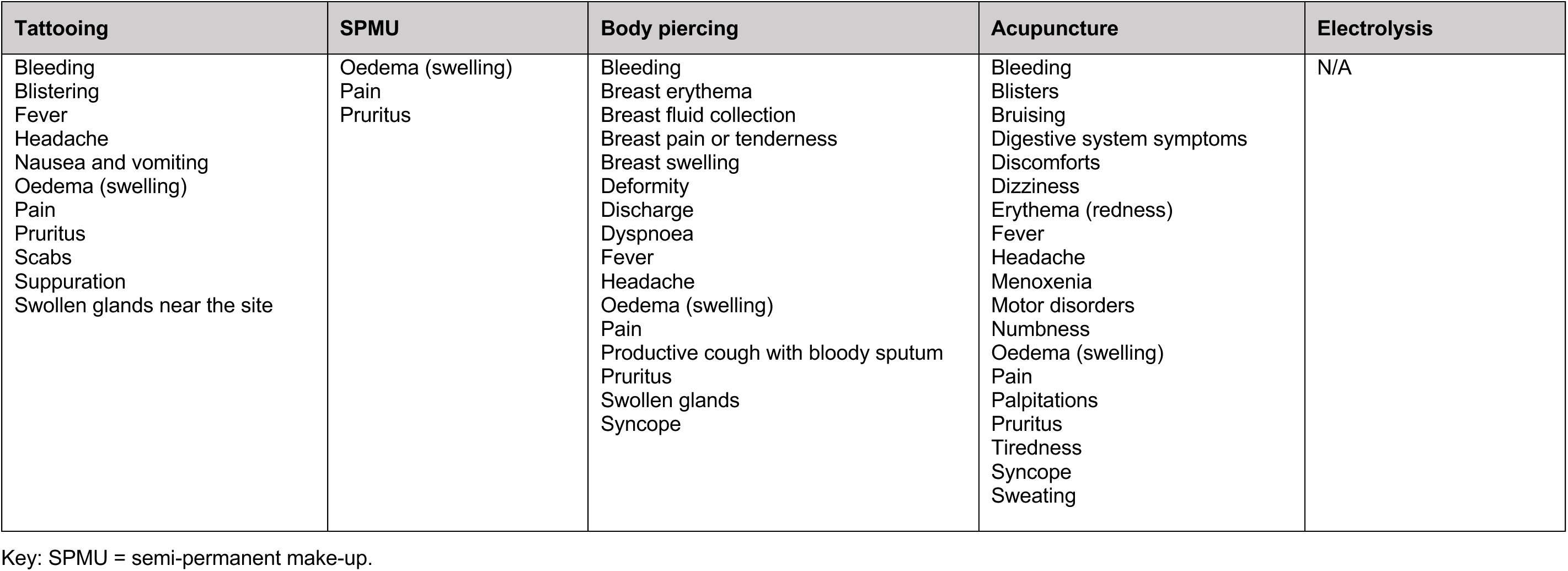
Summary of clinical presentations reported in people who had undergone special procedures.

**Table 2.**
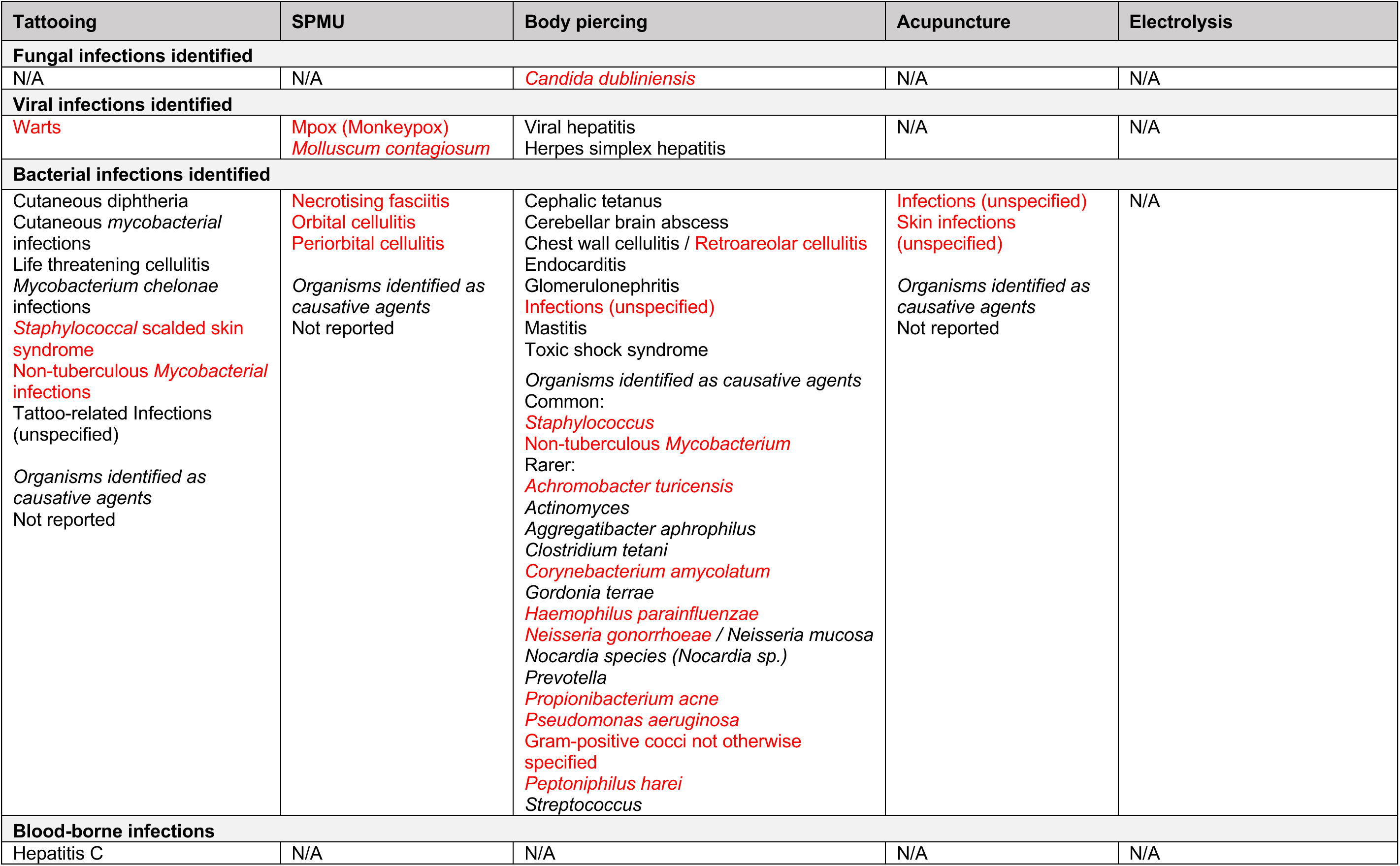
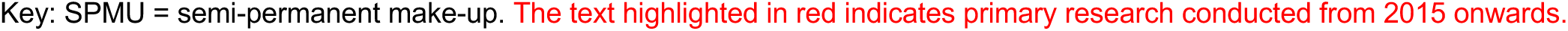
Summary of microbiological complications reported in people who had undergone special procedures.

| Tattooing | SPMU | Body piercing | Acupuncture | Electrolysis |
| --- | --- | --- | --- | --- |
| **Fungal infections identified** |  |  |  |  |
| N/A | N/A | *Candida dubliniensis* | N/A | N/A |
| **Viral infections identified** |  |  |  |  |
| **Warts** | *Mpox (Monkeypox)*<br>*Molluscum contagiosum* | Viral hepatitis<br>Herpes simplex hepatitis | N/A | N/A |
| **Bacterial infections identified** |  |  |  |  |
| Cutaneous diphtheria<br>Cutaneous *mycobacterial* infections<br>Life threatening cellulitis<br>*Mycobacterium chelonae* infections<br>*Staphylococcal* scalded skin syndrome<br>Non-tuberculous *Mycobacterial* infections<br>Tattoo-related Infections (unspecified)<br>*Organisms identified as causative agents*<br>Not reported | *Necrotising fasciitis*<br>*Orbital cellulitis*<br>*Periorbital cellulitis*<br>*Organisms identified as causative agents*<br>Not reported | Cephalic tetanus<br>Cerebellar brain abscess<br>Chest wall cellulitis / *Retroareolar cellulitis*<br>Endocarditis<br>Glomerulonephritis<br>*Infections (unspecified)*<br>Mastitis<br>Toxic shock syndrome<br>*Organisms identified as causative agents*<br>Common:<br>*Staphylococcus*<br>*Non-tuberculous Mycobacterium*<br>Rarer:<br>*Achromobacter turicensis*<br>*Actinomyces*<br>*Aggregatibacter aphrophilus*<br>*Clostridium tetani*<br>*Corynebacterium amycolatum*<br>*Gordonia terrae*<br>*Haemophilus parainfluenzae*<br>*Neisseria gonorrhoeae* / *Neisseria mucosa*<br>*Nocardia species (Nocardia sp.)*<br>*Prevotella*<br>*Propionibacterium acne*<br>*Pseudomonas aeruginosa*<br>Gram-positive cocci not otherwise specified<br>*Peptoniphilus harei*<br>*Streptococcus* | *Infections (unspecified)*<br>*Skin infections (unspecified)*<br>*Organisms identified as causative agents*<br>Not reported | N/A |
| **Blood-borne infections** |  |  |  |  |
| Hepatitis C | N/A | N/A | N/A | N/A |

Key: SPMU = semi-permanent make-up. The text highlighted in red indicates primary research conducted from 2015 onwards.

**Table 3.** Summary of non-microbiological complications reported in people who had undergone special procedures.

| Tattooing | SPMU | Body piercing | Acupuncture | Electrolysis |
| --- | --- | --- | --- | --- |
| **Allergic reactions** |  |  |  |  |
| Allergic reactions (unspecified)<br>Contact dermatitis<br>Dye allergy | Contact dermatitis<br>Nickel sensitisation<br>Reaction to iron oxide | Allergy (unspecified)<br>Contact dermatitis<br>Nickel sensitisation | Skin allergy reactions (unspecified) | N/A |
| **Malignant growths** |  |  |  |  |
| Basal cell carcinoma<br>Dermatofibrosarcoma protuberans<br>Leiomyosarcoma<br>Melanoma<br>Squamous cell carcinoma | N/A | Basal cell carcinoma | N/A | N/A |
| **Benign growths** |  |  |  |  |
| Epithelioid osteoblastoma<br>Dermatofibroma<br>Haemangioma | N/A | Fibroma | N/A | N/A |
| **Sarcoidosis-related reactions (including granulomatous reactions)** |  |  |  |  |
| Cutaneous sarcoidosis<br>Granulomatous dermatitis<br>Granulomatous infiltrate<br>Pulmonary Sarcoidosis<br>Sarcoid uveitis<br>Sarcoidosis | Chronic granulomatous inflammation (plaques)<br>Cutaneous sarcoidosis<br>Granulomatous dermatitis<br>Löfgren's syndrome<br>Non-caseating granulomas<br>Pulmonary sarcoidosis<br>Sarcoidal foreign body (granulomatous) reactions<br>Systemic sarcoidosis | Granulomatous dermatitis<br>Granulomatous perichondritis<br>Granulomatous tissue | N/A | Cutaneous sarcoidosis |
| **Skin-related adverse reactions** |  |  |  |  |
| Dermatitis (eczema)<br>Inflammatory reactions<br>Irritation<br>Keratoacanthoma<br>Lichenoid reaction<br>Nodular dermal lymphohistiocytic infiltration<br>Papulo-nodular reactions | Crusting<br>Cutaneous lymphoid hyperplasia<br>Erythema (Redness)<br>Hyperpigmentation<br>Hyperplasia<br>Induration<br>Irritation | Acne<br>Breast induration<br>Cysts<br>Eczema/dermatitis<br>Hyperpigmentation<br>Inflammatory reactions<br>Scarification/keloid formation<br>Skin tearing | Eczema<br>Haematoma<br>Induration | Papules |
| Pseudoepitheliomatous hyperplasia<br>Pyoderma gangrenosum Koebner phenomenon<br>Scar formation<br>Skin thickening | Lichenoid reaction<br>Pustules<br>Rash<br>Scaling<br>Vitiligo from koebnerisation | Papules |  |  |
| **Eye-related adverse reactions** |  |  |  |  |
| Sarcoid uveitis | Bilateral eye swelling, redness, and pain<br>Conjunctival injection<br>Irritation<br>Pigmented stain on the eye<br>Right eye proptosis and painful eye movement<br>Uveitis | N/A | N/A | N/A |
| **Oral and mucosal lesions** |  |  |  |  |
| N/A | N/A | Fibrous hyperplasia<br>Lesions on the hard palate<br>Lichenoid reactions<br>Macules, ulcers, papules, nodules and bruises<br>Mucosal atrophy<br>Soft tissue or mucosal injury | N/A | N/A |
| **Periodontal issues** |  |  |  |  |
| N/A | N/A | Bleeding on probing, clinical attachment loss, and probing depth<br>Dental mobility<br>Dentine hypersensitivity<br>Erythematous palatal mucosa<br>Gingival injury<br>Gingival recession<br>Gingivitis<br>Periodontitis<br>Tongue fissure | N/A | N/A |
| **Teeth damage** |  |  |  |  |
| N/A | N/A | Demineralized enamel<br>Dental hypersensitivity<br>Dental injuries/ damage (non-specific)<br>Enamel abrasion or abnormal tooth wear | N/A | N/A |
|  |  | Enamel abrasions<br>Enamel fractures<br>Nonspecific dental injuries<br>Tooth chipping<br>Tooth fracture |  |  |
| **Alterations (changes)** |  |  |  |  |
| N/A | N/A | Chewing<br>Halitosis<br>Microbiologic alterations in the mouth<br>Oral Galvanism<br>Saliva<br>Soft plaque or calculus formation<br>Speech<br>Swallowing<br>Taste<br>Temporomandibular joint dysfunction | N/A | N/A |
| **Other adverse reactions** |  |  |  |  |
| Lymphadenopathy<br>Photosensitivity<br>Priapism resulting from a tattoo of the penis | N/A | Adornment aspiration<br>Embedded earring<br>Lymphadenopathy<br>Nipple ring ripped out<br>Rejection<br>Split tongue | Motor disorder<br>Neuromuscular disease | N/A |

Key: SPMU = semi-permanent make-up.

## 3. DISCUSSION

### 3.1 Summary of the findings

The present rapid review summarised the available evidence on the physical health risks associated with special procedures, namely tattooing (including semi-permanent make-up), body piercing, acupuncture, and electrolysis.

All of the special procedures covered in this rapid review involve skin piercing, posing an infection risk. The skin acts as a natural defence against infections, and any breach in this defence can introduce bacteria or harmful microorganisms into the body. This rapid review has reported on a wide range of adverse events, including infections, in individuals who had undergone special procedures.

The wider literature reports that infections typically stem from two primary sources (Royal Society for Public Health 2019). The first source involves microorganisms that naturally inhabit the skin, known as “endogenous agents”, which are usually harmless but can cause infections when the skin’s protective barrier is compromised, and the second source is “exogenous agents”, which are not naturally found on the individual’s body but can be introduced, often through contaminated needles or ink, unsterilised equipment, unsanitary facilities, or practices that compromise hygiene standards (Royal Society for Public Health 2019).

This review identified a range of bacterial, viral, and fungal infections, referred to as microbiological complications, in individuals who had received special procedures. Bacterial infections that have been reported in studies published since 2015 included *Staphylococcal* scalded skin syndrome and non-tuberculous *Mycobacterial* skin infections (tattooing), necrotising fasciitis, orbital cellulitis, and periorbital cellulitis (semi-permanent make-up), retroareolar cellulitis (body piercing), as well as unspecified infections (body piercing, acupuncture). Viral infections reported in primary studies since 2015 included warts (tattooing) as well as monkeypox and *Molluscum contagiosum* (semi-permanent make-up), and fungal infections included *Candida dubliniensis* (body piercing). A wider range of infections, especially bacterial infections, was identified in the evidence published pre-2015 that was reported in the included reviews. The occurrence of infections is thought to decrease with improved disinfection practices, which highlights the importance of adhering to high standards of hygiene (Xu et al. 2023).

All special procedures carry the potential for allergic reactions, primarily due to the metals, especially nickel, used in needles (Royal Society for Public Health 2019). Tattoo ink can also trigger allergic reactions, leading to various manifestations such as allergic contact dermatitis (Royal Society for Public Health 2019). In this rapid review, such allergic reactions were experienced by individuals who had undergone special procedures with the exception of electrolysis (possibly due to the paucity of evidence).

Other non-microbiological reactions have been reported in people who had received special procedures as occurring anywhere up to 10 years later. These included malignant and benign growth (tattooing, body piercing), sarcoidosis-related reactions (tattooing, semi-permanent make-up, body piercing, electrolysis), skin-related adverse reactions (all types of special procedures), eye-related adverse reactions (tattooing, semi-permanent make-up) as well as oral and mucosal lesions, periodontal issues, and teeth damage and alterations (body piercing).

### 3.2 Strengths and limitations of the available evidence

The evidence included in this review covers a wide range of physical health issues in people who have undergone special procedures. The quality of the SRs and maps/overviews of SRs that covered adverse events associated with tattooing, body piercing, and acupuncture was generally good (three high quality reviews, three moderate quality, one low quality), with a critical flaw identified in only one review. This suggest that the included review evidence was able to identify and summarise the majority of the relevant literature that existed when the searches were carried out.

However, most of the primary research, both included in the SRs of tattooing and body piercing and identified as part of this review for semi-permanent make-up and electrolysis, did not employ designs that would have enabled it to gather information on the causal links between the special procedures and adverse physical health effects and on the prevalence of complications.

The evidence identified for semi-permanent make-up and electrolysis came solely from case reports. The majority of the primary studies included in the SRs of tattooing and body piercing also employed case report or case series designs, with some cross-sectional, cohort, before-after, and case-control studies and a total of only two studies with more robust designs (randomised controlled trial and quasi-experimental, both of body piercing). Case reports are considered among the least robust forms of evidence for inferring causality (Joanna Briggs Institute 2013), however, they are valuable sources of information on potential adverse reactions and complications, especially when other forms of evidence are not available. Because of the nature of the primary studies informing the evidence base for all of the special procedures, no estimate of how many people undergo special procedures and how many experience complications was available.

As for the evidence map and the overview of SRs of acupuncture, while the vast number of included SRs (535 SRs in the evidence map and 24 SRs in the overview, of which six reported adverse events) suggests that most of the secondary evidence in the field had been captured, the overlap of the included primary studies and their robustness were unclear.

### 3.3 Strengths and limitations of this Rapid Review

The main strength of this rapid review is that, employing a comprehensive search of available evidence across multiple sources (seven databases and almost 50 websites as well as review unpicking where relevant and forward and backward citation tracking), it has gathered all relevant evidence for physical health risks associated with special procedures in one place: secondary evidence for tattooing, body piercing, and acupuncture and primary evidence for semi-permanent make-up and electrolysis due to the lack of existing reviews. The evidence included in the present review was selected using a robust process, with two reviewers independently screening all citations. Measures were taken to ensure the accuracy of data extraction and quality appraisal of the available literature: these were conducted by one reviewer and checked by another.

However, it has a number of limitations. First of all, due to the volume of available literature and to avoid duplication of research effort, existing SRs were used for the evidence of physical health risks associated with tattooing and body piercing and maps/overviews of SRs for acupuncture. This meant that reporting in the present review depended on the level of detail and robustness of the evidence synthesis in the identified SRs or maps/overviews of SRs. As a result, the way that information was presented across the special procedures differed and some information that might be contained in the primary studies included in the SRs was not available. The limited time frame for this review in light of the amount of available evidence also meant that in-depth synthesis of the evidence was not possible, so high-level descriptive syntheses was carried out.

Regarding the topic of body piercing, as multiple SRs were included, overlap of their primary studies had to be considered. In this review, corrected covered area was calculated to determine overlap, which was found moderate. This means that some primary studies were included multiple times across the four SRs, so it is possible that some of the reported numbers of cases and adverse events of body piercing are overestimated in this rapid review. Determining the overlap between the primary studies included in the SRs reported in the overviews of SRs of acupuncture was not possible due to the volume of SRs and the time constraints.

### 3.4 Implications for policy and practice

The evidence contained in this review will not only be used to inform the training of local authority enforcement officers and special procedures practitioners, but to educate members of the public who seek to use these services.

It is important to note that the causal links between the special procedures and adverse events reported in this review could not be established due to the designs of the primary studies contributing to the evidence base. However, due to the nature of the special procedures which involves piercing the skin and poses the risk of introducing infections to the body, high standards of hygiene may reduce the rate of infection.

### 3.5 Implications for future research

All identified evidence for the physical health risks associated with semi-permanent make-up and electrolysis came from case reports. The majority of the primary studies included in the SRs of tattooing and body piercing were also case reports or case series. Future research may employ more robust study designs to improve the quality of available evidence and give an indication as to the causal links between these special procedures and adverse physical health events as well as to the prevalence of such events in people receiving special procedures.

The present review identified a paucity of evidence of the physical health risks associated with electrolysis. Only one case report of complications following electrolysis was found, indicating that this topic is under-researched. More evidence is needed to identify the risks associated with this procedure.

## Data Availability

All data produced in the present study are available upon reasonable request to the authors

## 5. RAPID REVIEW METHODS

### 5.1 Eligibility criteria

Eligibility criteria for each of the sub-questions are presented in Tables 4–8.

**Table 4.** Eligibility criteria: Tattooing.

|  | Inclusion criteria | Exclusion criteria |
| --- | --- | --- |
| **Population** | People of any age receiving tattoos on any body part |  |
| **Intervention/exposure** | Cosmetic skin tattooing (including semi-permanent make-up, i.e., microblading and micropigmentation) | Non-skin tattooing (e.g., corneal tattooing), medical tattooing |
| **Control/comparison** | N/A |  |
| **Outcome measures** | Any physical health risks | Mental health risks |
| **Setting/context** | Commercial/business setting | Healthcare setting |
| **Study design** | Systematic Reviews<br>Meta-analyses<br>Rapid Reviews using systematic search methods and critical appraisal | Primary studies<br>Narrative reviews |
| **Countries** | No geographic limits |  |
| **Language of publication** | English |  |
| **Publication date** | Since 2015 |  |
| **Publication type** | Published and grey literature | Letters, commentary, editorials, conference abstracts, protocols |
| **Other factors**<br>**Any other key points to note** | Contraindications will not be purposefully searched for but will be included in the RR report if described in the identified evidence for physical health risks. |  |

**Table 5.**
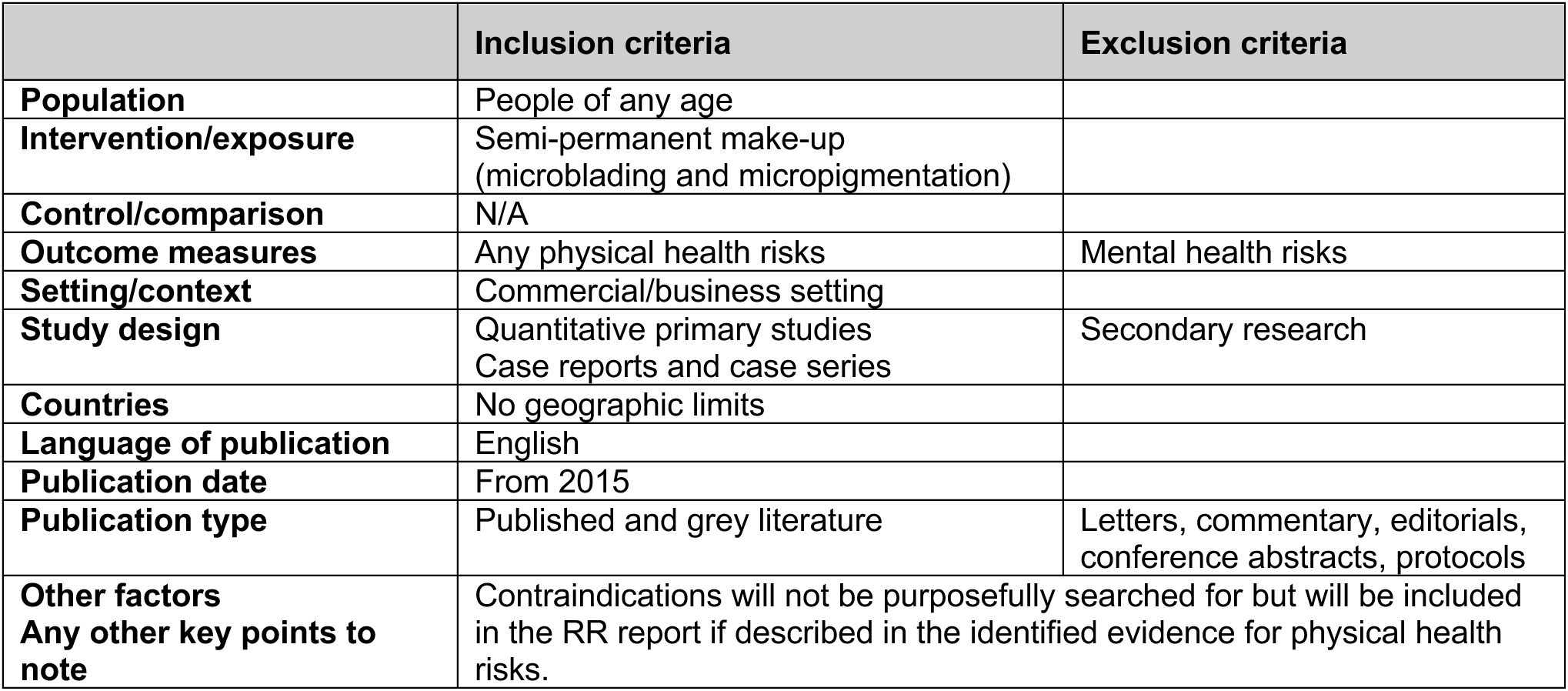
Eligibility criteria: Semi-permanent make-up.

**Table 6.** Eligibility criteria: Body piercing.

|  | **Inclusion criteria** | **Exclusion criteria** |
| --- | --- | --- |
| **Population** | People of any age receiving body piercings on any body part |  |
| **Intervention/exposure** | Cosmetic body piercing | Self-piercing |
| **Control/comparison** | N/A |  |
| **Outcome measures** | Any physical health risks | Mental health risks |
| **Setting/context** | Commercial/business setting |  |
| **Study design** | Systematic Reviews<br>Meta-analyses<br>Rapid Reviews using systematic search methods and including critical appraisal | Primary studies<br>Narrative reviews |
| **Countries** | No geographic limits |  |
| **Language of publication** | English |  |
| **Publication date** | From 2015 |  |
| **Publication type** | Published and grey literature | Letters, commentary, editorials, conference abstracts, protocols |
| **Other factors**<br>**Any other key points to note** | Contraindications will not be purposefully searched for but will be included in the RR report if described in the identified evidence for physical health risks. |  |

**Table 7.** Eligibility criteria: Acupuncture.

|  | **Inclusion criteria** | **Exclusion criteria** |
| --- | --- | --- |
| **Population** | General population of any age | Specific patient populations |
| **Intervention/exposure** | Needle acupuncture | Non-needle acupuncture (e.g., laser) |
| **Control/comparison** | N/A |  |
| **Outcome measures** | Any physical health risks | Mental health risks |
| **Setting/context** | Any setting |  |
| **Study design** | Overviews of systematic reviews and meta-analyses (umbrella reviews) | Primary studies<br>Systematic Reviews<br>Rapid Reviews<br>Scoping Reviews<br>Narrative reviews |
| **Countries** | No geographic limits |  |
| **Language of publication** | English |  |
| **Publication date** | From 2015 |  |
| **Publication type** | Published and grey literature | Letters, commentary, editorials, conference abstracts, protocols |
| **Other factors**<br>**Any other key points to note** | Contraindications will not be purposefully searched for but will be included in the RR report if described in the identified evidence for physical health risks. |  |

**Table 8.** Eligibility criteria: Electrolysis.

|  | Inclusion criteria | Exclusion criteria |
| --- | --- | --- |
| **Population** | People of any age receiving electrolysis on any body part |  |
| **Intervention/exposure** | Cosmetic electrolysis (hair removal technique) | Preoperative hair removal<br>Laser-based or light-based hair removal (photoepilation) |
| **Control/comparison** | N/A |  |
| **Outcome measures** | Any physical health risks | Mental health risks |
| **Setting/context** | Commercial/business setting | Healthcare setting |
| **Study design** | Quantitative primary studies<br>Case reports and case series | Secondary research |
| **Countries** | No geographic limits |  |
| **Language of publication** | English |  |
| **Publication date** | From 2015 |  |
| **Publication type** | Published and grey literature | Letters, commentary, editorials, conference abstracts, protocols |
| **Other factors**<br>**Any other key points to note** | Contraindications will not be purposefully searched for but will be included in the RR report if described in the identified evidence for physical health risks. |  |

### 5.2 Literature search

#### 5.2.1 Evidence sources

Comprehensive searches were conducted across the following databases for English-language publications from 2015 to August 2023:

- On the Ovid Platform: Medline, Embase, Ovid Emcare, Allied and Complementary Medicine Database (AMED)
- On the EBSCO Platform: Cumulative Index of Nursing and Allied Health Literature (CINAHL)
- Epistemonikos (Questions 1a, 2 and 3 only)
- Cochrane Database of Systematic Reviews (Questions 1a, 2 and 3 only)

The websites of key third sector and government organisations and accreditation bodies were also searched and can be seen in Appendix 3.

#### 5.2.2 Search strategy

An initial search of PUBMED was undertaken using the following keywords (tattoo* OR microblading* OR micropigmentation* OR makeup* OR pierc* OR electrolysis OR acupuncture) AND (adverse* or complication* or infectio* or allerg* or anaphylax* OR contra* OR sepsis OR harm OR safe*) This search was filtered by review or meta-analysis.

This initial search was followed by an analysis of the most relevant records to establish the most frequently used text words contained in the titles and abstracts and index terms used to describe articles. This informed the development of a comprehensive search strategy which was tailored for each information source. Full search strategies for all five questions are detailed in Appendix 2. Forward and backward citation tracking was completed using the citation chaser application (Haddaway et al. 2021), and relevant studies were added to the review.

#### 5.2.3 Peer review

To ensure the robustness and validity of the searches, we implemented a rigorous peer review process:

- The review team and a wider collaboration of stakeholders, including both information and subject specialists, were involved in the development, testing and reviewing of the search strategies prior to their final run.
- Test searches were assessed using “known relevant records”, and the searches were adjusted accordingly.
- The Peer Review of Electronic Search Strategies (PRESS) checklist was used (McGowan et al. 2016) to assess all elements of the search process.

#### 5.2.4 Reference management

All citations retrieved from the database searches were imported or entered manually into EndNote^TM^ (Thomson Reuters, CA, USA) and duplicates were removed. Following this process, the remaining citations were exported as a TXT file and imported to Rayyan^TM^, where any remaining duplicates were removed prior to sifting.

### 5.3 Study selection process

All citations were independently screened, using the information provided in the title and abstract, by two reviewers using the software package Rayyan^TM^, with any conflicts resolved by a third reviewer. For citations that appeared to meet the inclusion criteria or in cases in which a definite decision could not be made based on the title and/or abstract alone, the full texts were retrieved. The full texts were then screened for inclusion using a customised screening tool by two reviewers, with any disagreements resolved by a third reviewer. The flow of citations through each stage of the review process for each question is displayed using a table adapted from the PRISMA flow diagram, which can be found in Section 6.1. The list of studies excluded at the full-text stage and the reasons for exclusion are reported in Table 18 in Appendix 4.

### 5.4 Data extraction

All relevant data were extracted directly into tables by one reviewer and checked by another. The data extracted included specific details about the populations, study methods, and outcomes of significance to the review questions and objectives and can be found in Section 6.2. Data extraction templates were piloted on manuscripts for each of the included study designs and amendments were made prior to data extraction.

### 5.5 Quality appraisal

#### 5.5.1 Quality of the SRs and map/overview of reviews

Eligible SRs and maps/overviews of SRs were critically appraised using the JBI critical appraisal checklist for systematic reviews and research syntheses (Aromataris et al. 2015).. When a study or review met a criterion for inclusion, a score of one was given. Where a particular point for inclusion was regarded as not met, “unclear”, or “not applicable”, a score of zero was given. Overall critical appraisal scores were presented. The results of critical appraisal were reported in narrative form as well as in Table 16. Methodological quality assessment was conducted by one reviewer and checked by another. All SRs and maps/overviews of SRs, regardless of the results of their methodological quality, underwent data extraction and synthesis.

Alternative appraisal tools that can be used for assessing the quality of SRs, evidence maps and overviews of reviews include the AMSTAR-2 (Shea et al. 2017). While in this rapid review, the JBI critical appraisal checklist for systematic reviews and research syntheses (Aromataris et al. 2015) was selected due to its ability to be completed more swiftly than AMSTAR-2, four of the JBI quality checklist questions could be matched to the domains deemed critical in the AMSTAR-2 which were considered relevant to this review. A fifth AMSTAR-2 critical domain also assessed by the JBI checklist, the impact of publication bias, was not considered as relevant to this review because the majority of studies within the included SRs were case reports.

As a result, the JBI domains considered critical after the mapping include the following:

Q3: Was the search strategy appropriate?
Q4: Were the sources and resources used to search for studies adequate?
Q5: Were the criteria for appraising studies appropriate?
Q8: Were the methods used to combine studies appropriate?

Each review was then assessed based on the answers provided to the four critical domains as well as the remaining, non-critical, domains, and an overall rating of quality for each review was generated as detailed below.

- High [++]: No or one non-critical weakness. The systematic review provides an accurate and comprehensive summary of the results of the available studies that address the question of interest.
- Moderate [+]: More than one non-critical weakness^2^. The systematic review has more than one weakness but no critical flaws. It may provide an accurate summary of the results of the available studies that were included in the review.
- Low [-]: One critical flaw with or without non-critical weaknesses. The review has a critical flaw and may not provide an accurate and comprehensive summary of the available studies that address the question of interest.
- Critically low [--]: More than one critical flaw with or without non-critical weaknesses. The review has more than one critical flaw and should not be relied on to provide an accurate and comprehensive summary of the available studies.

#### 5.5.2 Quality of the case reports

Case reports were critically appraised using the JBI checklist for case reports (Moola et al. 2020). When a study or review met a criterion for inclusion, a score of one was given. Where a particular point for inclusion was regarded as not met, “unclear”, or “not applicable”, a score of zero was given. Overall critical appraisal scores were presented. The results of critical appraisal were reported in narrative form as well as in Table 17. Methodological quality assessment was conducted by one reviewer and checked by another. No formal process was used to rate the overall quality of the case reports, but where this was reported within individual reviews, it was extracted.

### 5.6 Synthesis

The data were reported narratively as a series of thematic summaries (Thomas et al. 2017) and was structured around the type of special procedure and adverse events.

For question 2 (body piercing), the overlap of primary research studies included in the SRs was checked and reported narratively and using tables and figures. For question 3 (acupuncture), the overlap of SRs in the maps/overviews of SRs was checked manually and reported narratively. The term overlap is used when multiple SRs on a topic include the same primary research studies (Lunny et al. 2021). Overlap can lead to issues with precision, as duplicate studies could cause an overestimation of sample sizes and number of events. Hence, it is important to determine how much overlap exists between included SRs (Lunny et al. 2021).

As a first stage for determining overlap, primary studies included in the four body piercing SRs were listed in MS Excel to see which ones were included in more than one review. Listing was assisted by the use of Web of Science, as it enabled the download of SR reference lists as an MS Excel file. To ensure the accuracy of the Web of Science generated reference lists, these were compared to data extraction and summary tables of SRs, and the included primary research studies were selected. In cases where the Web of Sciences did not identify the included primary research studies, these were manually added.

After studies were tabulated for each SR, a final overlap table was created where only unique studies were listed. Unique studies were coded depending on which review they were included in. Based on the format developed by (Bougioukas et al. 2023), the number 1 was allocated to studies when they appeared in a review, and the number 0 if they were not included. The final overlap table is presented in Appendix 5 (Table 19). Following coding, the overlap tables were imported into R Studio software to calculate the corrected covered area (CCA; Pieper et al. 2014). The corrected covered area is used to determine the degree of overlap between SRs. Using this approach, less than 5% CCA is a slight overlap, 6-10% CCA is a moderate overlap, 11-15% CCA is a high overlap and >15% CCA is a very high overlap (Pieper et al. 2014). In R Studio, the cca R package was used for calculation both across the four SRs and pairwise and to create a heatmap depicting pairwise CCA (Bougioukas et al. 2022).

The corrected covered area across the four SRs was 8.8%, indicating a moderate overlap. There were 136 unique primary studies out of 172 reported across the four SRs. While the narrative of the four SRs indicated 174 included primary studies, in one review (Sindoni et al. 2022), references for two studies could not be found. Therefore, these two could not be compared to studies reported in the other SRs, leading to only 172 studies used for the CCA calculation.

Regarding pairwise comparison between SRs, Acuña-Chavez et al. (2022) had no overlap with Hennequin-Hoenderdos et al. (2016) and Passos et al. (2022). This is due to Acuña-Chavez et al. (2022) focusing on nipple piercings specifically, while the other two focused on oral piercings. The highest degree of overlap (CCA=25.5%) was between Hennequin-Hoenderdos et al. (2016) and Passos et al. (2022), as apart from one study, Passos et al. (2022) contained all studies from Hennequin-Hoenderdos et al. (2022). Sindoni et al. (2022) focused on all piercing sites; hence, it overlapped with all the other three SRs. The heatmap depicting further details and comparisons is presented in Figure 1 below.

**Figure 1.**
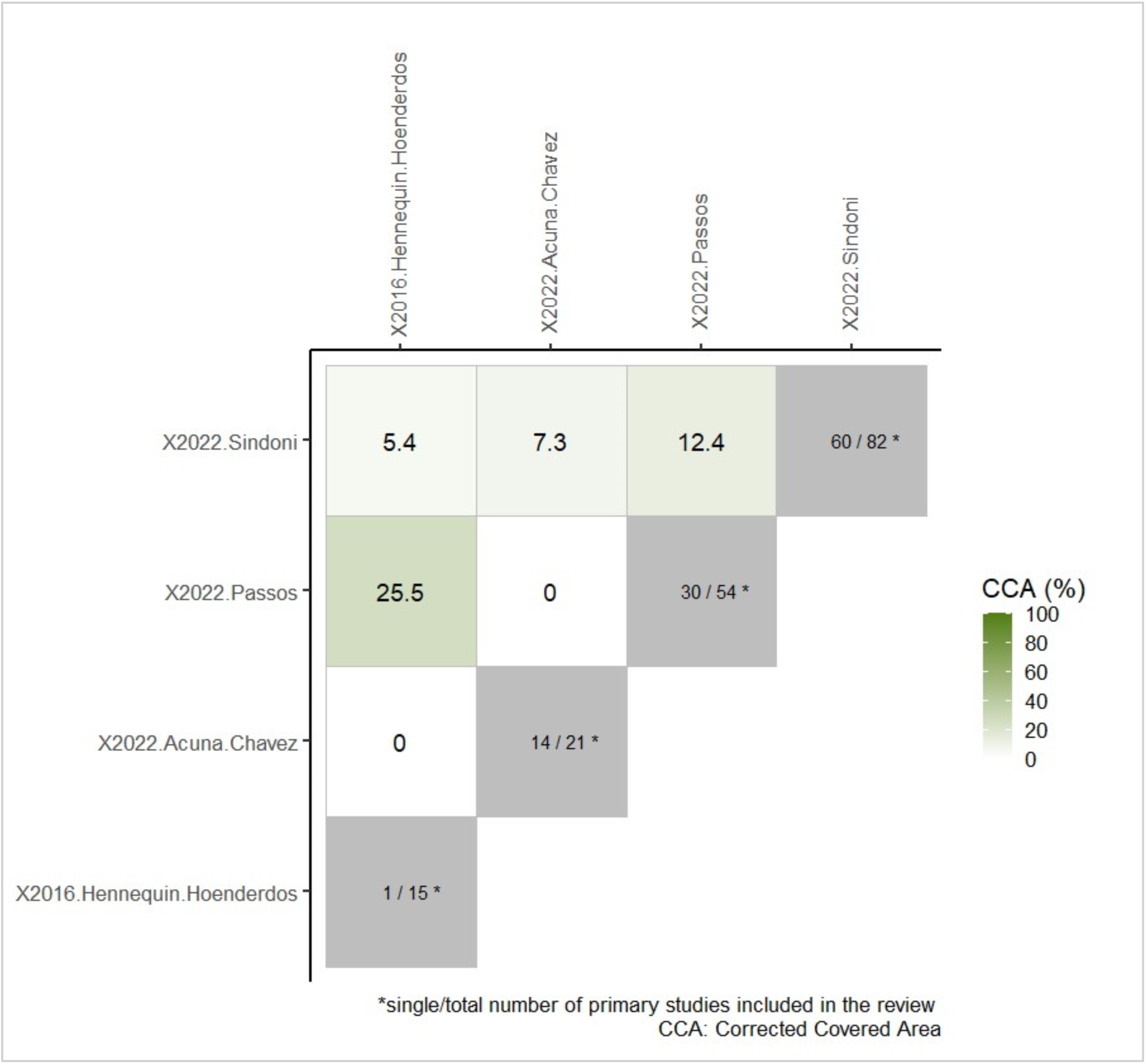
Heatmap depicting pairwise corrected covered area comparison of body piercing systematic reviews. As for the map and overview of SRs of acupuncture reported here (Xu et al. 2023, Yang et al. 2015), the overlap of SRs included in them was checked manually due to the fact that one of the overviews of SRs (Yang et al. 2015) only included six relevant SRs. One of these SRs was also reported in Xu et al. (2023). Determining the overlap between the primary studies included in the SRs reported in the overviews of SRs was not possible due to the volume of SRs and the time constraints of this rapid review.

### 5.7 Assessment of body of evidence

No formal process was used to rate the overall confidence in the evidence, but where it was reported within individual reviews, this was extracted. The GRADE (Grading of Recommendations, Assessment, Development, and Evaluation) approach assigns a certainty value to provide stakeholders with a measure of confidence in the quality of the evidence (Guyatt et al. 2011). As GRADE assessment is usually conducted using outcome effect estimates from primary studies or from a meta-analysis in rapid reviews where SRs are included, the Cochrane Rapid Review Methods Group recommends using existing GRADE from these reviews (Gartlehner et al. 2023). In this rapid review, of the five included SRs, an evidence map, and an overview of reviews, only one (Passos et al. 2022) used the GRADE approach. Due to the rapid review timeframe and the types of evidence included in the SRs with no GRADE, it was not possible for us to establish certainty of evidence based on the GRADE approach.

## 6. EVIDENCE

### 6.1 Search results and study selection

Searches for SRs of physical health risks associated with tattooing and semi-permanent make-up identified two relevant SRs, one of which focused on tattooing and the other on micropigmentation. The SR of the risks of micropigmentation had not identified any relevant literature, so separate searches for primary research into the risks of semi-permanent make-up were conducted. They resulted in 31 included studies. Searches for the other types of special procedures identified four SRs of body piercing, one evidence map, four overviews of SRs of acupuncture, and one primary study of electrolysis. Details of the methodology, including the search and selection process, are provided in Section 5 of this report. The flow of studies through the study selection process is reported in Table 9.

**Table 9.**
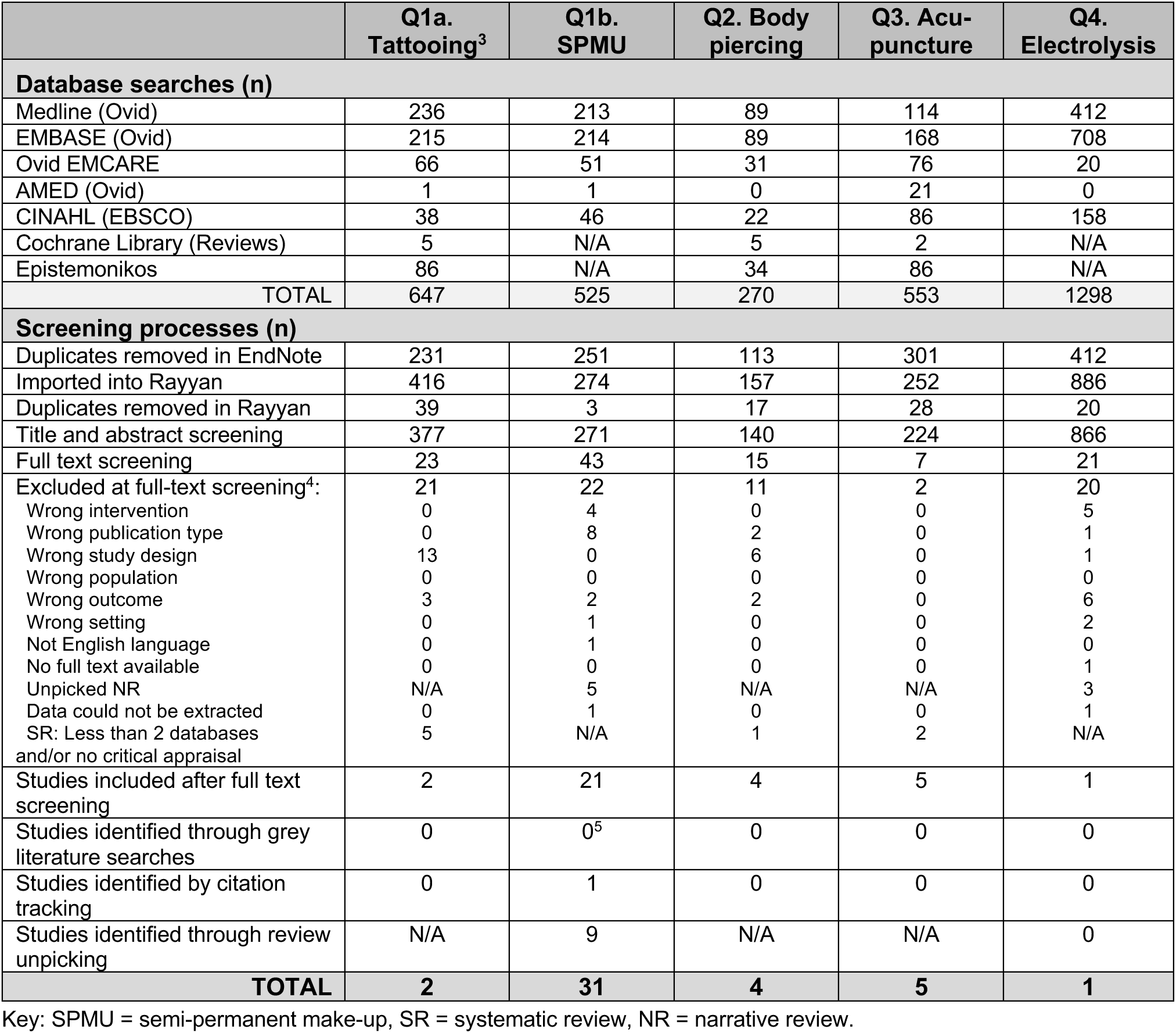
Flow of studies through the study selection process.

### 6.2 Data extraction

Separate tables are presented for the data extraction for each special procedure.

**Table 10.** Summary of included systematic reviews – Tattooing.

| Citation | Review details<br>Participants<br>Outcomes of interest | Characteristics of<br>included primary studies | Details of search<br>Quality of the<br>primary research<br>Quality of the review | Findings |
| --- | --- | --- | --- | --- |
| Sindoni et al. 2021<br>Adverse effects related to tattoos in the community setting: a systematic review.<br>https://doi.org/10.1136/jech-2021-216874 | <u>Aim</u><br>To assess the literature evidence about decorative tattoos complications, considering both infective and non-infective risks<br><u>Special procedure/s</u><br>Tattoos (made by injecting one or more inks into the skin by the use of needles)<br><u>Punctured site</u><br>Microbiological (n=10):<br>Not reported (n=3)<br>Multiple (n=2)<br>Leg (n=3)<br>Arm (n=2)<br>Non-microbiological (n=37)<br>Not reported (n=7)<br>Multiple (n=8)<br>Torso (n=7)<br>Arm (n=7)<br>Leg (n=3)<br>Face (n=3)<br>Buttock (n=1)<br>Penis (n=1) | <u>Number of included studies</u><br>55<br><u>Study designs</u><br>Case report (n=32)<br>Case series (n=13)<br>Cross-sectional (n=7)<br>Cohort study (n=2)<br>Retrospective study (n=1)<br><u>Date period covered by primary studies</u><br>1991-2020<br><u>Time frame for reporting/experiencing adverse effects</u><br>Microbiological (reported in 4/10)<br>6 & 10 days (2 cases, 1 study)<br>3 weeks (n=1)<br>2 months (n=2)<br>Non-microbiological (reported in 19/37):<br>2 weeks (n=2)<br>1-2 months (n=3)<br>3-6 months (n=2)<br>3,6 and 7 months (3 cases)<br>1 study<br>1-2 years (n=9) | <u>Date restrictions</u><br>Inception until April 2020<br><u>Databases searched</u><br>PubMed<br>Scopus<br>Web of Science<br><u>Language restrictions</u><br>English<br><u>Appraisal scale</u><br>JBI checklist for case reports<br>National Institute of Health Quality Assessment Tools (cohort studies, case series, cross sectional studies – Newcastle-Ottawa Quality Assessment Tool for Cohort Studies<br>Jadad Scale (RCTs)<br><u>Appraisal rating</u><br>Good quality (n=40)<br>Fair quality (n=5)<br>Poor quality (n=2)<br>CA of another 8 studies is not reported | <u>Microbiological adverse effects</u><br>10 studies described the following<br>60 cases of non-tuberculous *Mycobacteria* skin infections across 2 case series (age range 19-54) were also found from tattoos performed with diluted ink<br>14 cases of *Mycobacterium chelonae* from 17 tissue specimens + 4 cases of cutaneous Mycobacterial infection (*Mycobacterium chelonae* infection) among 134 tattooed people. premixed ink contaminated before distribution was hypothesised as a risk factor<br>2 cases of life-threatening cellulitis<br>1 case of cutaneous diphtheria across 2 studies<br>1 case of Staphylococcal scalded skin syndrome caused by tattoos or by illegally imported tattoo inks<br>1 case of HCV infection in a man with around 750 tattoos, some performed by himself<br>1 case warts (a case series found that warts were seven times more likely to be localised within black ink of tattoos when compared with coloured ink). Caused by tattoos or by illegally imported tattoo inks<br>Outbreaks were associated with poor infection control practices at tattoo studios and contamination of grey wash ink at point of manufacture<br><u>Non-microbiological adverse effects</u><br>There were 27 case reports, 8 case series, 1 descriptive study and 1 cohort study that described non microbiological effects. Thirty-three studies had a good quality, four showed a fair quality<br>From the descriptive survey<br>Skin problems (67.5%)<br>Systemic reactions (6.6%)<br>From the cohort study |
|  | Both microbiological and non-biological (n=8)<br>Not reported (n=8)<br><u>Practitioner</u><br>Not reported<br><u>Participants</u><br>Mostly adults (up to ~80 years)<br>3 studies included children under 18<br><u>Sex</u><br>Case Reports (32)<br>Males (16)<br>Females (14)<br>Mix (2)<br>Remaining study types (23):<br>Males (2)<br>Females (1)<br>Mix (7)<br>Not reported (13)<br><u>Outcomes of interest</u><br>Microbiological and non-microbiological adverse effects as a result of tattoos | Several years (n=1)<br>8 years (n=1)<br>Over 10 years (n=1)<br><u>Countries</u><br>Not reported<br><u>Settings</u><br>Not reported | <u>Microbiological adverse effects</u><br>Good quality (n=7)<br>Poor quality (n=2)<br>Fair quality (n=1)<br><u>Microbiological adverse effects</u><br>Good quality (n=33)<br>Fair quality (n=4)<br><u>Review appraisal score</u><br>Scored 8 out of 11 on the JBI critical appraisal checklist for systematic reviews and research syntheses. | • Short tear film break-up time<br>• Corneal erosion and meibomian gland loss<br>• Increased tear film instability and exacerbated signs and symptoms of ocular surface disease<br>From the case reports<br>• Scar, intractable nausea and vomiting<br>• Dermatofibrosarcoma protuberans<br>• Cutaneous and pulmonary sarcoidosis<br>• Extensive deposition of tattoo ink in lymph nodes<br>• Systemic anaphylaxis<br>• Pseudoepitheliomatous hyperplasia<br>• Blistering of the skin over therapeutic application on tattoo site<br>• Systemic anaphylaxis<br>• Pyoderma gangrenosum<br>• Allergic contact dermatitis<br>• Hemangioma<br>• Epithelioid osteoblastoma<br>• Lichenoid and inflammatory reactions to chemically reactive substances of the tattoo dye<br>• Skin graft and reconstructive surgery not possible due to multiple tattoos in donor sites<br>• Sarcoidosis and sarcoid uveitis<br>• Granulomatous dermatitis<br>• Bleeding and priapism<br>• Burning pain and skin reaction during Magnetic Resonance Imaging execution<br>• Inflammatory reactions<br>From case series<br>Skin capacitance higher in tattooed skin compared to non-tattooed skin.<br>• Allergic tattoo reactions<br>• Melanoma<br>• Pseudoepitheliomatous hyperplasia<br>• Lichenoid reaction<br>• Keratoacanthoma<br>• Squamous cell carcinoma<br>• Basal cell carcinoma<br>• Systemic sarcoidosis<br>• Uveitis<br>• Allergic contact dermatitis to a red-brown dye |
|  |  |  |  | • Dermatofibrosarcoma protuberans<br>• Dermatofibroma<br>• Tattoo pigment in lymph nodes mimicking nodal metastasis in melanoma<br>• Granulomatous infiltrate<br>• Nodular dermal lymphohistiocytic infiltration<br>• Granulomatous dermatitis<br>• Leiomyosarcoma, follicular, nonresponse to laser treatment due to overrepresentation of titanium<br><u>Studies evaluating both microbiological and non-microbiological (8 studies)</u><br>1. Infections<br>2. Allergic reactions (37%), papulo-nodular reactions (13%), bacterial infections (11%), Sarcoidosis (5%) and photosensitivity, pain syndrome and lymphopathy<br>3. Unusual bleeding, local skin irritation, local infection, dye allergy, swollen glands near to the site<br>4. Oedema, pruritus, allergy, local infection and Koebner phenomenon<br>5. Skin irritations (short term redness, dry skin or tenderness), Hepatitis<br>6. Infection, pruritus and pain.<br>7. Pain, swelling, dermatitis, eczema, itching, skin thickening, allergic reactions and also suppuration, bleeding, dizziness, headache, scabs and fever<br>8. Tattoo-related infections followed by swelling, pain and other |

Key: CA: critical appraisal; HCV: Hepatitis C Virus.

**Table 11.**
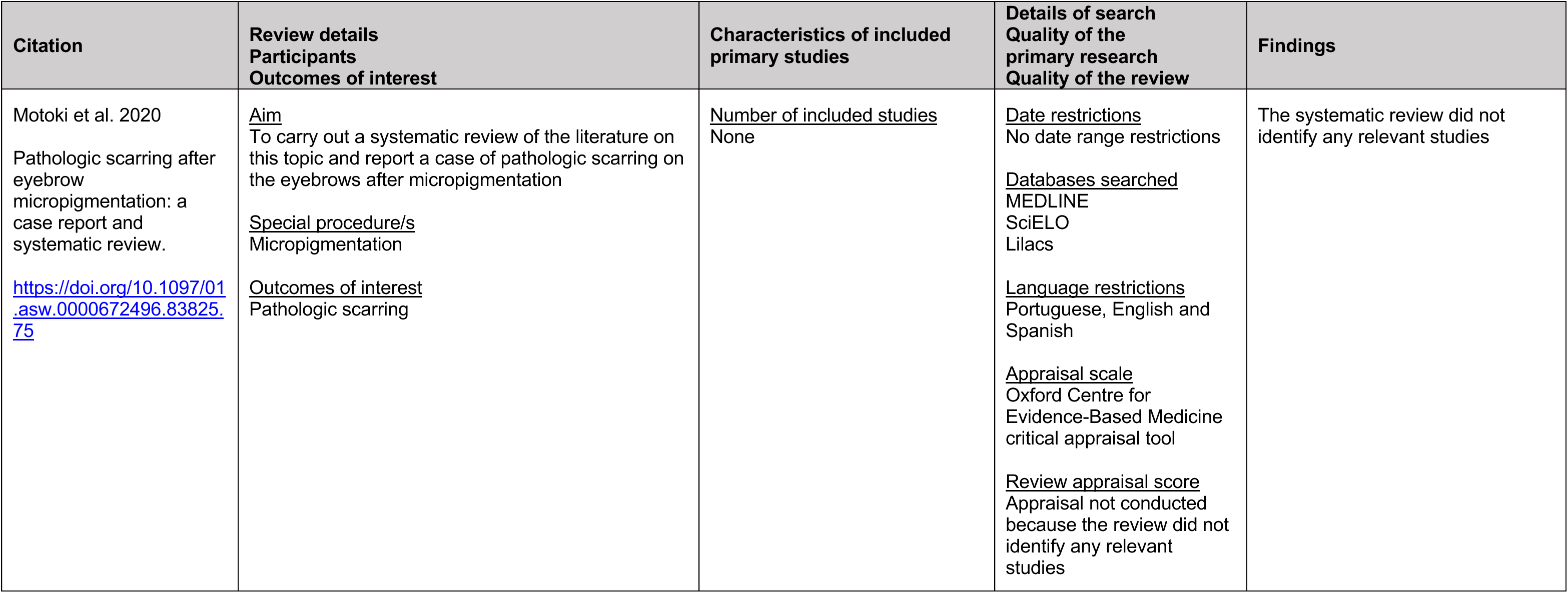
Summary of included systematic reviews – Semi-permanent make-up.

**Table 12.** Summary of included primary studies – Semi-permanent make-up.

| Citation | Study details<br>Quality ratings | Participants<br>Setting<br>Time frame for<br>experiencing/ reporting<br>adverse effect | Findings |
| --- | --- | --- | --- |
| Abtahi-Naeini 2019<br>Multiple pigmented macules as a sequel of cosmetic lip micro-pigmentation new clinical presentation of tattoo reactions<br>https://doi.org/10.4103/npmj.npmj_88_19 | <u>Study design</u><br>Case report<br><u>Outcome/s of interest</u><br>Lichenoid tattoo reactions<br><u>Comparator intervention or control</u><br>NA<br><u>Data collection methods</u><br>NA<br><u>Country</u><br>Iran<br><u>Quality rating</u><br>Scored 7 out of 8 on the JBI critical appraisal checklist for case reports | <u>Sample size</u><br>1<br><u>Participants</u><br>35-year-old woman<br><u>Setting</u><br>Commercial<br><u>Practitioner</u><br>Nonmedical professional<br><u>Puncture site</u><br>Lips<br><u>Time frame</u><br>5 weeks | A 35-year-old woman received permanent make-up (red-pink) of lips and lip lines by micro-pigmentation and developed hyperpigmentation, associated with mild pruritus, occurred on her upper lip line, 5 weeks after a single session of tattooing. The histopathological sections of a biopsy from pigmented lip macule were notable lichenoid lymphocytic infiltrations. Mild acanthosis, basal vacuolar degeneration and prominent band-like infiltration were notable. A diagnosis of lichenoid tattoo reactions following lip tattooing was made. |
| Akoh 2021<br>A rare case of microblading-induced preseptal cellulitis<br>https://doi.org/10.1016/j.jdc.2021.08.014 | <u>Study design</u><br>Case report<br><u>Outcome/s of interest</u><br>Preseptal cellulitis<br><u>Comparator intervention or control</u><br>NA<br><u>Data collection methods</u><br>NA | <u>Sample size</u><br>1<br><u>Participants</u><br>39-year-old woman<br><u>Setting</u><br>Commercial<br><u>Practitioner</u><br>Aesthetician<br><u>Puncture site</u> | A 39-year-old woman with sickle cell disease presented with a significant periocular facial eruption (pustules in the malar area of the right cheek, which progressed to bilateral periorbital edema and pustules draining purulent exudate) after undergoing microblading 3 weeks prior. She had no history of allergies, photosensitivity, or similar skin eruptions. A few days later, she presented again with bilateral periorbital hyperpigmentation, edema, erythema, crusting, and pustules with involvement of the malar area of the cheeks and was disoriented, afebrile, and tachycardic. Preseptal cellulitis secondary to microblading was diagnosed. |
|  | <u>Country</u><br>USA<br><u>Quality rating</u><br>Scored 8 out of 8 on the JBI critical appraisal checklist for case reports | <u>Eyebrows</u><br><u>Time frame</u><br>3 weeks |  |
| Bashinskaya 2022<br>Permanent make-up procedure heralds the development of systemic sarcoidosis<br>https://doi.org/10.7759/cureus.30918 | <u>Study design</u><br>Case report<br><u>Outcome/s of interest</u><br>Systemic sarcoidosis<br>Cutaneous sarcoidosis, including non-caseating epithelioid granulomas<br><u>Comparator intervention or control</u><br>NA<br><u>Data collection methods</u><br>NA<br><u>Country</u><br>USA<br><u>Quality rating</u><br>Scored 5 out of 8 on the JBI critical appraisal checklist for case reports | <u>Sample size</u><br>1<br><u>Participants</u><br>48-year-old Hispanic woman<br><u>Setting</u><br>Commercial<br><u>Practitioner</u><br>NR<br><u>Puncture site</u><br><u>Eyebrows</u><br><u>Time frame</u><br>NR | A report on a 48-year-old Hispanic female with a new onset of scar sarcoidosis that progressed to a systemic condition. Erythematous maculopapular eruptions arose on her left eyebrow area at the sites of scars from cosmetic tattooing, prior to the small airway disease exacerbation. Histopathologic examination revealed typical findings of cutaneous sarcoidosis, including non-caseating epithelioid granulomas. |
| Bombonato 2015<br>Orange color a dermoscopic clue for the diagnosis of granulomatous skin diseases<br>https://dx.doi.org/10.1016/j.jaad.2014.07.059 | <u>Study design</u><br>Case report<br><u>Outcome/s of interest</u><br>Chronic granulomatous inflammation<br><u>Comparator intervention or control</u><br>NA | <u>Sample size</u><br>1<br><u>Participants</u><br>38-year-old woman<br><u>Setting</u><br>Commercial<br><u>Practitioner</u><br>NR | A report of 38-year-old woman who presented with asymptomatic coalescing hard yellowish red papules located on her eyebrows overlying a cosmetic tattoo. Histologic evaluation revealed a chronic granulomatous inflammation without necrosis in the superficial and deep dermis. In the dermis, giant cells and foreign amorphous material and black pigments were detected, which suggested the diagnosis of granulomatous reaction on the top of a cosmetic tattoo. |
|  | <u>Data collection methods</u><br>NA<br><u>Country</u><br>Italy<br>Quality rating<br>Scored 4 out of 8 on the JBI critical appraisal checklist for case reports | <u>Puncture site</u><br><u>Eyebrows</u><br><u>Time frame</u><br>NR |  |
| Castaño-Fernández 2023<br>Microblading-transmitted Monkeypox (mpox) infection fomites matter<br>https://doi.org/10.1093/bjd/ljad063 | <u>Study design</u><br>Case report<br><u>Outcome/s of interest</u><br>Monkeypox (mpox) infection<br><u>Comparator intervention or control</u><br>NA<br><u>Data collection methods</u><br>Physical examination and Lab Test<br><u>Country</u><br>Spain<br><u>Quality rating</u><br>Scored 5 out of 8 on the JBI critical appraisal checklist for case reports | <u>Sample size</u><br>1<br><u>Participants</u><br>47-year-old woman<br><u>Setting</u><br>Commercial<br><u>Practitioner</u><br>NR<br><u>Puncture site</u><br><u>Eyebrows</u><br><u>Time frame</u><br>10 days | A 47-year-old woman with no relevant medical history presented with a 10-day facial skin eruption, after undergoing eyebrow microblading 4 days before the lesions started. Physical examination revealed a plaque with central necrotic ulceration on the medial margin of her left eyebrow. The mpox virus polymerase chain reaction test came back positive while other tests were negative. Mpox is transmitted by direct skin contact or by infected fomites. |
| Ebrahimiadib 2021<br>Intermediate uveitis associated with tattooing of eyebrows as a manifestation of systemic sarcoidosis report of two cases<br>https://doi.org/10.1080/09273948.2019.1699581 | <u>Study design</u><br>Case report<br><u>Outcome/s of interest</u><br>Systematic sarcoidosis<br>Non-caseating granulomas uveitis<br><u>Comparator intervention or control</u> | <u>Sample size</u><br>2<br><u>Participants</u><br>46-year-old woman<br>49-year-old woman<br><u>Setting</u><br>Commercial | Case report describing two women presented with tattoo-associated uveitis as the first manifestations of systemic sarcoidosis. They developed intermediate uveitis shortly after skin inflammation several months after permanent make-up tattooing of eyebrows. Both had scaling and red papules on tattoo sites and one had swelling. Lung involvement, high angiotensin-converting enzyme levels, and negative PPD were present. Biopsy confirmed the presence of non-caseating granulomas consistent with systemic sarcoidosis in both. Both had non-granulomatous intermediate uveitis of moderate intensity. Skin granuloma formation was |
|  | NA<br><u>Data collection methods</u><br>NA<br><u>Country</u><br>Iran<br><u>Quality rating</u><br>Scored 7 out of 8 on the JBI critical appraisal checklist for case reports | <u>Practitioner</u><br>NR<br><u>Puncture site</u><br>Eyebrows<br><u>Time frame</u><br>NR | diffused over the area of the tattoo in one patient and localized in the other. |
| Gilhooley 2020<br>Photoinduced granulomatous reaction of cosmetically tattooed lips<br>https://doi.org/10.1111/jocd.13664 | <u>Study design</u><br>Case report<br><u>Outcome/s of interest</u><br>Non-caseating granulomas<br><u>Comparator intervention or control</u><br>NA<br><u>Data collection methods</u><br>NA<br><u>Country</u><br>Ireland<br><u>Quality rating</u><br>Scored 8 out of 8 on the JBI critical appraisal checklist for case reports | <u>Sample size</u><br>1<br><u>Participants</u><br>33-year-old woman<br><u>Setting</u><br>Commercial<br><u>Practitioner</u><br>NR<br><u>Puncture site</u><br>Eyebrows and lips<br><u>Time frame</u><br>18 months after additional ultraviolet sunbed exposure | A 33-year-old woman presented with erythematous, edematous, and inflamed lips and reported an acute onset of symptoms following ultraviolet radiation exposure on a sunbed 6 months prior. She had cosmetically tattooed her lips and eyebrows 18 months prior. It was the first exposure to sunbed ultraviolet radiation after tattooing.<br>Viral and bacterial swabs, serological investigations, fecal calprotectin test and chest X-ray did not identify any findings. On histopathology, aggregated epithelioid histiocytes formed discrete noncaseating granulomas within the dermis. In the superficial dermis, histiocytes displayed pigment. Under polarized light, the pigment was consistent with a foreign body. These findings were consistent with a granulomatous reaction to tattoo pigment. |
| Goldberg 2018<br>Inadvertent corneal pigmentation following cosmetic blepharopigmentation<br>https://doi.org/10.1016/j.ajoc.2018.09.002 | <u>Study design</u><br>Case report<br><u>Outcome/s of interest</u><br>Conjunctival injection and pigmented stain on the eye | <u>Sample size</u><br>1<br><u>Participants</u><br>63-year-old woman<br><u>Setting</u> | A 63 year old woman presented with redness and irritation in her left eye for two days. A thick black pigmentation line of both upper and lower lash lines was evident. In the left eye there was conjunctival injection and on the nasal part of the cornea there was a large black pigmented stain. Three days prior to she had undergone permanent eyeliner tattooing on the upper and lower eyelids of both eyes and was not aware of any complications during |
|  | <u>Comparator intervention or control</u><br>NA<br><u>Data collection methods</u><br>NA<br><u>Country</u><br>Israel<br><u>Quality rating</u><br>Scored 8 out of 8 on the JBI critical appraisal checklist for case reports | Commercial<br><u>Practitioner</u><br>NR<br><u>Puncture site</u><br>Eyelids<br><u>Time frame</u><br>One day | the tattooing. On the following day she started feeling progressive irritation in the left eye. The pigment contained mainly carbon, oxygen and potassium, and small amounts of calcium, sulfur, chlorine and phosphorus. The presence of lead or copper was ruled out. |
| Hinojosa 2017<br>Verrucous eyebrows a cutaneous manifestation of a systemic disease<br>https://doi.org/10.1111/jdv.14246 | <u>Study design</u><br>Case report<br><u>Outcome/s of interest</u><br>Cutaneous sarcoidosis<br>Systematic sarcoidosis<br>Conjunctival injection<br><u>Comparator intervention or control</u><br>NA<br><u>Data collection methods</u><br>NA<br><u>Country</u><br>Unclear – Mexico or USA<br><u>Quality rating</u><br>Scored 8 out of 8 on the JBI critical appraisal checklist for case reports | <u>Sample size</u><br>1<br><u>Participants</u><br>52-year-old Hispanic woman<br><u>Setting</u><br>Commercial<br><u>Practitioner</u><br>NA<br><u>Puncture site</u><br>Eyebrows<br><u>Time frame</u><br>3 years | A 52-year-old Hispanic woman presented with a 5-month history of lesions on her eyebrows (brownish-pink verrucous plaques with slight crusting on both eyebrows). She also exhibited bilateral conjunctival injection without discharge. 3 years prior to presentation, she had undergone cosmetic eyebrow tattooing with an unspecified 'natural' substance in the same location. Microscopic examination revealed nodular collections of epithelioid histiocytes surrounded by a sparse infiltrate of lymphocytes throughout the dermis as well as the presence of an asteroid body. No foreign material was identified following the use of polarized light. A diagnosis of cutaneous sarcoidosis was made. Computed tomography of the chest confirmed the diagnosis of systemic sarcoidosis, with multiple subpleural nodules, multiple hilar nodules and prominent mediastinal and hilar lymph nodes. |
| Hirai 2022<br>Sarcoidosis and sarcoidal foreign body reaction after permanent eye makeup application Analysis by immunohistochemistry with commercially available | <u>Study design</u><br>Case report<br><u>Outcome/s of interest</u><br>Non-caseating Granulomas | <u>Sample size</u><br>2<br><u>Participants</u><br>75-year-old woman | Two cases of granulomas that occurred at permanent makeup application sites. In both cases, noncaseating epithelioid cell granulomas developed at the permanent eye makeup injection site. A 75-year-old woman presented with a rash on both eyebrows after permanent makeup procedure 1 year and 9 months prior. A skin |
| antibodies specific to Cutibacterium acnes and Mycobacteria<br>https://doi.org/10.1111/cup.14219 | Sarcoidosis and sarcoidal foreign body reaction<br><u>Comparator intervention or control</u><br>NA<br><u>Data collection methods</u><br>NA<br><u>Country</u><br>Japan<br><u>Quality rating</u><br>Scored 7 out of 8 on the JBI critical appraisal checklist for case reports | 46-year-old woman<br><u>Setting</u><br>Commercial<br><u>Practitioner</u><br>NR<br><u>Puncture site</u><br>Eyebrows<br><u>Time frame</u><br>21 months<br>10 years | biopsy of the eyebrow lesion revealed granulomatous inflammation in the upper dermis and subcutaneous tissue, but no necrotic changes. Most of the black brown granules of the permanent makeup were not present in the granulomas but were localized in the upper dermis. Sarcoidosis was diagnosed based on bilateral hilar lymphadenopathy, elevated ACE, abnormal accumulation in the hilar lymph nodes on gallium scintigraphy, and histopathological evidence in the skin biopsy specimen.<br>A 46-year-old woman presented with nodules on both eyebrows after a permanent makeup procedure 10 years prior. A skin biopsy of the eyebrow lesion revealed noncaseating epithelioid cell granulomas with few lymphocytes. Most of the black-brown granules of the permanent makeup were predominantly distributed outside of the granuloma, some of the black-brown granules were phagocytized in the granulomas. A diagnosis of sarcoidal foreign-body reaction was made. |
| Huisman 2019<br>Granulomatous tattoo reactions in permanent makeup of the eyebrows<br>https://doi.org/10.1111/jocd.12540 | <u>Study design</u><br>Case report<br><u>Outcome/s of interest</u><br>Granulomatous inflammation (5 cases)<br>Systemic sarcoidosis (1 case)<br><u>Comparator intervention or control</u><br>NA<br><u>Data collection methods</u><br>NA<br><u>Country</u><br>The Netherlands<br><u>Quality rating</u><br>Scored 7 out of 8 on the JBI critical appraisal checklist for case reports | <u>Sample size</u><br>5<br><u>Participants</u><br>All female, aged 33-57<br><u>Setting</u><br>Commercial<br><u>Practitioner</u><br>NR<br><u>Puncture site</u><br>Eyebrows (5 cases)<br><u>Time frame</u><br>4 to 15 years | The clinical presentation in the 5 patients includes elevated and frequently yellowish plaques, sharply demarcated around the tattooed skin. All patients got permanent makeup of their eyebrows several times during the last 4-15 years. The time between the last placement of the cosmetic tattoo and the onset of complaints varied between 1 and 18 months. The dyes used were in the red, brown, and black spectrum. No triggering factors could be identified, and there was no history of systemic complaints. Histopathological evaluation revealed granulomatous inflammation in all patients and additional stains were negative for fungi and acid-resistant mycobacteria. In patient 1, an elevated angiotensin-converting enzyme level of 126 (range 20-70 U/L) was found. In the same patient, high-resolution computed tomography revealed mediastinal lymphadenopathy, concluding systemic sarcoidosis. In patient 4, a chest X-ray revealed hilar adenopathy and dubious nodules. In the other patients, we did not find any signs of systemic sarcoidosis. Patients were treated with topical or intralesional corticosteroids, resulting in almost complete resolution of the skin manifestation. |
| Ibraheim 2023<br>Microblading-induced granulomatous reaction case report and review of the literature<br>https://doi.org/10.1097/DAD.0000000000002449 | <u>Study design</u><br>Case report<br><u>Outcome/s of interest</u><br>Foreign body granulomatous reaction<br>Cutaneous sarcoidosis<br>Contact dermatitis<br>Granulomatous dermatitis<br><u>Comparator intervention or control</u><br>NA<br><u>Data collection methods</u><br>NA<br><u>Country</u><br>Canada<br><u>Quality rating</u><br>Scored 7 out of 8 on the JBI critical appraisal checklist for case reports | <u>Sample size</u><br>1<br><u>Participants</u><br>25-year-old woman<br><u>Setting</u><br>Commercial<br><u>Practitioner</u><br>NR<br><u>Punctures site</u><br>Eyebrows<br><u>Time frame</u><br>7 months | A 25-year-old woman with no medical problems presented with a 1-month history of itchy, raised plaques affecting the eyebrows. She had previously undergone 5 microblading treatments, with the final one 7 months before the plaques appeared. The differential diagnosis included foreign-body granulomatous reaction, cutaneous sarcoidosis, and contact dermatitis. Punch biopsy revealed a dense, granulomatous dermatitis with associated brisk lymphoid infiltrate and pigment deposition. |
| Iwayama 2017<br>Granulomatous reaction to permanent eyebrow makeup successfully treated with topical steroids in combination with topical tacrolimus<br>https://doi.org/10.1684/ejd.2017.3106 | <u>Study design</u><br>Case report<br><u>Outcome/s of interest</u><br>Granulomatous reaction<br><u>Comparator intervention or control</u><br>NA<br><u>Data collection methods</u><br>NA<br><u>Country</u><br>Japan<br><u>Quality rating</u> | <u>Sample size</u><br>1<br><u>Participants</u><br>32-year-old woman<br><u>Setting</u><br>Commercial<br><u>Practitioner</u><br>NR<br><u>Puncture site</u><br>Eyebrows<br><u>Time frame</u><br>4 years | A 32-year-old Japanese woman presented with papules on her right eyebrow, 4 years after receiving permanent eyebrow makeup. Based on clinical and histopathological findings, the skin lesions were diagnosed as a granulomatous reaction to the colourants. |
|  | Scored 7 out of 8 on the JBI critical appraisal checklist for case reports |  |  |
| Lahouel 2022<br>Allergic contact dermatitis post-microblading eyebrow tattooing a manifestation of pre-existing nickel sensitization<br>https://doi.org/10.1111/jocd.14920 | <u>Study design</u><br>Case report<br><u>Outcome/s of interest</u><br>Allergic contact dermatitis (nickel induced)<br><u>Comparator intervention or control</u><br>NA<br><u>Data collection methods</u><br>NA<br><u>Country</u><br>Tunisia<br><u>Quality rating</u><br>Scored 7 out of 8 on the JBI critical appraisal checklist for case reports | <u>Sample size</u><br>1<br><u>Participants</u><br>50-year-old woman<br><u>Setting</u><br>Commercial<br><u>Practitioner</u><br>NR<br><u>Puncture site</u><br>Eyebrows<br><u>Time frame</u><br>2 days | A 50-year-old woman with a history of intolerance to fake jewellery presented with bilateral, symmetrical, itchy, erythematous lesions on the eyebrows, which appeared 2 days after the application of microblading to her eyebrows. The diagnosis was nickel-induced allergic contact dermatitis post-microblading eyebrow tattooing performed with nickel containing needles. |
| Leight-Dunn 2022<br>Delayed granulomatous reaction after eyebrow microblading<br>http://dx.doi.org/10.1097/DSS.0000000000003236 | <u>Study design</u><br>Case report<br><u>Outcome/s of interest</u><br>Granulomatous reaction<br>Granulomatous dermatitis<br><u>Comparator intervention or control</u><br>NA<br><u>Data collection methods</u><br>Clinical examination<br><u>Country</u><br>NR, possible the US | <u>Sample size</u><br>1<br><u>Participants</u><br>29-year-old women<br><u>Setting</u><br>NR, possibly commercial<br><u>Practitioner</u><br>NR<br><u>Puncture site</u><br>Eyebrows<br><u>Time frame</u> | A case of a 29-year-old woman who presented with asymptomatic, yellow–orange, flat, firm 2 to 3-mm papules and plaques confined to the eyebrow margins, and received a diagnosis of a granulomatous tattoo reaction to iron oxide. The papules appeared spontaneously, 4 months after her seventh microblading treatment which took place over a 5-year period. A shave biopsy showed granulomatous dermatitis to foreign pigment. The ink ingredients contained iron oxide, glycerine, and isopropyl alcohol. The granulomas had persisted for 1 year by the time the patient chose to suspend treatment. |
|  | <u>Quality rating</u><br>Scored 7 out of 8 on the JBI critical appraisal checklist for case reports | 4 months after 7<sup>th</sup> microblading treatment over a 5-year period |  |
| Maier 2015<br>High-definition optical coherence tomography and reflectance confocal microscopy in the in vivo visualization of a reaction to permanent make-up<br>https://doi.org/10.1111/jdv.12402 | <u>Study design</u><br>Case report<br><u>Outcome/s of interest</u><br>Granulomatous foreign body reaction<br><u>Comparator intervention or control</u><br>NA<br><u>Data collection methods</u><br>NA<br><u>Country</u><br>Germany<br><u>Quality rating</u><br>Scored 7 out of 8 on the JBI critical appraisal checklist for case reports | <u>Sample size</u><br>1<br><u>Participants</u><br>40-year-old woman<br><u>Setting</u><br>Commercial<br><u>Practitioner</u><br>NR<br><u>Punctures site</u><br>Lips<br><u>Time frame</u><br>6 months | A 40-year-old woman presented with erythema, swelling, nodular induration and scaling of her lips, one year ago after receiving permanent make-up. Half a year after the procedure she experienced a strong swelling, scaling, and tenderness of her lips. Both in high-definition optical coherence tomography and in reflectance confocal microscopy subepidermal pigment and granulomatous changes could be visualized and correlated with the histopathological findings. Regression of the lesions in response to topical steroids and intralesional injections of steroids and 5-fluorouracil is reported. |
| Marcelino 2021<br>*Molluscum contagiosum* as a tattoo complication a case report and literature review<br>https://doi.org/10.5935/scd1984-8773.2021130008 | <u>Study design</u><br>Case report<br><u>Outcome/s of interest</u><br>*Molluscum contagiosum*<br><u>Comparator intervention or control</u><br>NA<br><u>Data collection methods</u><br>NA<br><u>Country</u><br>Brazil | <u>Sample size</u><br>1<br><u>Participants</u><br>64-year-old woman<br><u>Setting</u><br>Commercial (aesthetic clinic)<br><u>Practitioner</u><br>Nonmedical professional<br><u>Puncture site</u><br>Eyebrows | A 64-year-old woman presented with 1 mm to 4 mm papules, pearly and normochromic, on the tattoo area on the right eyebrow, one month after eyebrows micropigmentation. The anatomopathological examination revealed a hyperplastic region in the epidermis characterized by cell proliferation from the Malpighian body, forming piriform invaginations into the dermis. Also found progressive accumulation of amorphous and eosinophilic material, compatible with *Molluscum contagiosum*. |
|  | <u>Quality rating</u><br>Scored 7 out of 8 on the JBI critical appraisal checklist for case reports | <u>Time frame</u><br>1 month |  |
| Mirzaei 2017<br>Adalimumab-Responsive Refractory Sarcoidosis Following Multiple Eyebrow Tattoos: A Case Report<br>https://pubmed.ncbi.nlm.nih.gov/28638429/ | <u>Study design</u><br>Case report<br><u>Outcome/s of interest</u><br>Löfgren's syndrome, an acute form of sarcoidosis<br><u>Comparator intervention or control</u><br>NA<br><u>Data collection methods</u><br>NA<br><u>Country</u><br>Iran<br><u>Quality rating</u><br>Scored 8 out of 8 on the JBI critical appraisal checklist for case reports | <u>Sample size</u><br>1<br><u>Participants</u><br>47- year-old woman<br><u>Setting</u><br>Commercial<br><u>Practitioner</u><br>NR<br><u>Puncture site</u><br>Eyebrows<br><u>Time frame</u><br>4 months | A 47- year-old woman with no significant medical history presented with polyarthritis along with symptoms of erythema nodosum-like nodules and low-grade fever and distinct red papules above the eyebrows. She reported multiple tattooing over the eyebrows, with the last tattooing 4 months before symptoms manifested. Results of hematological and biochemical tests were normal and Immunologic tests were negative. CT of thorax was showed bilateral hilar adenopathy with reticulonodular lesions in lower lobes of the lung. The diagnosis of Löfgren's syndrome, an acute form of sarcoidosis was confirmed based on the presence of the triad of erythema nodosum, bilateral hilar lymphadenopathy, and polyarthritis. |
| Motoki 2020<br>Pathologic scarring after eyebrow micropigmentation a case report and systematic review<br>https://doi.org/10.1097/01.ASW.0000672496.83825.75 | <u>Study design</u><br>Case report<br><u>Outcome/s of interest</u><br>Dermatitis<br><u>Comparator intervention or control</u><br>NA<br><u>Data collection methods</u><br>NA<br><u>Country</u><br>Brazil | <u>Sample size</u><br>1<br><u>Participants</u><br>50-year-old woman<br><u>Setting</u><br>Commercial<br><u>Practitioner</u><br>Medical<br><u>Puncture site</u><br>Eyebrows<br><u>Time frame</u> | A report on a 50-year-old female aesthetician having nodules on her eyebrows approximately 2 months after an eyebrow micropigmentation procedure. She had undergone eyebrow micropigmentation annually for the last 4 years using a medium brown coloring agent. She reported constant pruritus, mild sharp pain, local hyperemia, and peeling. Recent micropigmentation procedures occurred 10 months and 2 months prior to her current presentation. A dermatologist diagnosed her eyebrow scarring as dermatitis. At the outpatient clinic during the initial physical examination, it was noted a fibroproliferative scar with significant vertical and horizontal growth originating in the micropigmentation area suggestive of hypertrophic scarring. |
|  | <u>Quality rating</u><br>Scored 7 out of 8 on the JBI critical appraisal checklist for case reports | Undergoing microblading annually for 10 years and 2 months prior to current presentation |  |
| Naeini 2017<br>Looking beyond the cosmetic tattoo lesion near the eyebrow Screening the lungs<br>https://doi.org/10.4103/0022-3859.201421 | <u>Study design</u><br>Case report<br><u>Outcome/s of interest</u><br>Pulmonary sarcoidosis<br>Cutaneous sarcoidal granulomas<br><u>Comparator intervention or control</u><br>NA<br><u>Data collection methods</u><br>NA<br><u>Country</u><br>Iran<br><u>Quality rating</u><br>Scored 7 out of 8 on the JBI critical appraisal checklist for case reports | <u>Sample size</u><br>1<br><u>Participants</u><br>45-year-old woman<br><u>Setting</u><br>Commercial<br><u>Practitioner</u><br>NR<br><u>Puncture site</u><br>Eyebrows<br><u>Time frame</u><br>NR | A 45-year-old woman with a 15-month history of lesions on her eyebrow tattoo. Her tattoos had red, scaly patches. A skin biopsy showed non necrotising granulomas with few lymphocytes around the granuloma and negative results on acid-fast Bacillus and fungal stains. Chest imaging revealed perihilar lymphadenopathy and few small nodules in both lungs, increased Interstitial markings and peribronchial cuffing. Blood tests revealed an abnormally high level of angiotensin-converting enzyme. A diagnosis of pulmonary sarcoidosis was supported by the presence of cutaneous sarcoidal granulomas. |
| Navarro-Triviño 2021<br>Cutaneous lymphoid hyperplasia caused by cosmetic lip tattoo successfully treated with tacrolimus 0.1% ointment<br>https://doi.org/10.1111/cod.13726 | <u>Study design</u><br>Case report<br><u>Outcome/s of interest</u><br>Cutaneous lymphoid hyperplasia<br><u>Comparator intervention or control</u><br>NA<br><u>Data collection methods</u><br>NA | <u>Sample size</u><br>1<br><u>Participants</u><br>52-year-old Caucasian woman<br><u>Setting</u><br>Commercial<br><u>Practitioner</u><br>NR<br><u>Puncture site</u> | A 52-year-old Caucasian woman with red papules and nodules on the lip vermilion, exclusively affecting the tattooed area after a micropigmentation tattoo lasting 1 year. She reported a tattoo micropigmentation 5 years earlier. The patient showed a positive patch test reaction to tattoo ink. A skin lip biopsy revealed cutaneous lymphoid hyperplasia. The presence of exogenous red pigment was observed. Staining demonstrated CD3 and CD4 lymphocytes. Cutaneous lymphoid hyperplasia caused by permanent makeup ink was diagnosed. |
|  | <u>Country</u><br>Spain<br><u>Quality rating</u><br>Scored 7 out of 8 on the JBI critical appraisal checklist for case reports | Eyebrows<br><u>Time frame</u><br>5 years |  |
| Nie 2022<br>Eyebrow tattoo-associated sarcoidosis a case report<br>https://doi.org/10.3389/fmed.2022.1009135 | <u>Study design</u><br>Case report<br><u>Outcome/s of interest</u><br>Sarcoidosis<br>Non-caseating granulomas<br><u>Comparator intervention or control</u><br>NA<br><u>Data collection methods</u><br>NA<br><u>Country</u><br>China<br><u>Quality rating</u><br>Scored 7 out of 8 on the JBI critical appraisal checklist for case reports | <u>Sample size</u><br>1<br><u>Participants</u><br>41-year-old woman<br><u>Setting</u><br>Unclear. The report states that the patient tattooed her eyebrows, so may be a home procedure.<br><u>Practitioner</u><br>NR<br><u>Puncture site</u><br>Eyebrows<br><u>Time frame</u><br>3 months | A 41-year-old Chinese woman presented with a slowly enlarged rash on the bilateral eyebrows and grouped reddish and yellow–brown infiltrative papules, some of which coalesced into plaques scattered over the tattoo. She had tattooed her eyebrows 3 months before the rash appeared. CT of the chest with contrast demonstrated a few diffuse reticulonodular opacities in the bilateral lower lobe, bilateral hilar lymph nodes enlargement, and in the posterior segment of the right upper lobe are some small ground glass nodules. Tattoo-associated sarcoidosis was diagnosed based on the clinical manifestation of yellow–brown plaque and histopathological findings of non-caseating granulomas at the tattoo site. |
| Sano 2022<br>Bilateral periorbital cellulitis after eyebrow microblading<br>https://doi.org/10.1016/j.visj.2021.101172 | <u>Study design</u><br>Case report (Visual Case Discussion)<br><u>Outcome/s of interest</u><br>Bilateral periorbital cellulitis<br><u>Comparator intervention or control</u><br>NA<br><u>Data collection methods</u><br>NA | <u>Sample size</u><br>1<br><u>Participants</u><br>49-year-old woman<br><u>Setting</u><br>Commercial<br><u>Practitioner</u><br>NR<br><u>Punctured site</u> | A 49-year-old presented with bilateral eye swelling, redness, and pain for two weeks and foul-smelling discharge from both eyes for on the day of presentation, after eyebrow microblading. Examination of the patient shows notable bilateral periorbital erythema, swelling, and crusting of the skin around her eyebrows. Given her severe penicillin allergy, she was treated with IV fluids, IV vancomycin, and 750 mg PO levofloxacin for failed outpatient therapy of periorbital cellulitis. Orbital cellulitis was ruled out with a CT Orbit with IV contrast. In the inpatient setting 100 mg PO doxycycline was added to her antibiotic regimen. Her oedema and erythema improved with treatment, and she was discharged home with close follow-up and instructed to complete a total of 7 days of levofloxacin and doxycycline. |
|  | <u>Country</u><br>USA<br><u>Quality rating</u><br>Scored 6 out of 8 on the JBI critical appraisal checklist for case reports | <u>Eyebrows</u><br><u>Time frame</u><br>Same day |  |
| Soltany 2023<br>A rare case of microblading complicated by orbital exenteration and frontal defect reconstructed using crane principle<br>https://doi.org/10.1097/GOX.0000000000004857 | <u>Study design</u><br>Case report<br><u>Outcome/s of interest</u><br>Cellulitis<br>Necrotising fasciitis<br><u>Comparator intervention or control</u><br>NA<br><u>Data collection methods</u><br>NA<br><u>Country</u><br>Syria<br><u>Quality rating</u><br>Scored 8 out of 8 on the JBI critical appraisal checklist for case reports | <u>Sample size</u><br>1<br><u>Participants</u><br>43-year-old woman<br><u>Setting</u><br>Commercial (local beauty centre)<br><u>Practitioner</u><br>NR<br><u>Punctured site</u><br>Eyebrows<br><u>Time frame</u><br>4 days | A 43-year-old woman with diabetes had significant periocular facial eruption with edema four days after microblading for both eyebrows, and another few days of local signs of right periorbital and orbital cellulitis were noticed. Necrosis started at the 10th day. She developed right eye proptosis and painful eye movement, besides associated systemic manifestations of infection. Periorbital (preseptal) and orbital cellulitis were diagnosed. Despite treatment, the patient developed progressive blurry vision, winding up with vision loss with a vast periorbital necrosis representing a case of necrotising fasciitis. There was a 12 × 7 cm forehead defect with bony exposure measuring 7 × 4 cm, surrounded by granulation tissue. |
| Suleman 2023<br>Chronic granulomatous reaction to semi-permanent eyebrow tint<br>https://doi.org/10.7759/cureus.44070 | <u>Study design</u><br>Case report<br><u>Outcome/s of interest</u><br>Chronic granulomatous dermatitis<br><u>Comparator intervention or control</u><br>NA<br><u>Data collection methods</u> | <u>Sample size</u><br>1<br><u>Participants</u><br>49-year-old Hispanic woman<br><u>Setting</u><br>Unclear<br><u>Practitioner</u> | A 49-year-old Hispanic presented with a past history of type II diabetes mellitus and hypertension presented with a pruritic rash over her eyebrows, erythematous and indurated plaques over the entire left eyebrow and a smaller plaque with crusting over the right lateral eyebrow a month after microblading. Results of the biopsy showed discrete granulomas in the lower portions of the dermis, most of which were naked with no surrounding cuff of mononuclear cells. There was fine-black intracellular particulate material in the cytoplasm of histiocytes in the papillary dermis, but no polarizable material was evident. Given the biopsy results, negative chest X-ray findings, and normal ACE levels, the patient was diagnosed with a chronic granulomatous dermatitis secondary to microblading |
|  | NA<br><u>Country</u><br>Mexico<br><u>Quality rating</u><br>Scored 7 out of 8 on the JBI critical appraisal checklist for case reports | Unclear. Referred to as a doctor but later discussion suggests otherwise<br><u>Puncture site</u><br>Eyebrows<br><u>Time frame</u><br>NR | tint. |
| Sun 2023<br>Granulomatous reaction to permanent eyebrow makeup successfully treated with oral tofacitinib<br>https://doi.org/10.1111/jocd.15679 | <u>Study design</u><br>Case report<br><u>Outcome/s of interest</u><br>Granulomatous reaction<br><u>Comparator intervention or control</u><br>NA<br><u>Data collection methods</u><br>NA<br><u>Country</u><br>China<br><u>Quality rating</u><br>Scored 8 out of 8 on the JBI critical appraisal checklist for case reports | <u>Sample size</u><br>1<br><u>Participants</u><br>52-year-old woman<br><u>Setting</u><br>Commercial<br><u>Practitioner</u><br>NR<br><u>Puncture site</u><br>Eyebrows<br><u>Time frame</u><br>NR | A 52-year-old woman presented with multiple small subcutaneous papules along the eyebrows for half a year. Those papules developed with a delay of several weeks after permanent eyebrow makeup. The skin lesion was diagnosed as a granulomatous reaction to the colourant. |
| Tierney 2021<br>Koebnerization secondary to microblading<br>https://doi.org/10.1111/jocd.13695 | <u>Study design</u><br>Case report<br><u>Outcome/s of interest</u><br>Vitiligo, arising from the koebnerisation<br><u>Comparator intervention or control</u><br>NA | <u>Sample size</u><br>1<br><u>Participants</u><br>28-year-old woman<br><u>Setting</u><br>Commercial<br><u>Practitioner</u><br>NR | A 28-year-old Lithuanian woman presented with bilateral, symmetrical periocular depigmentation after 8 years of repeated eyebrow microblading. Examination under Woods light showed symmetrical, sharply demarcated, periocular depigmentation extending beyond both eyebrows and toward the medial canthi bilaterally. She was also noted to have a patch of depigmentation on her right forearm measuring 4 × 6 cm which had been stable for a number of years. She was diagnosed with vitiligo, arising from the koebnerisation effect of microblading. Tacrolimus 0.1% ointment was prescribed with advice on sun protection and avoidance for the depigmented areas. |
|  | <u>Data collection methods</u><br>NA<br><u>Country</u><br>UK<br><u>Quality rating</u><br>Scored 6 out of 8 on the JBI critical appraisal checklist for case reports | <u>Puncture site</u><br><u>Eyebrows</u><br><u>Time frame</u><br>8 years after repeated microblading |  |
| Tittelbach 2018<br>Sarcoidal foreign body reaction as a severe side-effect to permanent makeup successful treatment with intralesional triamcinolone<br>https://doi.org/10.2340/00015555-2876 | <u>Study design</u><br>Case report<br><u>Outcome/s of interest</u><br>Granulomatous foreign body reaction of sarcoidal type<br><u>Comparator intervention or control</u><br>NA<br><u>Data collection methods</u><br>NA<br><u>Country</u><br>Germany<br><u>Quality rating</u><br>Scored 7 out of 8 on the JBI critical appraisal checklist for case reports | <u>Sample size</u><br>1<br><u>Participants</u><br>41-year-old woman<br><u>Setting</u><br>Commercial<br><u>Practitioner</u><br>NR<br><u>Puncture site</u><br>Eyebrows<br><u>Time frame</u><br>18 months | A 41-year-old woman presented with itchy red papules developing in the tattooed area, starting on one side, 18 months previously. She had permanent makeup 10 years previously, which had been refreshed after 5 years. Biopsy revealed a "granulomatous foreign body reaction". The patient received laser treatment, which resulted in a temporary improvement, but was followed by new, now partially ulcerating, lesions. Dermatological examination revealed yellowish erythematous indurated papules with mild scaling accentuated on the medial eyebrows. Patch test showed positive test results at 72 h only for nickel (II) sulphate (+++) and methylisothiazolinone (++), which were not present in the tattoo material. A skin biopsy showed granulomatous accumulation of epithelioid cells in the whole dermis, surrounded by a sparse lymphocytic infiltrate and some multinucleated histiocytes on the dermatopathological investigation. A diagnosis of a granulomatous foreign body reaction of sarcoidal-type to tattoo-ink in the permanent makeup was made. |
| Tukenmez Demirci 2016<br>Is it a sarcoidal foreign-body granuloma or a cutaneous sarcoidosis on a permanent eyebrow make-up?<br>https://doi.org/10.3109/14764172.2015.1052516 | <u>Study design</u><br>Case report<br><u>Outcome/s of interest</u><br>Sarcoidal foreign-body reaction<br>Non-caseating granulomas<br><u>Comparator intervention or control</u> | <u>Sample size</u><br>1<br><u>Participants</u><br>34-year-old woman<br><u>Setting</u><br>Commercial<br><u>Practitioner</u> | A 34-year-old woman presented with multiple hard, shiny yellowish, 0.2–0.5-cm-sized papules on her eyebrows for a month. She has had cosmetic permanent make-up of these areas for several times during the last 10 years, with the last procedure 2 years prior. Histopathology of a papule disclosed several naked, non-caseating granulomas, which were composed of histiocytes, a few giant cells of foreign-body type, and sparse lymphocytes. All the laboratory investigations were within normal limits except for a mild elevation in angiotensin-converting enzyme or ACE level. A diagnosis of sarcoidal-type foreign-body reaction to tattoo pigments was made. |
|  | NA<br><u>Data collection methods</u><br>NA<br><u>Country</u><br>Turkey<br><u>Quality rating</u><br>Scored 7 out of 8 on the JBI critical appraisal checklist for case reports | NR<br><u>Puncture site</u><br>Eyebrows<br><u>Time frame</u><br>Procedures over the past 10 years with last one 2 years ago | The lesions mostly improved after topical corticosteroid treatment. |
| Valbuena 2017<br>Sarcoidal granulomatous reaction due to tattoos report of two cases<br>http://dx.doi.org/10.1590/abd1806-4841.20175860 | <u>Study design</u><br>Case report<br><u>Outcome/s of interest</u><br>Sarcoidal granulomatous<br><u>Comparator intervention or control</u><br>NA<br><u>Data collection methods</u><br>NA<br><u>Country</u><br>Colombia<br><u>Quality rating</u><br>Scored 6 out of 8 on the JBI critical appraisal checklist for case reports | <u>Sample size</u><br>1<br><u>Participants</u><br>55-year-old woman<br><u>Setting</u><br>Commercial<br><u>Practitioner</u><br>NR<br><u>Puncture site</u><br>Eyebrows<br><u>Time frame</u><br>16 years | A 55-year-old woman with a 2-year history of itchy persistent lesions on the eyebrows. She had a permanent makeup cosmetic tattoo 16 years before the onset of lesions. Histopathological examination of one of the lesions showed thick dermis with lymphocytic infiltrates around epithelioid granulomas involving the entire dermis, with presence of black pigment in the central portion. She was diagnosed with sarcoidal granulomatous reaction secondary to tattoo. |
| Vera 2018<br>Red and itchy bilateral supraorbital swellings<br>https://doi.org/10.1111/ddg.13687 | <u>Study design</u><br>Case report<br><u>Outcome/s of interest</u><br>Systemic sarcoidosis<br><u>Comparator intervention or control</u><br>NA | <u>Sample size</u><br>1<br><u>Participants</u><br>A 50-year-old woman<br><u>Setting</u><br>Commercial | A 50-year-old woman with Fitzpatrick skin type IV presented with bilateral, symmetrical, livid red, and itchy supraorbital skin swellings and loss of both eyebrows. The lesions had grown progressively in size over the previous four months at the sites where she had cosmetic tattooing of the eyebrows 7 years prior. A punch biopsy showed orthokeratosis and collagen bundles in the papillary dermis, with black pigment between the bundles. The dermis showed a dense nodular infiltrate with epithelioid nodules, histiocytes, many giant multinucleated Langerhans cells as well as |
|  | <u>Data collection methods</u><br>NA<br><u>Country</u><br>Unclear, probably<br>Venezuela<br><u>Quality rating</u><br>Scored 5 out of 8 on the JBI<br>critical appraisal checklist for<br>case reports | <u>Practitioner</u><br>NR<br><u>Puncture site</u><br>Eyebrows<br><u>Time frame</u><br>NA | intra- and extracellular foreign body-like black granulomas. A CT scan showed bilateral and symmetrical hilar lymphadenopathy, with lymph nodes up to 1.5 cm in diameter. A diagnosis of systemic sarcoidosis with tattoo-induced skin lesions was made. |

Key ACE: Angiotensin-converting enzyme; CT: Computerised tomography; NA: not applicable; NR: not reported; PO: Per os (by mouth); PPD: Purified-protein derivative

**Table 13.**
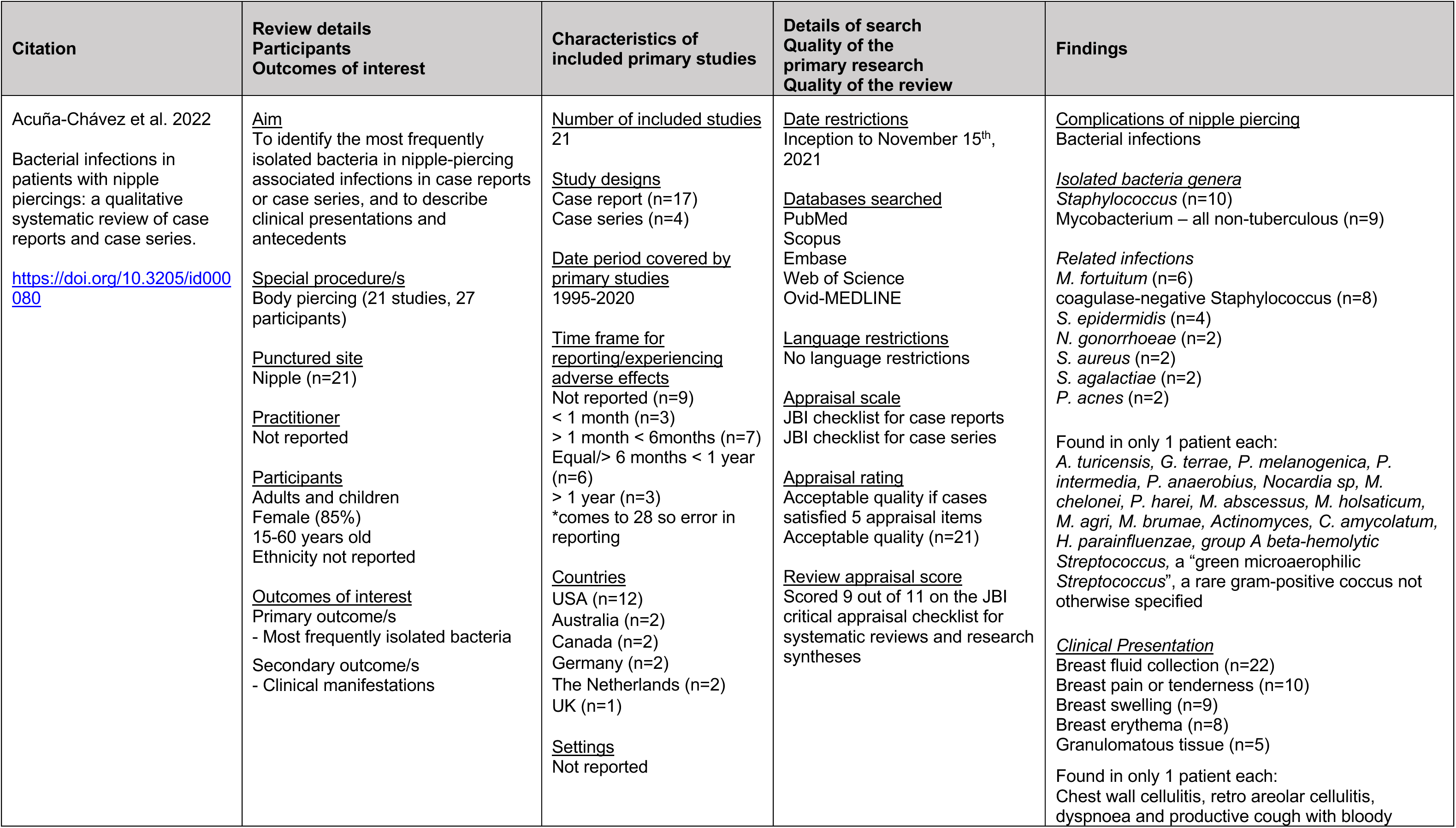
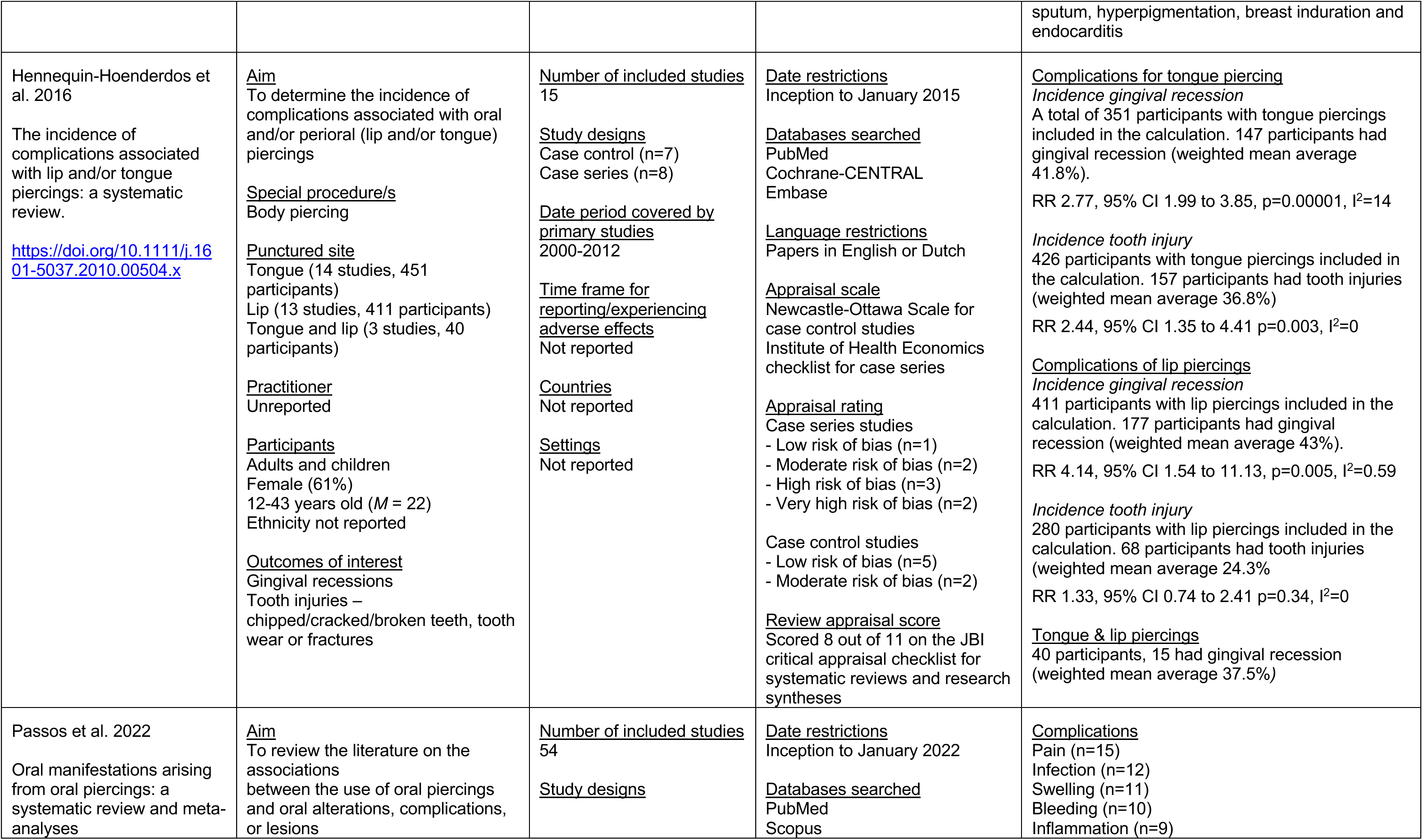
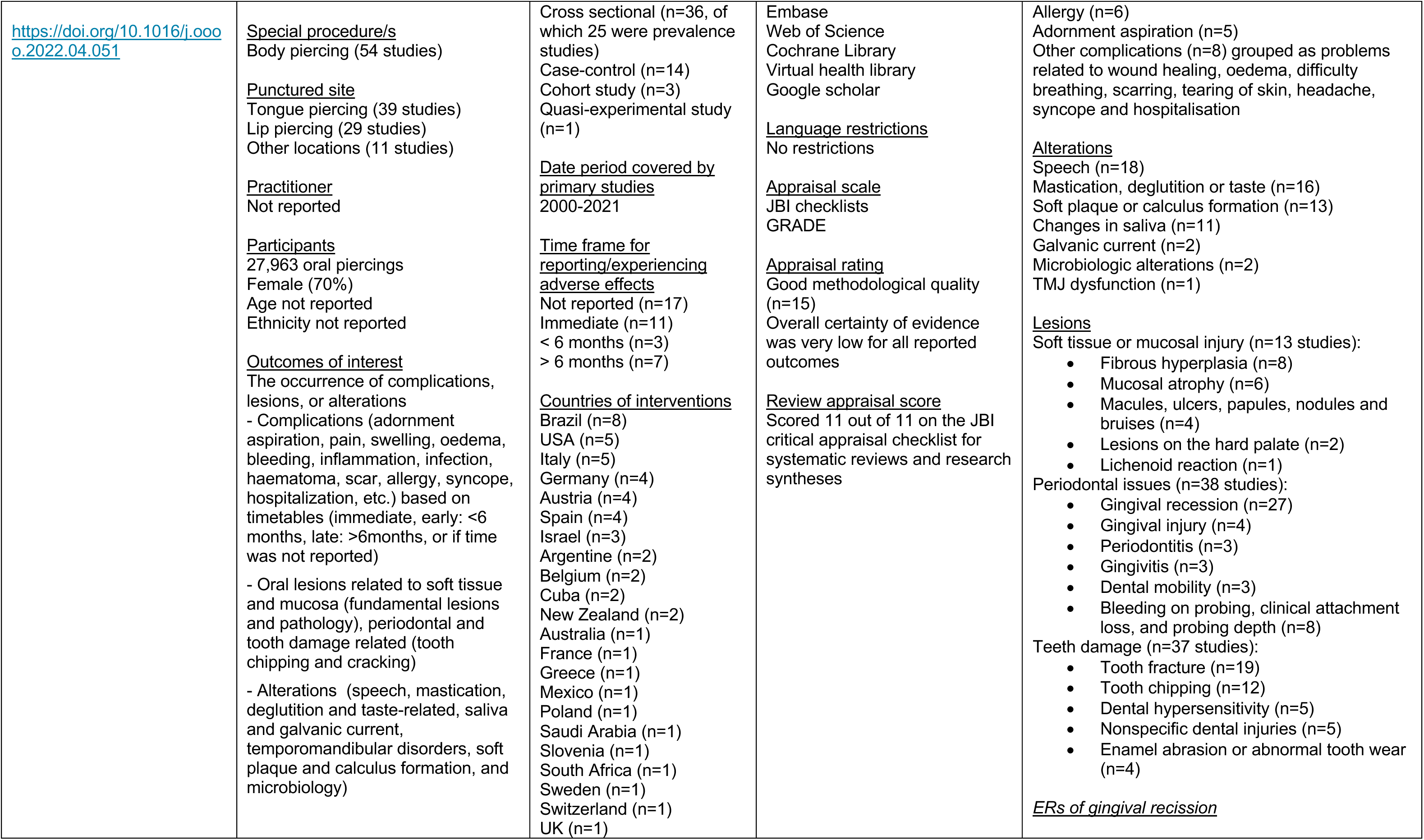
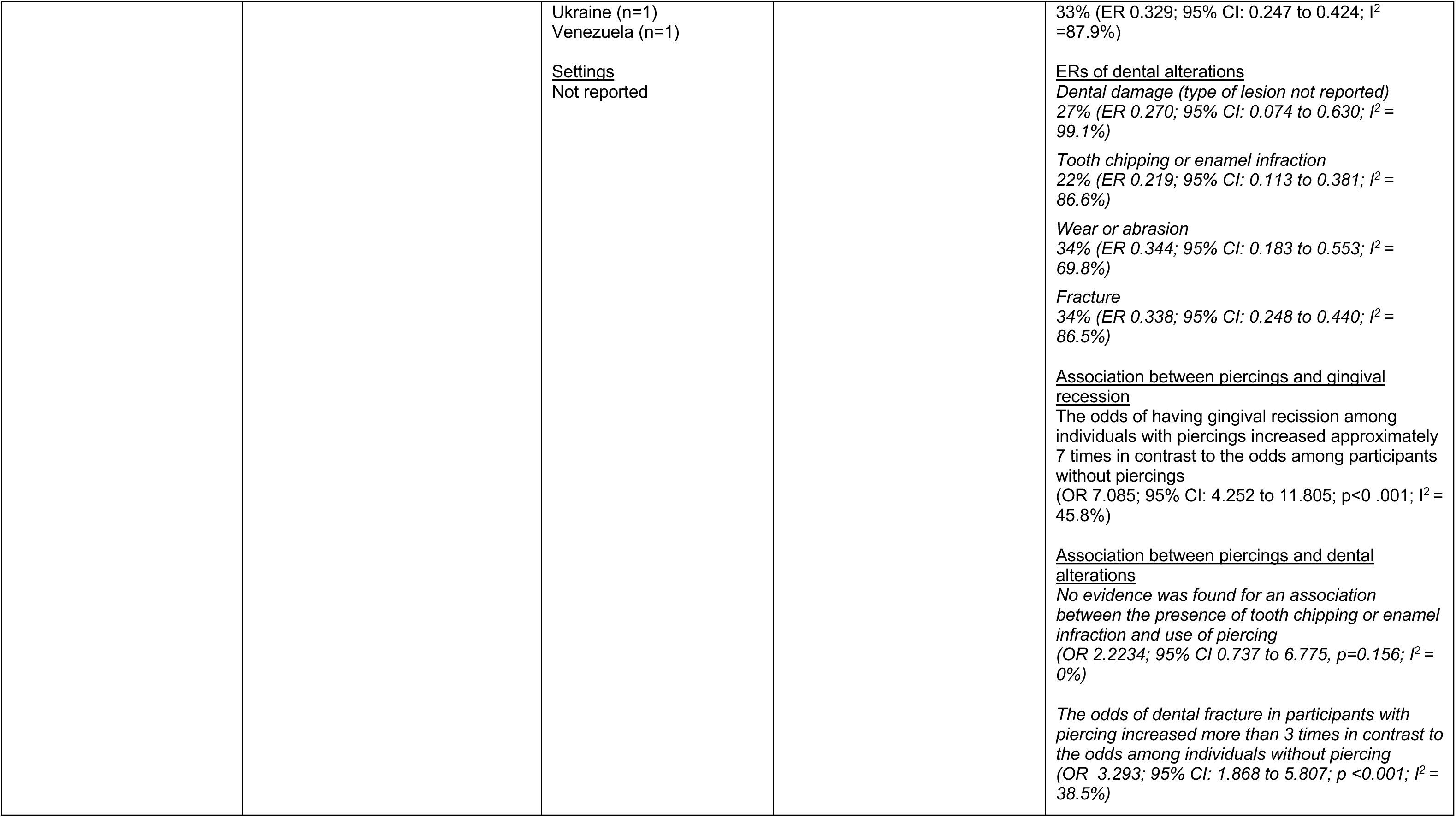
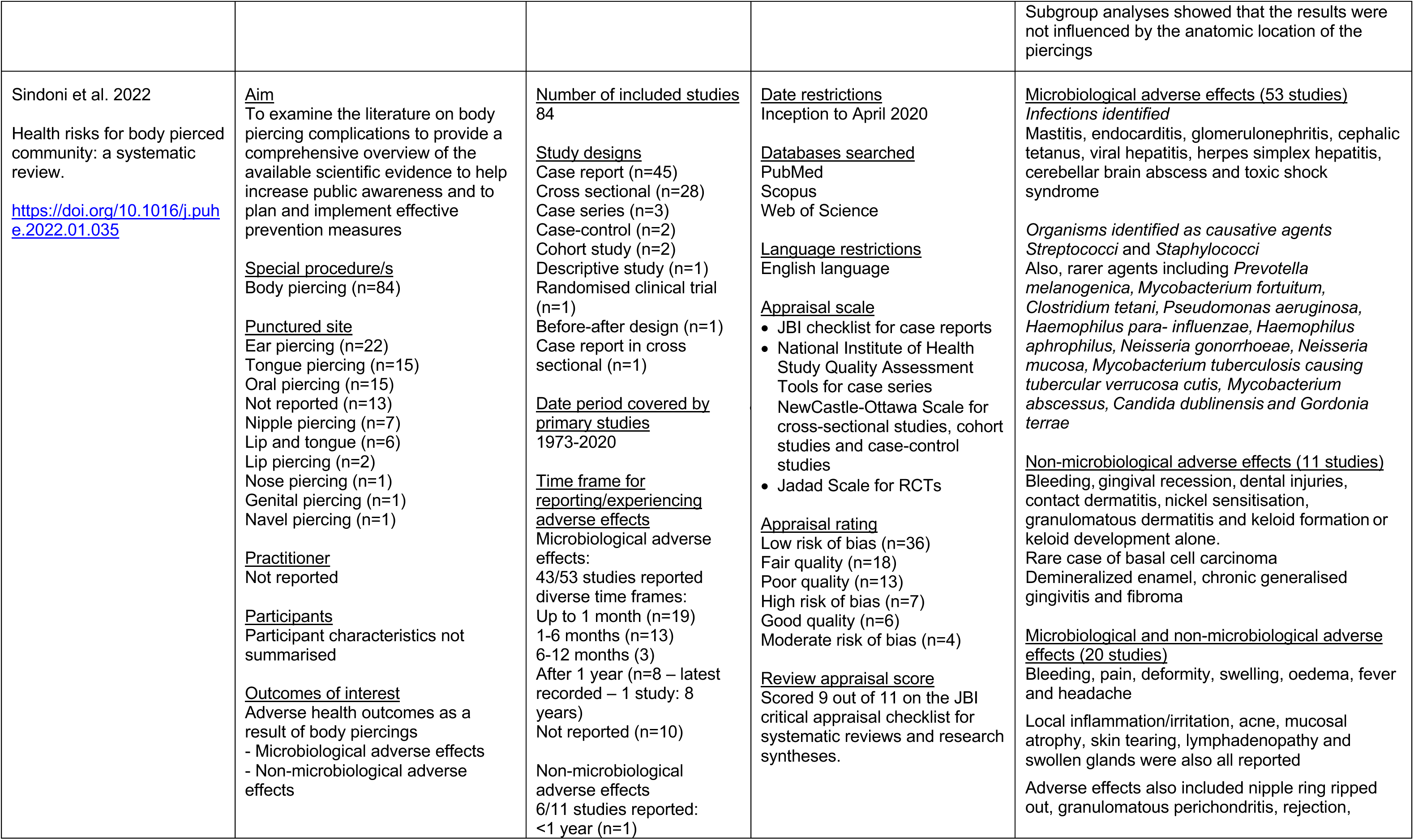
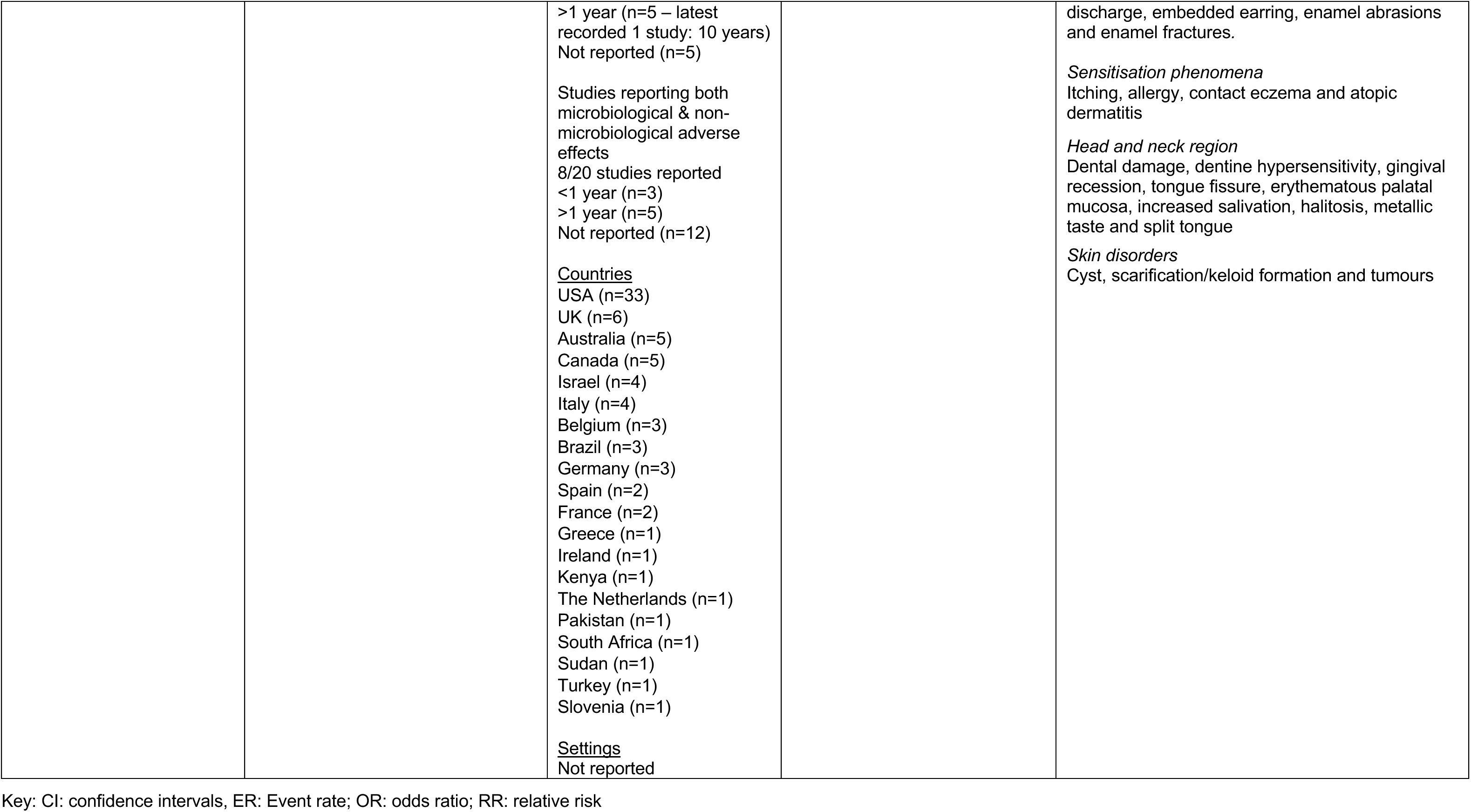
Summary of included systematic reviews – Body piercing.

**Table 14.**
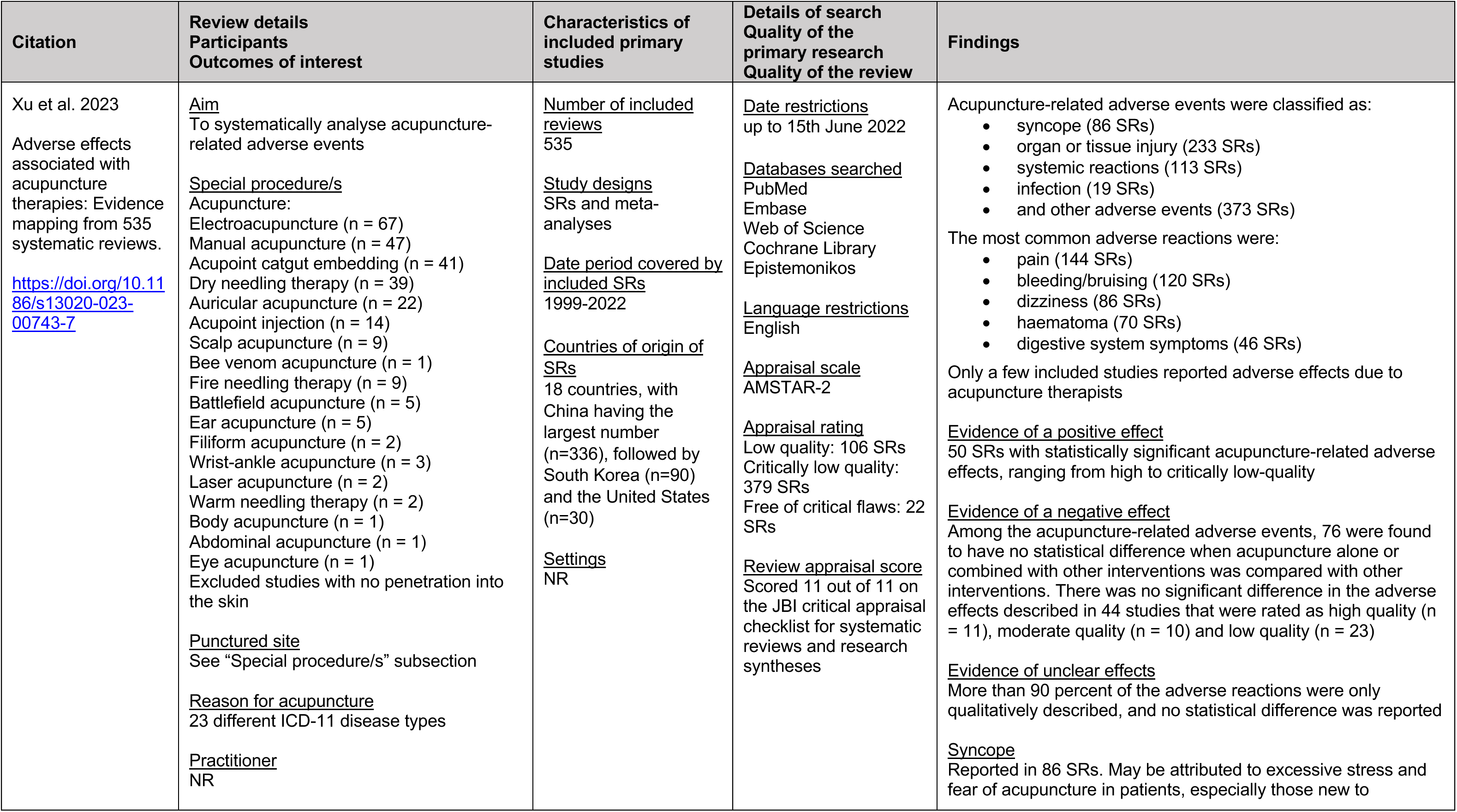
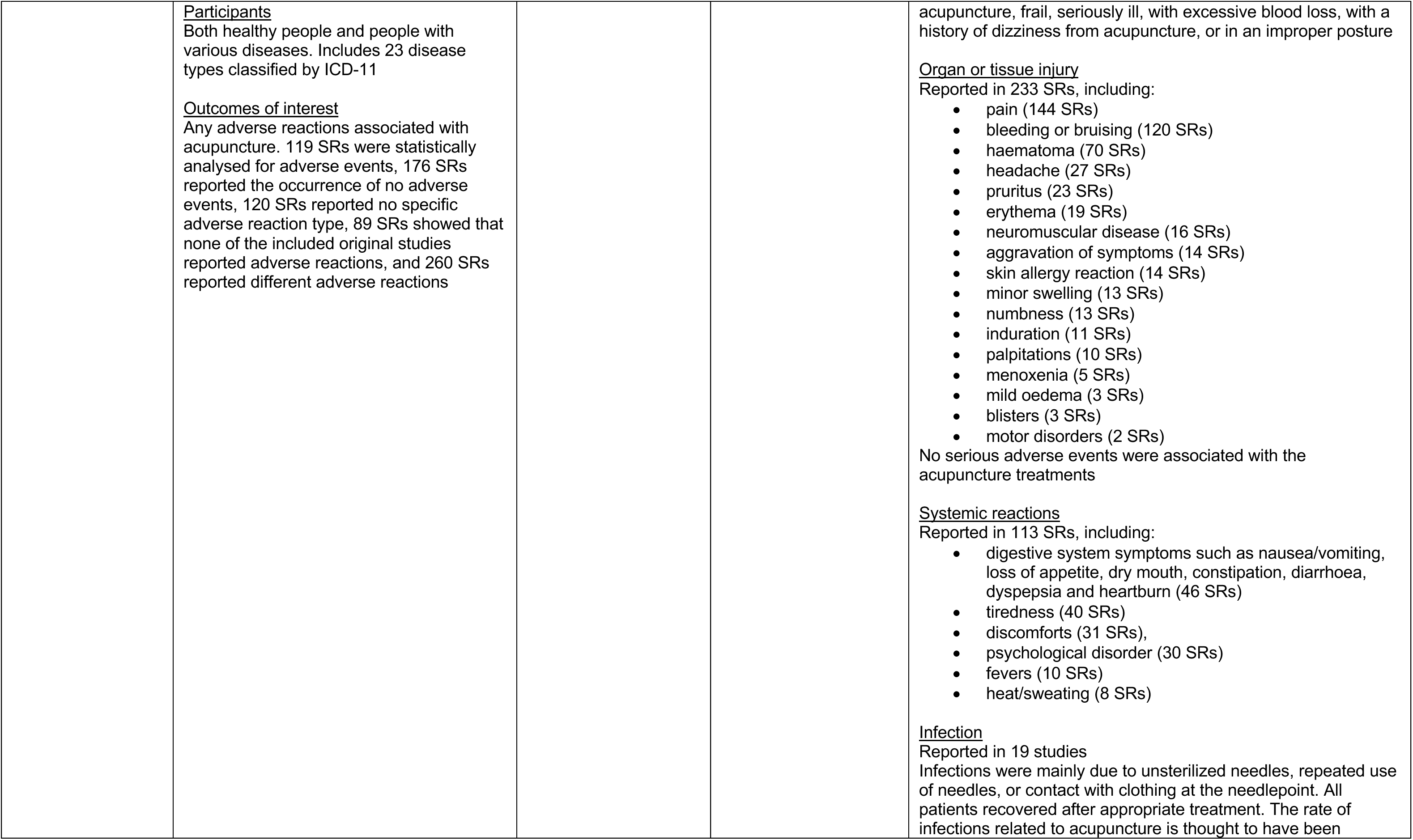
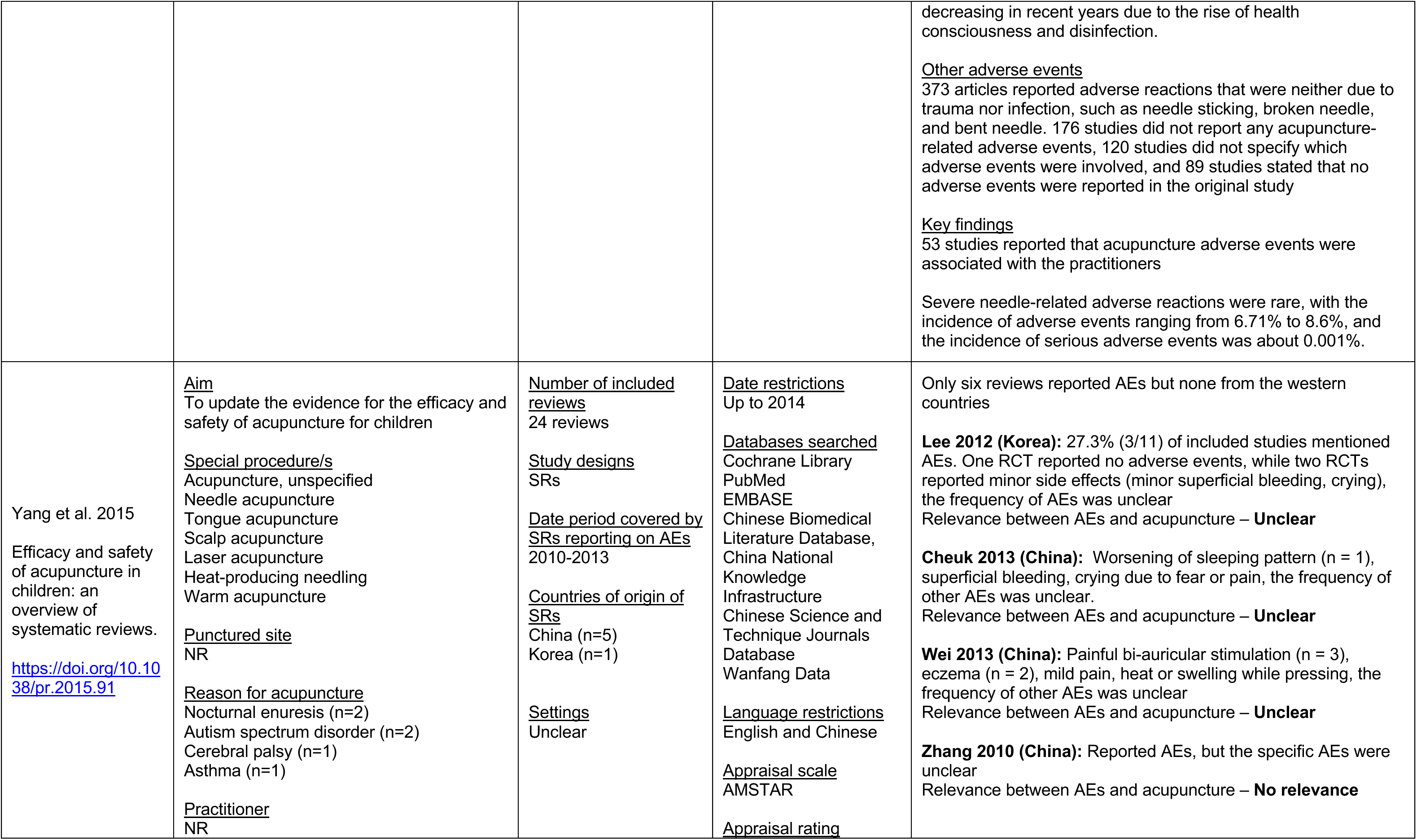
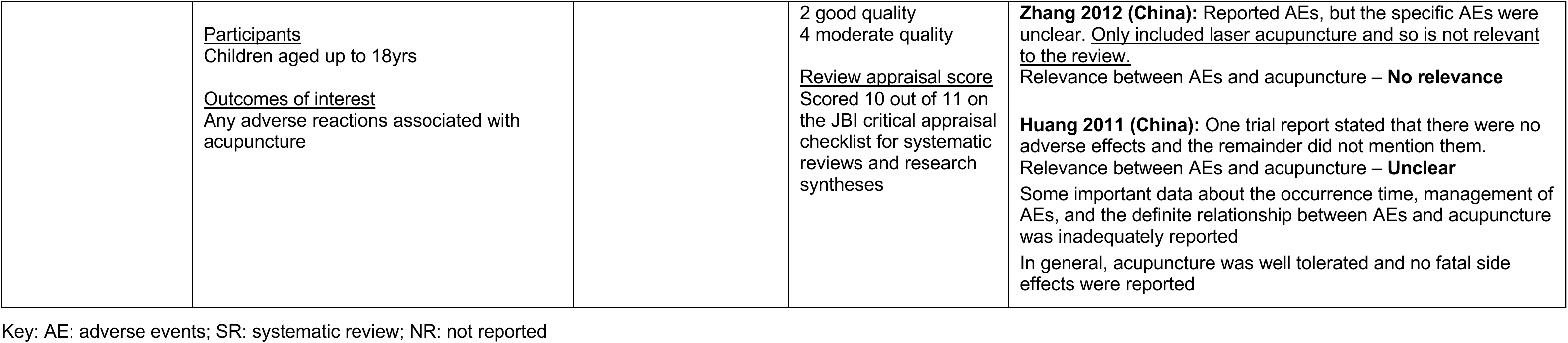
Summary of included umbrella reviews – Acupuncture.

| Citation | Review details<br>Participants<br>Outcomes of interest | Characteristics of<br>included primary<br>studies | Details of search<br>Quality of the<br>primary research<br>Quality of the review | Findings |
| --- | --- | --- | --- | --- |
| Xu et al. 2023<br>Adverse effects associated with acupuncture therapies: Evidence mapping from 535 systematic reviews.<br>https://doi.org/10.1186/s13020-023-00743-7 | <u>Aim</u><br>To systematically analyse acupuncture-related adverse events<br><u>Special procedure/s</u><br>Acupuncture:<br>Electroacupuncture (n = 67)<br>Manual acupuncture (n = 47)<br>Acupoint catgut embedding (n = 41)<br>Dry needling therapy (n = 39)<br>Auricular acupuncture (n = 22)<br>Acupoint injection (n = 14)<br>Scalp acupuncture (n = 9)<br>Bee venom acupuncture (n = 1)<br>Fire needling therapy (n = 9)<br>Battlefield acupuncture (n = 5)<br>Ear acupuncture (n = 5)<br>Filiform acupuncture (n = 2)<br>Wrist-ankle acupuncture (n = 3)<br>Laser acupuncture (n = 2)<br>Warm needling therapy (n = 2)<br>Body acupuncture (n = 1)<br>Abdominal acupuncture (n = 1)<br>Eye acupuncture (n = 1)<br>Excluded studies with no penetration into the skin<br><u>Punctured site</u><br>See “Special procedure/s” subsection<br><u>Reason for acupuncture</u><br>23 different ICD-11 disease types<br><u>Practitioner</u><br>NR | <u>Number of included reviews</u><br>535<br><u>Study designs</u><br>SRs and meta-analyses<br><u>Date period covered by included SRs</u><br>1999-2022<br><u>Countries of origin of SRs</u><br>18 countries, with China having the largest number (n=336), followed by South Korea (n=90) and the United States (n=30)<br><u>Settings</u><br>NR | <u>Date restrictions</u><br>up to 15th June 2022<br><u>Databases searched</u><br>PubMed<br>Embase<br>Web of Science<br>Cochrane Library<br>Epistemonikos<br><u>Language restrictions</u><br>English<br><u>Appraisal scale</u><br>AMSTAR-2<br><u>Appraisal rating</u><br>Low quality: 106 SRs<br>Critically low quality: 379 SRs<br>Free of critical flaws: 22 SRs<br><u>Review appraisal score</u><br>Scored 11 out of 11 on the JBI critical appraisal checklist for systematic reviews and research syntheses | Acupuncture-related adverse events were classified as:<br>• syncope (86 SRs)<br>• organ or tissue injury (233 SRs)<br>• systemic reactions (113 SRs)<br>• infection (19 SRs)<br>• and other adverse events (373 SRs)<br>The most common adverse reactions were:<br>• pain (144 SRs)<br>• bleeding/bruising (120 SRs)<br>• dizziness (86 SRs)<br>• haematoma (70 SRs)<br>• digestive system symptoms (46 SRs)<br>Only a few included studies reported adverse effects due to acupuncture therapists<br><u>Evidence of a positive effect</u><br>50 SRs with statistically significant acupuncture-related adverse effects, ranging from high to critically low-quality<br><u>Evidence of a negative effect</u><br>Among the acupuncture-related adverse events, 76 were found to have no statistical difference when acupuncture alone or combined with other interventions was compared with other interventions. There was no significant difference in the adverse effects described in 44 studies that were rated as high quality (n = 11), moderate quality (n = 10) and low quality (n = 23)<br><u>Evidence of unclear effects</u><br>More than 90 percent of the adverse reactions were only qualitatively described, and no statistical difference was reported<br><u>Syncope</u><br>Reported in 86 SRs. May be attributed to excessive stress and fear of acupuncture in patients, especially those new to |

|  |  |  |  |
| --- | --- | --- | --- |
|  | <u>Participants</u><br>Both healthy people and people with various diseases. Includes 23 disease types classified by ICD-11<br><u>Outcomes of interest</u><br>Any adverse reactions associated with acupuncture. 119 SRs were statistically analysed for adverse events, 176 SRs reported the occurrence of no adverse events, 120 SRs reported no specific adverse reaction type, 89 SRs showed that none of the included original studies reported adverse reactions, and 260 SRs reported different adverse reactions |  | acupuncture, frail, seriously ill, with excessive blood loss, with a history of dizziness from acupuncture, or in an improper posture<br><u>Organ or tissue injury</u><br>Reported in 233 SRs, including:<br>• pain (144 SRs)<br>• bleeding or bruising (120 SRs)<br>• haematoma (70 SRs)<br>• headache (27 SRs)<br>• pruritus (23 SRs)<br>• erythema (19 SRs)<br>• neuromuscular disease (16 SRs)<br>• aggravation of symptoms (14 SRs)<br>• skin allergy reaction (14 SRs)<br>• minor swelling (13 SRs)<br>• numbness (13 SRs)<br>• induration (11 SRs)<br>• palpitations (10 SRs)<br>• menoxenia (5 SRs)<br>• mild oedema (3 SRs)<br>• blisters (3 SRs)<br>• motor disorders (2 SRs)<br>No serious adverse events were associated with the acupuncture treatments<br><u>Systemic reactions</u><br>Reported in 113 SRs, including:<br>• digestive system symptoms such as nausea/vomiting, loss of appetite, dry mouth, constipation, diarrhoea, dyspepsia and heartburn (46 SRs)<br>• tiredness (40 SRs)<br>• discomforts (31 SRs),<br>• psychological disorder (30 SRs)<br>• fevers (10 SRs)<br>• heat/sweating (8 SRs)<br><u>Infection</u><br>Reported in 19 studies<br>Infections were mainly due to unsterilized needles, repeated use of needles, or contact with clothing at the needlepoint. All patients recovered after appropriate treatment. The rate of infections related to acupuncture is thought to have been |
|  |  |  |  | decreasing in recent years due to the rise of health consciousness and disinfection.<br><u>Other adverse events</u><br>373 articles reported adverse reactions that were neither due to trauma nor infection, such as needle sticking, broken needle, and bent needle. 176 studies did not report any acupuncture-related adverse events, 120 studies did not specify which adverse events were involved, and 89 studies stated that no adverse events were reported in the original study<br><u>Key findings</u><br>53 studies reported that acupuncture adverse events were associated with the practitioners<br>Severe needle-related adverse reactions were rare, with the incidence of adverse events ranging from 6.71% to 8.6%, and the incidence of serious adverse events was about 0.001%. |
| Yang et al. 2015<br>Efficacy and safety of acupuncture in children: an overview of systematic reviews.<br>https://doi.org/10.1038/pr.2015.91 | <u>Aim</u><br>To update the evidence for the efficacy and safety of acupuncture for children<br><u>Special procedure/s</u><br>Acupuncture, unspecified<br>Needle acupuncture<br>Tongue acupuncture<br>Scalp acupuncture<br>Laser acupuncture<br>Heat-producing needling<br>Warm acupuncture<br><u>Punctured site</u><br>NR<br><u>Reason for acupuncture</u><br>Nocturnal enuresis (n=2)<br>Autism spectrum disorder (n=2)<br>Cerebral palsy (n=1)<br>Asthma (n=1)<br><u>Practitioner</u><br>NR | <u>Number of included reviews</u><br>24 reviews<br><u>Study designs</u><br>SRs<br><u>Date period covered by SRs reporting on AEs</u><br>2010-2013<br><u>Countries of origin of SRs</u><br>China (n=5)<br>Korea (n=1)<br><u>Settings</u><br>Unclear | <u>Date restrictions</u><br>Up to 2014<br><u>Databases searched</u><br>Cochrane Library<br>PubMed<br>EMBASE<br>Chinese Biomedical Literature Database,<br>China National Knowledge Infrastructure<br>Chinese Science and Technique Journals Database<br>Wanfang Data<br><u>Language restrictions</u><br>English and Chinese<br><u>Appraisal scale</u><br>AMSTAR<br><u>Appraisal rating</u> | Only six reviews reported AEs but none from the western countries<br>**Lee 2012 (Korea):** 27.3% (3/11) of included studies mentioned AEs. One RCT reported no adverse events, while two RCTs reported minor side effects (minor superficial bleeding, crying), the frequency of AEs was unclear<br>Relevance between AEs and acupuncture – **Unclear**<br>**Cheuk 2013 (China):** Worsening of sleeping pattern (n = 1), superficial bleeding, crying due to fear or pain, the frequency of other AEs was unclear.<br>Relevance between AEs and acupuncture – **Unclear**<br>**Wei 2013 (China):** Painful bi-auricular stimulation (n = 3), eczema (n = 2), mild pain, heat or swelling while pressing, the frequency of other AEs was unclear<br>Relevance between AEs and acupuncture – **Unclear**<br>**Zhang 2010 (China):** Reported AEs, but the specific AEs were unclear<br>Relevance between AEs and acupuncture – **No relevance** |
|  | <u>Participants</u><br>Children aged up to 18yrs<br><u>Outcomes of interest</u><br>Any adverse reactions associated with acupuncture |  | 2 good quality<br>4 moderate quality<br><u>Review appraisal score</u><br>Scored 10 out of 11 on the JBI critical appraisal checklist for systematic reviews and research syntheses | **Zhang 2012 (China):** Reported AEs, but the specific AEs were unclear. <u>Only included laser acupuncture and so is not relevant to the review.</u><br>Relevance between AEs and acupuncture – **No relevance**<br>**Huang 2011 (China):** One trial report stated that there were no adverse effects and the remainder did not mention them.<br>Relevance between AEs and acupuncture – **Unclear**<br>Some important data about the occurrence time, management of AEs, and the definite relationship between AEs and acupuncture was inadequately reported<br>In general, acupuncture was well tolerated and no fatal side effects were reported |

Key: AE: adverse events; SR: systematic review; NR: not reported

**Table 15.** Summary of included primary research – Electrolysis.

| Citation | Study details<br>Quality ratings | Participants<br>Setting<br>Time frame of reporting or experiencing<br>adverse effects | Findings |
| --- | --- | --- | --- |
| Morand et al.<br>2015<br>Systemic sarcoidosis revealed by axillary electrolysis<br>https://doi.org/10.1177/1203475415575016 | <u>Study design</u><br>Case report<br><u>Outcome/s of interest</u><br>Cutaneous sarcoidosis<br><u>Comparator intervention or control</u><br>NA<br><u>Data collection methods</u><br>Clinical examination<br><u>Country</u><br>Canada<br><u>Quality rating</u><br>Scored 6 out of 8 on the JBI critical appraisal checklist for case reports | <u>Sample size</u><br>1<br><u>Participants</u><br>35-year-old woman<br><u>Setting</u><br>NR<br><u>Practitioner</u><br>NR<br><u>Time frame</u><br>NA | The patient received electrolysis for many years and developed lesions distributed symmetrically bilaterally after her last treatment. On physical examination, follicular brownish papules were noted. The diagnosis of cutaneous sarcoidosis was suggested by the clinical manifestations and confirmed by histopathologic findings. This case report suggests that sarcoidosis can arise from skin that has received electrolysis hair removal treatments. |

Key: NA: not applicable; NR: not reported

### 6.3 Quality appraisal

**Table 16.** JBI critical appraisal scores of systematic reviews.

| Study | JBI Appraisal items |  |  |  |  |  |  |  |  |  |  | Score | Overall quality<sup>6</sup> |
| --- | --- | --- | --- | --- | --- | --- | --- | --- | --- | --- | --- | --- | --- |
|  | 1 | 2 | 3 | 4 | 5 | 6 | 7 | 8 | 9 | 10 | 11 |  |  |
| Tattooing |  |  |  |  |  |  |  |  |  |  |  |  |  |
| Sindoni et al. 2021 | Y | Y | Y | Y | Y | Y | U | Y | N | Y | n/a | 8 | + Moderate |
| Body piercing |  |  |  |  |  |  |  |  |  |  |  |  |  |
| Acuña-Chavez et al. 2022 | Y | Y | Y | Y | Y | U | Y | Y | N | Y | Y | 9 | + Moderate |
| Hennequin-Hoenderdos et al. 2016 | Y | Y | Y | Y | Y | Y | Y | N | N | Y | N | 8 | - Low |
| Passos et al. 2022 | Y | Y | Y | Y | Y | Y | Y | Y | Y | Y | Y | 11 | ++ High |
| Sindoni et al. 2022 | Y | Y | Y | Y | Y | Y | U | Y | N | Y | Y | 9 | + Moderate |
| Acupuncture |  |  |  |  |  |  |  |  |  |  |  |  |  |
| Yang et al. 2015 | Y | Y | Y | Y | Y | Y | Y | Y | Y | N | Y | 10 | ++ High |
| Xu et al. 2023 | Y | Y | Y | Y | Y | Y | Y | Y | Y | Y | Y | 11 | ++ High |

Key: Y – Yes; N – No; U – Unclear; n/a – not applicable
1. Is the review question clearly and explicitly stated? 2. Were the inclusion criteria appropriate for the review question? 3. **Was the search strategy appropriate?** 4. **Were the sources and resources used to search for studies adequate?** 5. **Were the criteria for appraising studies appropriate?** 6. Was critical appraisal conducted by two or more reviewers independently? 7. Were there methods to minimize errors in data extraction? 8. **Were the methods used to combine studies appropriate?** 9. Was the likelihood of publication bias assessed? 10. Were recommendations for policy and/or practice supported by the reported data? 11. Were the specific directives for new research appropriate?

**Table 17.**
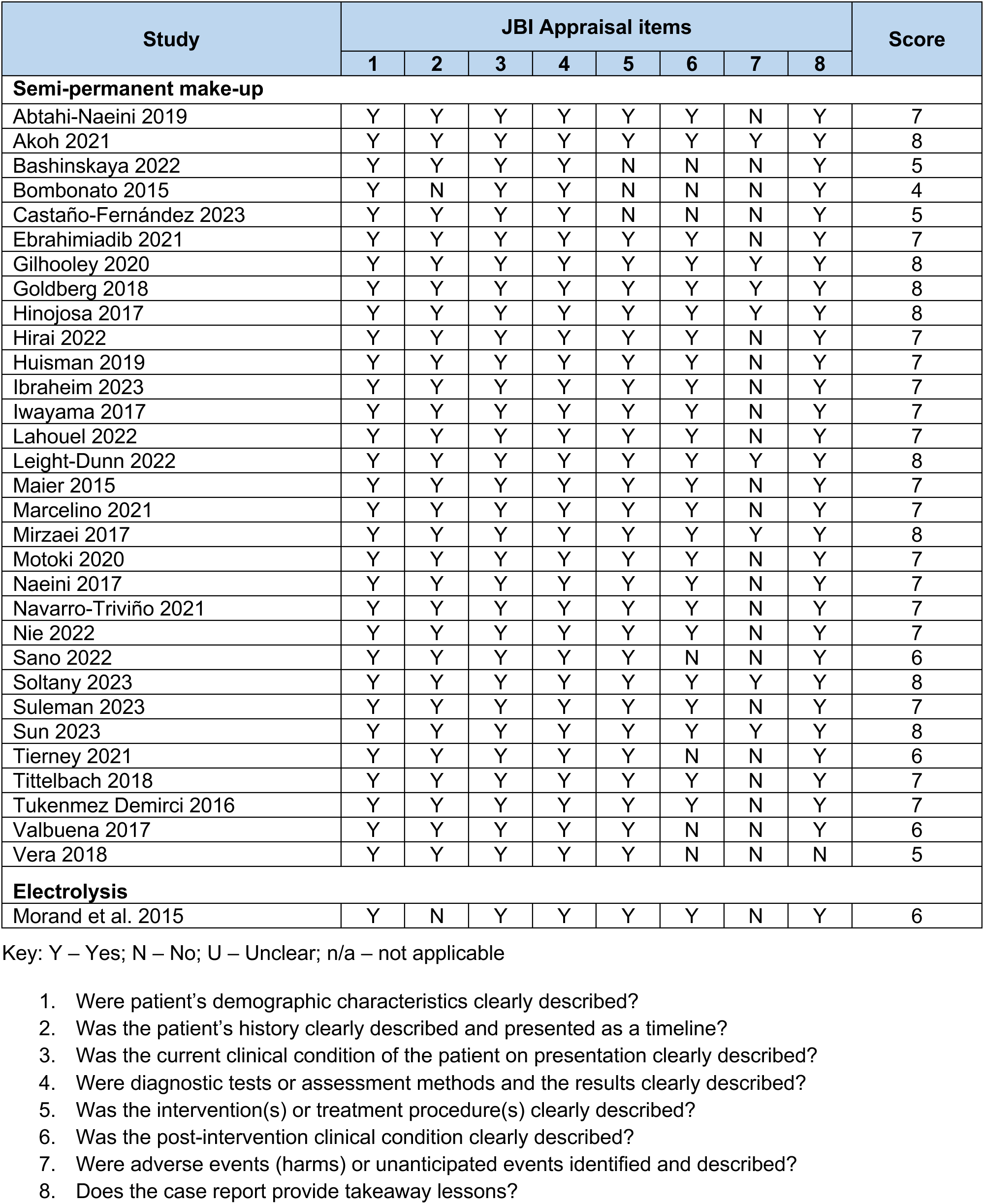
JBI critical appraisal scores for case reports.

### 6.4 Information available on request

The protocol is available on request.

## 7. ADDITIONAL INFORMATION

### 7.2 Acknowledgements

The authors would like to thank Sarah Jones, Sally Anstey and Nathan Davies for their contributions during stakeholder meetings in guiding the focus of the review. We would also like thank Nia Davies and Isobel Davies for their contributions to carrying out this review.

## 8. APPENDIX

## APPENDIX 1: Glossary

***Achromobacter turicensis***: A species of bacteria found in the environment that can occasionally cause infections in humans, particularly in individuals with compromised immune systems.

***Actinomyces***: A genus of bacteria commonly found in the oral, gastrointestinal, and genital tracts, typically harmless but can cause infections, particularly actinomycosis, when invading deeper tissues.

***Aggregatibacter aphrophilus*** (formerly ***Haemophilus aphrophilus*):** Bacteria found in the oral cavity, typically harmless but can cause infections like endocarditis and brain abscesses.

**Breast induration:** Firmness or hardness in the breast tissue.

***Candida dubliniensis***: A yeast species that can cause fungal infections, particularly in individuals with weakened immune systems, often affecting the oral cavity.

**Cellulitis:** A bacterial skin infection marked by redness, swelling, and tenderness, often occurring due to bacteria entering through skin break.

***Cephalic tetanus***: A rare form of tetanus, a potentially life-threatening bacterial infection caused by *Clostridium tetani*.

**Cerebellar brain abscess:** A rare medical condition characterised by the formation of a pus-filled cavity (abscess) within the cerebellum, a region located at the back of the brain responsible for coordinating movements and maintaining balance.

***Clostridium tetani:*** A bacterium responsible for causing tetanus, a serious bacterial infection affecting the nervous system.

**Conjunctival injection:** Redness or dilation of the blood vessels in the conjunctiva, which is the thin, transparent layer of tissue that covers the white part of the eye (sclera) and lines the inside of the eyelid.

***Corynebacterium amycolatum*:** A bacterium typically residing harmlessly on human skin and mucous membranes but which can cause infections in individuals with weakened immune systems.

**Cutaneous diphtheria:** A skin infection caused by the bacterium Corynebacterium diphtheriae.

**Cutaneous lymphoid hyperplasia:** A benign skin condition marked by increased lymphocytes, macrophages, and dendritic cells in the skin, resulting in raised skin lesions.

**Dental mobility**: Dental mobility refers to the ability of a tooth to move within its socket in the jawbone.

**Dentine hypersensitivity**: Tooth sensitivity that is characterised by brief, sharp pain triggered by various stimuli due to exposed dentine.

**Dermatofibroma:** A benign skin nodule or bump, often found on the limbs, that typically appears as a small, firm, reddish-brown or tan growth in the skin.

**Dermatofibrosarcoma protuberans:** A rare, slow growing and locally aggressive skin tumour.

**Dyspepsia**: Upset stomach.

**Dyspnoea:** Difficulty breathing.

**Eczema**: A common skin condition characterized by redness, itching, and inflammation of the skin.

**Endocarditis:** Inflammation of the heart’s inner lining and valves.

**Epithelioid osteoblastoma:** A rare bone tumour.

**Erythema:** Redness of the skin, often caused by increased blood flow to the skin’s blood vessels.

**Erythematous palatal mucosa**: Red or reddened appearance of the mucous membrane (soft tissue) on the palate, which is the roof of the mouth.

**Fibroma:** A benign (non-cancerous) tumour that originates from fibrous or connective tissue.

**Fibrous hyperplasia:** A benign overgrowth of fibrous tissue in the oral cavity.

**Gingival recession**: The condition in which the gum tissue around teeth recedes or pulls back, exposing more of the tooth’s root,

**Gingivitis:** Gingivitis is the mildest and earliest form of gum disease (periodontal disease), characterised by inflammation of the gums (gingiva).

**Glomerulonephritis**: A group of kidney diseases characterised by inflammation and damage to the glomeruli, the tiny filtering units in the kidneys.

***Gordonia terrae*:** A bacterium commonly found in soil, occasionally causing infections in individuals with a compromised immune system.

**Granulomatous dermatitis:** Inflammation of the skin characterized by the formation of granulomas.

**Granulomatous perichondritis:** Inflammation and granuloma formation in the tissue surrounding cartilage, often affecting the ears or nose, leading to symptoms like pain, swelling, and deformity.

**Granulomatous tissue:** Tissue reaction involving the formation of organised clusters of immune cells, called granulomas, in response to infections, foreign substances, or inflammatory conditions.

**Haemangioma**: A noncancerous tumour of blood vessels.

**Haematoma**: A localized collection of blood outside blood vessels, typically caused by injury or bleeding,

***Haemophilus parainfluenzae***: A bacterium found in human respiratory and oral cavities, usually harmless but can cause infections, especially in immunocompromised individuals.

**Halitosis**: Persistent and unpleasant bad breath

**Hepatitis:** Inflammation of the liver, which can be caused by infections, toxins, or autoimmune reactions.

**Herpes simplex hepatitis:** A rare and serious condition in which the herpes simplex virus infects and causes inflammation in the liver

**Hyperpigmentation**: Darkening of the skin due to excess melanin.

**Induration:** Refers to the process of hardening or thickening of a tissue or organ.

**Keloid formation**: The creation of raised, enlarged scars called keloids, often resulting from excessive collagen production during the healing process.

**Keratoacanthoma:** A benign, rapidly growing skin tumour that typically appears as a dome-shaped nodule with a central crater or horn in the middle.

**Koebnerization (Koebner Phenomenon):** Skin conditions developing or worsening in areas of skin subjected to injury or trauma.

**Leiomyosarcoma:** A tumour of smooth muscle cells.

**Lichenoid reaction**: A skin condition characterized by the appearance of lichen planus-like eruptions on the tattooed area.

**Löfgren’s syndrome:** A specific acute form of sarcoidosis characterized by symptoms such as fever, joint pain, erythema nodosum (painful skin nodules), and bilateral hilar lymphadenopathy (enlarged lymph nodes in the chest), typically with a good prognosis.

**Lymphadenopathy:** The enlargement or swelling of lymph nodes

**Mastitis:** Inflammation of the breast tissue, often caused by bacterial infection, which can result in pain, redness, swelling, and other symptoms

**Menoxenia**: An abnormal or irregular menstrual cycle.

***Molluscum contagiosum*:** Viral skin infection with small, flesh-coloured bumps.

**Monkeypox (Mpox) infection**: A rare viral disease similar to smallpox, transmitted from animals to humans, causing fever and skin rash.

**Mucosal atrophy:** Thinning, weakening, or shrinking of the mucous membrane lining in a body organ or cavity.

***Mycobacterium chelonae* Infection**: A specific type of bacterial infection caused by the bacterium *Mycobacterium chelonae*.

**Necrotising fasciitis**: A severe bacterial infection causing rapid tissue death.

***Neisseria gonorrhoeae:*** A bacterium responsible for the sexually transmitted infection (STI) known as gonorrhoea.

***Neisseria mucosa:*** A bacterium commonly found in the upper respiratory tract and mucous membranes of the human body.

***Nocardia* species:** A diverse group of bacteria belonging to the Nocardia genus found in the environment and capable of causing infections, particularly in those with weakened immune systems.

**Nodular dermal lymphohistiocytic infiltration**: Abnormal accumulation of immune cells in the dermal layer of the skin.

**Non-tuberculous *Mycobacterial* infections:** An infection caused by mycobacteria other than Mycobacterium tuberculosis, which can affect various body systems, including the lungs, skin, and soft tissues.

**Oedema**: Abnormal accumulation of excess fluid within the body’s tissue.

**Oral Galvanism:** The phenomenon in the mouth where dissimilar dental materials create an electrical circuit, causing symptoms like pain, metallic taste, sensitivity, and gum irritation.

**Orbital cellulitis:** A serious medical condition characterised by a bacterial infection and inflammation of the tissues within the eye socket (orbit).

**Papulo-nodular reactions**: Refer to skin changes characterised by the development of small papules and nodules on the skin’s surface.

**Periodontitis:** A severe form of gum disease (periodontal disease) characterised by inflammation and infection of the tissues supporting the teeth, including the gums, ligaments, and bone

**Periorbital cellulitis:** Infection and inflammation of the soft tissues in the eyelid and the skin around the eye.

***Prevotella***: A genus of anaerobic bacteria commonly found in the human body, including the oral, gastrointestinal, and genital tracts, with the potential for causing infections.

**Priapism:** Painful, prolonged erection unrelated to sexual arousal, requiring immediate medical attention.

***Propionibacterium:*** A genus of bacteria often involving skin and soft tissues, including conditions like acne and surgical site infections.

**Proptosis**: Abnormal protrusion or bulging of an eyeball from its normal position within the eye socket (orbit).

**Pruritus**: Itching.

**Pseudoepitheliomatous hyperplasia:** Unusual proliferation of skin cells that mimics the appearance of squamous cell carcinoma.

***Pseudomonas aeruginosa:*** A pathogenic bacterium known for causing various infections in the body.

**Pyoderma gangrenosum:** Rare skin condition characterized by painful, rapidly progressing ulcers, often associated with immune system abnormalities, not caused by infection.

**Sarcoidosis:** A multisystem inflammatory disorder characterised by the presence of granulomas in various organs and tissues throughout the body.

***Staphylococcus***: A genus of bacteria that commonly inhabit the skin, mucous membranes, and respiratory and gastrointestinal tracts.

***Streptococcus***: A diverse group of bacteria, some of which are part of the normal microbiota, while others can cause a wide range of infections.

**Syncope:** A sudden and temporary loss of consciousness and posture, usually caused by a temporary decrease in blood flow to the brain.

**Temporomandibular joint dysfunction:** A condition affecting the jaw joints and surrounding muscles, characterized by symptoms such as jaw pain, limited movement, clicking sounds, and muscle tension.

**Tongue fissure**: Grooves or furrows on the tongue’s surface, typically harmless and benign.

**Toxic shock syndrome**: A rare and potentially life-threatening medical condition caused by the release of toxins, often associated with certain bacterial infections.

**Uveitis:** Inflammation of the uvea, the middle layer of the eye.

**Vitiligo:** A chronic skin disorder characterised by the loss of pigmentation in specific areas of the skin, resulting in white patches or depigmented spots due to the destruction of melanocytes, the cells responsible for skin colouration.

## APPENDIX 2: Search Results

### Q1a: Tattooing

Medline (Ovid) 01.08.2023

| # | Query | Results from 1 Aug 2023 |
| --- | --- | --- |
| 1 | exp Tattooing/ | 4,103 |
| 2 | (tattoo* or body ink or body art or hand poke or hand poking or stick-and-poke).tw. | 5,924 |
| 3 | (microblading or microstroking).tw. | 19 |
| 4 | (eyebrow* adj2 (embroid* or feather* or enhance*)).tw. | 9 |
| 5 | ((permanent or semi-permanent) adj2 (makeup or make-up or cosmetic* or skin colo?ring or skin colo?ration)).tw. | 151 |
| 6 | micropigment*.tw. | 57 |
| 7 | dermal pigmentation.tw. | 54 |
| 8 | 1 or 2 or 3 or 4 or 5 or 6 or 7 | 6,735 |
| 9 | (ae or co or in or to or bl or mo).fs. | 6,447,430 |
| 10 | (adverse effect* or adverse event* or adverse outcome* or adverse reaction* or complication* or harm* or risk* or hazard* or safe* or side effect* or tolera* or toxic* or undesirable effect* or undesirable event* or unexpected effect* or unexpected event*).tw. | 6,000,253 |
| 11 | (injur* or infect* or allerg* or anaphylax* or contra* or sepsis or septi* or inflam* or bacteria* or microb* or hygien* or granulom* or xanthom* or lichenoid* or streptococci or staphylococci or pseudomonas or mycobacteri* or hepatitis* or tuberculosis or syphilis or HIV or bleeding or scar* or keloid* or cicatri* or fung* or viral*).tw. | 7,603,873 |
| 12 | exp Risk Assessment/ | 310,876 |
| 13 | bacterial infections/ | 74,916 |
| 14 | exp Sepsis/ | 141,719 |
| 15 | exp Blood-Borne Infections/ | 427,194 |
| 16 | exp Methicillin-Resistant Staphylococcus aureus/ | 19,621 |
| 17 | Mycobacterium/ or Lichenoid Eruptions/ or Pseudomonas/ or Granuloma/ or Xanthomatosis/ | 65,704 |
| 18 | Streptococcal infections/ | 35,044 |
| 19 | Staphylococcal infections/ or Staphylococcal skin infections/ | 66,949 |
| 20 | Hepatitis/ or exp hepatitis, viral, human/ | 170,647 |
| 21 | Tuberculosis/ or tuberculosis, cutaneous/ | 115,504 |
| 22 | Syphilis/ or Syphilis, Cutaneous/ | 24,240 |
| 23 | exp HIV/ | 107,665 |
| 24 | cicatrix/ or keloid/ | 29,650 |
| 25 | 9 or 10 or 11 or 12 or 13 or 14 or 15 or 16 or 17 or 18 or 19 or 20 or 21 or 22 or 23 or 24 | 15,083,997 |
| 26 | exp "Systematic Review"/ | 234,581 |
| 27 | exp Meta-Analysis/ | 184,981 |
| 28 | exp Systematic Reviews as Topic/ | 11,052 |
| 29 | exp Meta-Analysis as Topic/ | 27,353 |
| 30 | (systematic review* or meta-analysis or review*).pt. | 3,356,147 |
| 31 | (systematic adj3 (review* or overview*)).ti,ab,kf. | 312,677 |
| 32 | (quantitative adj3 (review* or overview* or synthes*)).ti,ab,kf. | 8,458 |
| 33 | (meta-analy* or metaanaly* or meta-synthes#s or metasynthes#s).tw. | 277,778 |
| 34 | rapid review*.ti,ab,kf. | 2,014 |
| 35 | 26 or 27 or 28 or 29 or 30 or 31 or 32 or 33 or 34 | 3,455,925 |
| 36 | 8 and 25 and 35 | 570 |
| 37 | limit 36 to (english language and yr="2015 -Current") | **236** |

#### EMBASE (Ovid) 01.08.2023

| # | Query | Results from 1 Aug 2023 |
| --- | --- | --- |
| 1 | exp tattooing/ | 4,098 |
| 2 | (tattoo* or body ink or body art or hand poke or hand poking or stick-and-poke).tw. | 8,771 |
| 3 | (microblading or microstroking).tw. | 21 |
| 4 | (eyebrow* adj2 (embroid* or feather* or enhance*)).tw. | 11 |
| 5 | ((permanent or semi-permanent) adj2 (makeup or make-up or cosmetic* or skin colo?ring or skin colo?ration)).tw. | 216 |
| 6 | micropigment*.tw. | 78 |
| 7 | dermal pigmentation.tw. | 91 |
| 8 | 1 or 2 or 3 or 4 or 5 or 6 or 7 | 9,898 |
| 9 | (co or si).fs. | 2,855,329 |
| 10 | (adverse effect* or adverse event* or adverse outcome* or adverse reaction* or complication* or harm* or risk* or hazard* or safe* or side effect* or tolera* or toxic* or undesirable effect* or undesirable event* or unexpected effect* or unexpected event*).tw. | 8,639,309 |
| 11 | (injur* or infect* or allerg* or anaphylax* or contra* or sepsis or septi* or inflam* or bacteria* or microb* or hygien* or granulom* or xanthom* or lichenoid* or streptococci or staphylococci or pseudomonas or mycobacteri* or hepatitis* or tuberculosis or syphilis or HIV or bleeding or scar* or keloid* or cicatri* or fung* or viral*).tw. | 10,136,128 |
| 12 | exp risk assessment/ | 737,099 |
| 13 | exp adverse event/ | 997,582 |
| 14 | exp bacterial infection/ | 1,158,817 |
| 15 | exp sepsis/ | 347,731 |
| 16 | exp bloodstream infection/ | 18,583 |
| 17 | exp methicillin resistant Staphylococcus aureus/ | 56,389 |
| 18 | Mycobacterium/ or Lichenoid Eruption/ or Pseudomonas/ or Granuloma/ or Xanthomatosis/ | 115,389 |
| 19 | Streptococcus infection/ | 27,728 |
| 20 | Staphylococcus infection/ | 36,437 |
| 21 | exp hepatitis/ | 384,417 |
| 22 | tuberculosis/ | 174,710 |
| 23 | cutaneous syphilis/ or syphilis/ | 44,997 |
| 24 | exp Human immunodeficiency virus/ | 220,415 |
| 25 | exp scar/ | 112,223 |
| 26 | 9 or 10 or 11 or 12 or 13 or 14 or 15 or 16 or 17 or 18 or 19 or 20 or 21 or 22 or 23 or 24 or 25 | 17,794,402 |
| 27 | exp "systematic review"/ | 423,648 |
| 28 | exp meta analysis/ | 288,337 |
| 29 | (systematic review* or meta-analysis or review*).pt. | 3,149,146 |
| 30 | (systematic adj3 (review* or overview*)).ti,ab,kf. | 381,465 |
| 31 | (quantitative adj3 (review* or overview* or synthes*)).ti,ab,kf. | 9,792 |
| 32 | (meta-analy* or metaanaly* or meta-synthes#s or metasynthes#s).tw. | 353,228 |
| 33 | rapid review*.ti,ab,kf. | 2,421 |
| 34 | 27 or 28 or 29 or 30 or 31 or 32 or 33 | 3,571,527 |
| 35 | 8 and 26 and 34 | 598 |
| 36 | limit 35 to (english and yr="2015 -Current") | 229 |
| 37 | conference abstract.pt. | 4,843,274 |
| 38 | 36 not 37 | **215** |

#### Ovid EMCARE 01.08.2023

| # | Query | Results from 1 Aug 2023 |
| --- | --- | --- |
| 1 | exp tattooing/ | 584 |
| 2 | (tattoo* or body ink or body art or hand poke or hand poking or stick-and-poke).tw. | 1,825 |
| 3 | (microblading or microstroking).tw. | 7 |
| 4 | (eyebrow* adj2 (embroid* or feather* or enhance*)).tw. | 3 |
| 5 | ((permanent or semi-permanent) adj2 (makeup or make-up or cosmetic* or skin colo?ring or skin colo?ration)).tw. | 36 |
| 6 | micropigment*.tw. | 12 |
| 7 | dermal pigmentation.tw. | 10 |
| 8 | 1 or 2 or 3 or 4 or 5 or 6 or 7 | 1,992 |
| 9 | (adverse effect* or adverse event* or adverse outcome* or adverse reaction* or complication* or harm* or risk* or hazard* or safe* or side effect* or tolera* or toxic* or undesirable effect* or undesirable event* or unexpected effect* or unexpected event*).tw. | 1,990,510 |
| 10 | (injur* or infect* or allerg* or anaphylax* or contra* or sepsis or septi* or inflam* or bacteria* or microb* or hygien* or granulom* or xanthom* or lichenoid* or streptococci or staphylococci or pseudomonas or mycobacteri* or hepatitis* or tuberculosis or syphilis or HIV or bleeding or scar* or keloid* or cicatri* or fung* or viral*).tw. | 1,732,403 |
| 11 | exp risk assessment/ | 154,500 |
| 12 | exp adverse event/ | 143,131 |
| 13 | exp bacterial infection/ | 118,694 |
| 14 | exp sepsis/ | 53,567 |
| 15 | exp bloodstream infection/ | 2,787 |
| 16 | exp methicillin resistant Staphylococcus aureus/ | 9,790 |
| 17 | lichenoid eruption/ or pseudomonas/ or granuloma/ or xanthomatosis/ | 6,833 |
| 18 | exp Streptococcus infection/ | 8,292 |
| 19 | exp Staphylococcus infection/ | 7,487 |
| 20 | exp hepatitis/ | 34,679 |
| 21 | tuberculosis/ | 19,918 |
| 22 | syphilis/ | 4,607 |
| 23 | exp Human immunodeficiency virus/ | 55,245 |
| 24 | exp scar/ | 16,613 |
| 25 | 9 or 10 or 11 or 12 or 13 or 14 or 15 or 16 or 17 or 18 or 19 or 20 or 21 or 22 or 23 or 24 | 3,237,987 |
| 26 | exp "systematic review"/ | 120,894 |
| 27 | exp meta analysis/ | 58,543 |
| 28 | (systematic review* or meta-analysis or review*).pt. | 845,073 |
| 29 | (systematic adj3 (review* or overview*)).ti,ab,kf. | 159,122 |
| 30 | (quantitative adj3 (review* or overview* or synthes*)).ti,ab,kf. | 4,273 |
| 31 | (meta-analy* or metaanaly* or meta-synthes#s or metasynthes#s).tw. | 122,834 |
| 32 | rapid review*.ti,ab,kf. | 1,222 |
| 33 | 26 or 27 or 28 or 29 or 30 or 31 or 32 | 968,082 |
| 34 | 8 and 25 and 33 | 176 |
| 35 | limit 34 to (english language and yr="2015 -Current") | 66 |

#### AMED (Ovid) 01.08.23

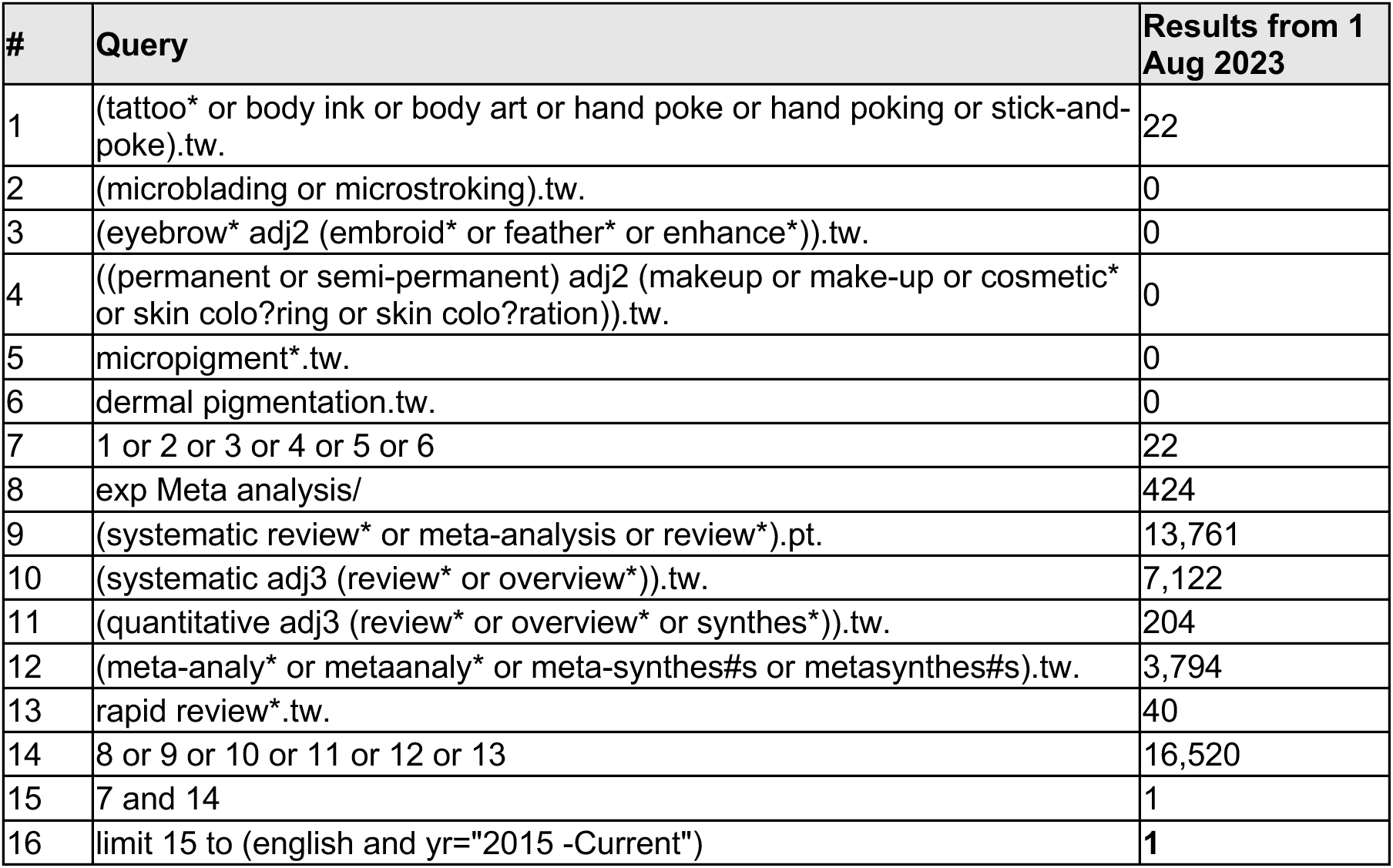

#### CINAHL (EBSCO) 01.08.2023

| # | Query | Results from 1 Aug 2023 |
| --- | --- | --- |
| 1 | (MH "Tattooing") | 1,357 |
| 2 | TI (tattoo* or "body ink" or "body art" or "hand poke" or "hand poking" or stick-and-poke) OR AB (tattoo* or "body ink" or "body art" or "hand poke" or "hand poking" or stick-and-poke) | 1,678 |
| 3 | TI (microblading or microstroking) OR AB (microblading or microstroking) | 9 |
| 4 | TI ( eyebrow* N2 (embroid* or feather* or enhance*) OR AB ( eyebrow* N2 (embroid* or feather* or enhance*) | 1 |
| 5 | TI ( ( permanent or semi-permanent) N2 (makeup or make-up or cosmetic* or skin colo#ring or skin colo#ration) ) OR AB ( ( permanent or semi-permanent) N2 (makeup or make-up or cosmetic* or skin colo#ring or skin colo#ration) ) ) | 32 |
| 6 | TI micropigment* OR AB micropigment* | 12 |
| 7 | TI "dermal pigmentation" OR AB "dermal pigmentation" | 6 |
| 8 | OR 1-7 | 2,042 |
| 9 | MW ae or mo or rf or bl or co or pc | 1,882,053 |
| 10 | TI ( "adverse effect*" or "adverse event*" or "adverse outcome*" or "adverse reaction*" or complication* or harm* or risk* or hazard* or safe* or "side effect*" or tolera* or toxic* or "undesirable effect*" or "undesirable event*" or "unexpected effect*" or "unexpected event*" ) OR AB ("adverse effect*" or "adverse event*" or "adverse outcome*" or "adverse reaction*" or complication* or harm* or risk* or hazard* or safe* or "side effect*" or tolera* or toxic* or "undesirable effect*" or "undesirable event*" or "unexpected effect*" or "unexpected event*" ) | 1,558,106 |
| 11 | TI ( injur* or infect* or allerg* or anaphylax* or contra* or sepsis or septi* or inflam* or bacteria* or microb* or hygien* or granulom* or xanthom* or | 1,211,678 |
|  | lichenoid* or streptococci or staphylococci or pseudomonas or mycobacteri* or hepatitis* or tuberculosis or syphilis or HIV or bleeding or scar* or keloid* or cicatri* or fung* or viral* ) OR AB ( injur* or infect* or allerg* or anaphylax* or contra* or sepsis or septi* or inflam* or bacteria* or microb* or hygien* or granulom* or xanthom* or lichenoid* or streptococci or staphylococci or pseudomonas or mycobacteri* or hepatitis* or tuberculosis or syphilis or HIV or bleeding or scar* or keloid* or cicatri* or fung* or viral* ) |  |
| 12 | (MH "Risk Assessment") | 150,332 |
| 13 | (MH "Bacterial Infections+") | 133,935 |
| 14 | (MH "Sepsis+") | 31,807 |
| 15 | (MH "Methicillin-Resistant Staphylococcus Aureus") | 5,872 |
| 16 | (MH "Mycobacterium+") | 6,055 |
| 17 | (MH "Lichenoid Eruptions+") | 1,225 |
| 18 | (MH "Pseudomonas Infections") | 2,961 |
| 19 | (MH "Granuloma") | 3,391 |
| 20 | (MH "Lipid Metabolism, Inborn Errors") | 4,171 |
| 21 | (MH "Streptococcal Infections+") | 11,679 |
| 22 | (MH "Hepatitis+") | 31,212 |
| 23 | (MH "Tuberculosis+") | 25,755 |
| 24 | (MH "Syphilis+") | 4,757 |
| 25 | (MH "Human Immunodeficiency Virus+") | 11,209 |
| 26 | (MH "Cicatrix+") | 6,909 |
| 27 | (MH "Keloid") | 642 |
| 28 | OR 9-27 | 3,321,671 |
| 29 | (MH "Systematic Review") | 122,674 |
| 30 | (MH "Meta Analysis") | 70,113 |
| 31 | PT "systematic review*" or "meta-analysis" or review* | 504,595 |
| 32 | TI ( systematic N3 (review* or overview*) ) OR AB ( systematic N3 (review* or overview*) ) | 151,714 |
| 33 | TI ( quantitative N3 (review* or overview* or syntheses*) ) OR AB ( quantitative N3 (review* or overview* or syntheses*) ) | 3,871 |
| 34 | TI ( meta-analy* or metaanaly* or meta-synthes?s or metasynthes?s ) OR AB ( meta-analy* or metaanaly* or meta-synthes?s or metasynthes?s ) | 111,207 |
| 35 | TI "rapid review*" OR AB "rapid review*" | 939 |
| 36 | OR 29-35 | 606,362 |
| 37 | 8 AND 28 AND 36 Limited 20150101 – 20230831; English Language | **38** |

#### COCHRANE LIBRARY 01.08.2023

| # | Query | Results from 1 Aug 2023 |
| --- | --- | --- |
| 1 | MeSH descriptor: [Tattooing] explode all trees | 51 |
| 2 | (tattoo* or "body ink" or "body art" or "hand poke" or "hand poking" or "stick and poke").ti,ab,kw | 278 |
| 3 | (microblading or microstroking).ti,ab,kw | 3 |
| 4 | (eyebrow* NEAR (embroid* or feather* or enhance*)).ti,ab,kw | 7 |
| 5 | ((permanent or semi-permanent) NEAR (makeup or make-up or cosmetic* or "skin colouring" or "skin colouration").ti,ab,kw (word variations have been searched) | 8 |
| 6 | (micropigment*).ti,ab,kw | 4 |
| 7 | ("dermal pigmentation").ti,ab,kw | 5 |
| 8 | OR 1-7 | 301 (296 trials, 5 reviews) |

#### EPISTEMONIKOS 01.08.2023

| # | Query | Results from 1 Aug 2023 |
| --- | --- | --- |
| 1 | (title:(tattoo* OR "body ink" OR "body art" OR "hand poke" OR "hand poking" OR "stick-and-poke" OR microblading OR microstroking) OR abstract:(tattoo* OR "body ink" OR "body art" OR "hand poke" OR "hand poking" OR "stick-and-poke" OR microblading OR microstroking)) | 607 |
| 2 | (title:((eyebrow*) AND (embroid* OR feather* OR enhance*)) OR abstract:((eyebrow*) AND (embroid* OR feather* OR enhance*))) | 8 |
| 3 | (title:((permanent OR semi-permanent OR "semi permanent") AND (makeup OR make-up OR "make up" OR cosmetic* OR "skin colouring" OR "skin coloring" OR "skin colouration" OR "skin coloration")) OR abstract:((permanent OR semi-permanent OR "semi permanent") AND (makeup OR make-up OR "make up" OR cosmetic* OR "skin colouring" OR "skin coloring" OR "skin colouration" OR "skin coloration"))) | 177 |
| 4 | (title:(micropigment* OR "dermal pigmentation") OR abstract:(micropigment* OR "dermal pigmentation")) | 5 |
| 5 | OR 1-5 | 790 |
| 6 | 5 Limited to Systematic Review and English Language | 86 |

#### Total References Added to Endnote

| Database | References |
| --- | --- |
| Medline (Ovid) | 236 |
| EMBASE (Ovid) | 215 |
| Ovid EMCARE | 66 |
| AMED (Ovid) | 1 |
| CINAHL (EBSCO) | 38 |
| Cochrane Library (Reviews) | 5 |
| Epistemonikos | 86 |
| **TOTAL** | **647** |
| Duplicates Identified in Endnote | 231 |
| **New TOTAL** | **416** |

### Q1b: Semi-permanent make-up

#### MEDLINE (Ovid) 05.09.2023

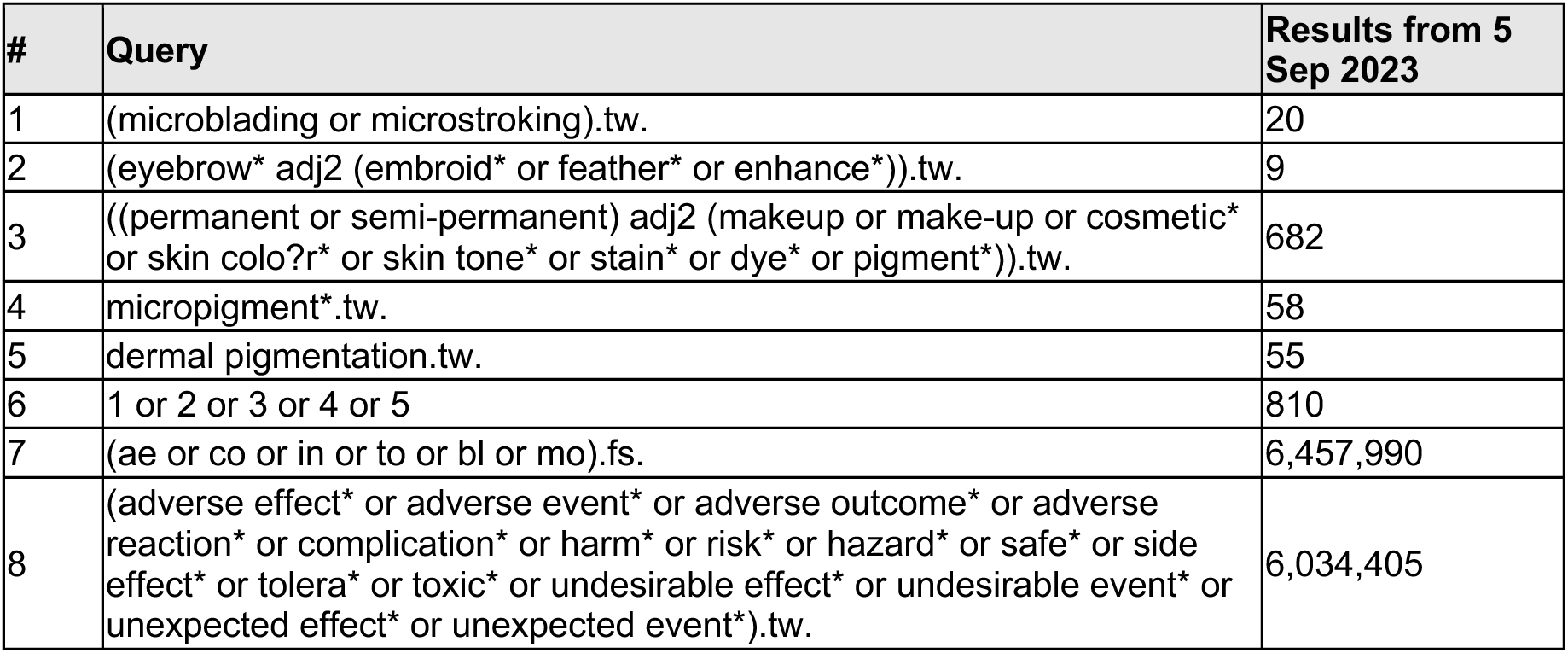
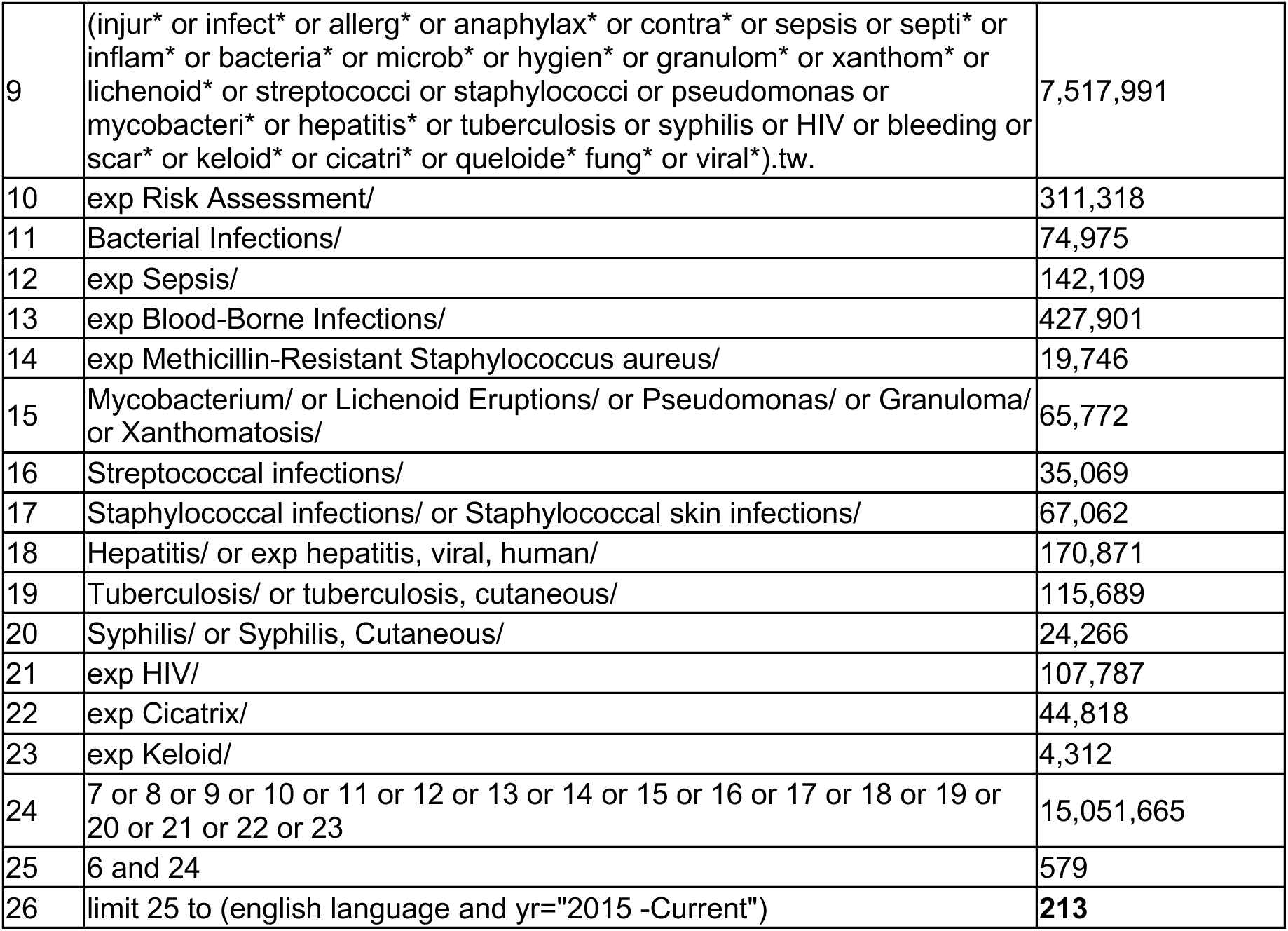

Q1b: Semi-permanent make-up MEDLINE (Ovid) 05.09.2023
| # | Query | Results from 5 Sep 2023 |
| --- | --- | --- |
| 1 | (microblading or microstroking).tw. | 20 |
| 2 | (eyebrow* adj2 (embroid* or feather* or enhance*)).tw. | 9 |
| 3 | ((permanent or semi-permanent) adj2 (makeup or make-up or cosmetic* or skin colo?r* or skin tone* or stain* or dye* or pigment*)).tw. | 682 |
| 4 | micropigment*.tw. | 58 |
| 5 | dermal pigmentation.tw. | 55 |
| 6 | 1 or 2 or 3 or 4 or 5 | 810 |
| 7 | (ae or co or in or to or bl or mo).fs. | 6,457,990 |
| 8 | (adverse effect* or adverse event* or adverse outcome* or adverse reaction* or complication* or harm* or risk* or hazard* or safe* or side effect* or tolera* or toxic* or undesirable effect* or undesirable event* or unexpected effect* or unexpected event*).tw. | 6,034,405 |
| 9 | (injur* or infect* or allerg* or anaphylax* or contra* or sepsis or septi* or inflam* or bacteria* or microb* or hygien* or granulom* or xanthom* or lichenoid* or streptococci or staphylococci or pseudomonas or mycobacteri* or hepatitis* or tuberculosis or syphilis or HIV or bleeding or scar* or keloid* or cicatri* or queloide* fung* or viral*).tw. | 7,517,991 |
| 10 | exp Risk Assessment/ | 311,318 |
| 11 | Bacterial Infections/ | 74,975 |
| 12 | exp Sepsis/ | 142,109 |
| 13 | exp Blood-Borne Infections/ | 427,901 |
| 14 | exp Methicillin-Resistant Staphylococcus aureus/ | 19,746 |
| 15 | Mycobacterium/ or Lichenoid Eruptions/ or Pseudomonas/ or Granuloma/ or Xanthomatosis/ | 65,772 |
| 16 | Streptococcal infections/ | 35,069 |
| 17 | Staphylococcal infections/ or Staphylococcal skin infections/ | 67,062 |
| 18 | Hepatitis/ or exp hepatitis, viral, human/ | 170,871 |
| 19 | Tuberculosis/ or tuberculosis, cutaneous/ | 115,689 |
| 20 | Syphilis/ or Syphilis, Cutaneous/ | 24,266 |
| 21 | exp HIV/ | 107,787 |
| 22 | exp Cicatrix/ | 44,818 |
| 23 | exp Keloid/ | 4,312 |
| 24 | 7 or 8 or 9 or 10 or 11 or 12 or 13 or 14 or 15 or 16 or 17 or 18 or 19 or 20 or 21 or 22 or 23 | 15,051,665 |
| 25 | 6 and 24 | 579 |
| 26 | limit 25 to (english language and yr="2015 -Current") | 213 |

#### EMBASE (Ovid) 05.09.2023

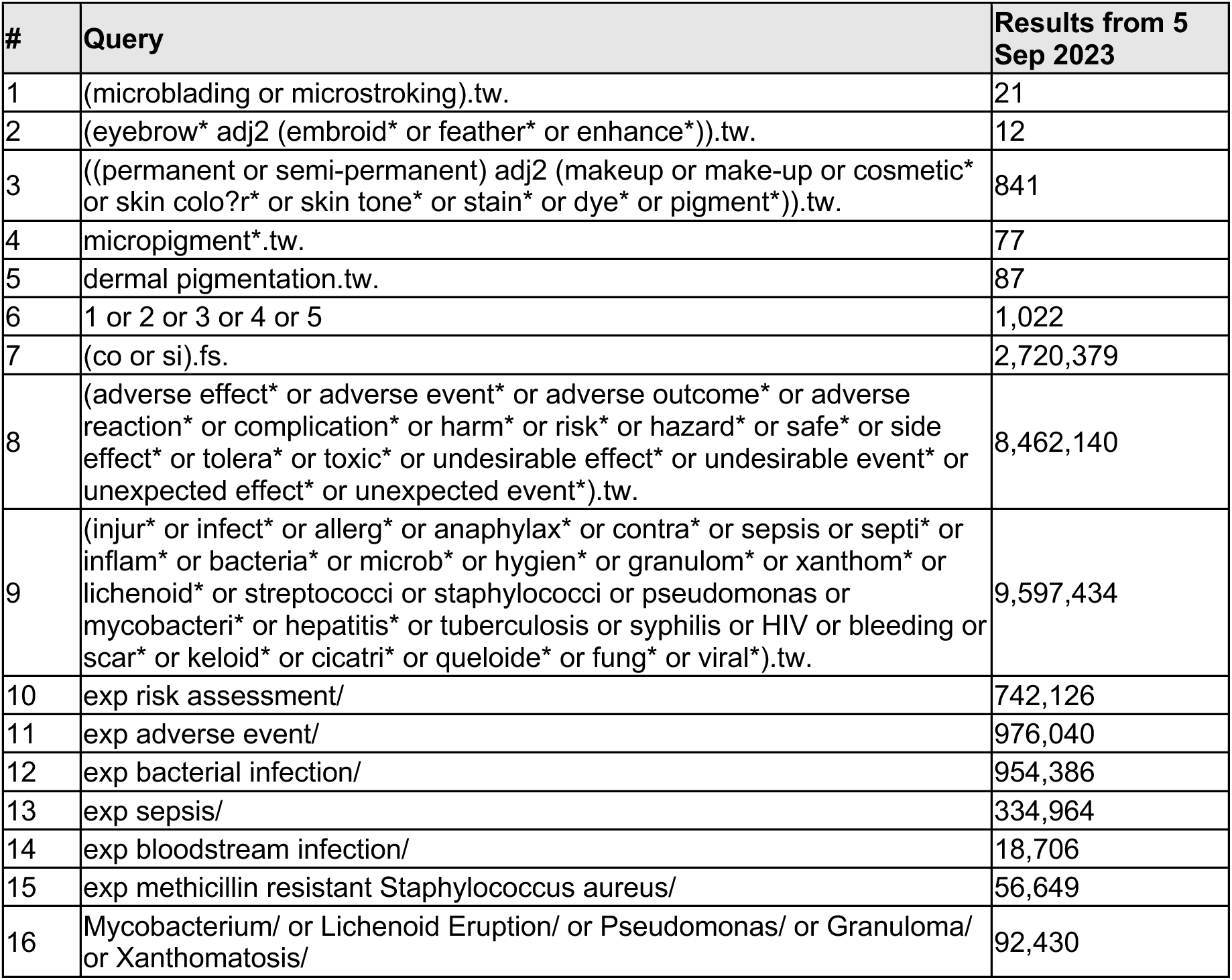
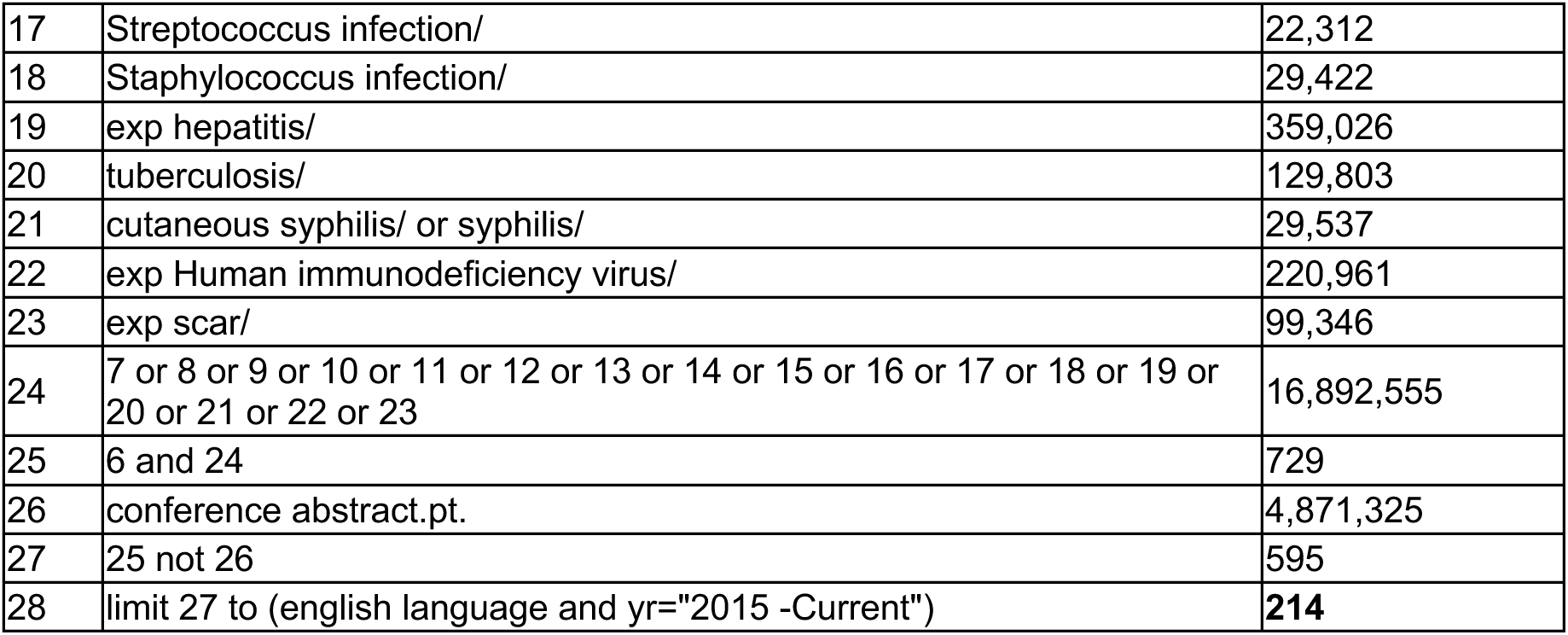

| # | Query | Results from 5 Sep 2023 |
| --- | --- | --- |
| 1 | (microblading or microstroking).tw. | 21 |
| 2 | (eyebrow* adj2 (embroid* or feather* or enhance*)).tw. | 12 |
| 3 | ((permanent or semi-permanent) adj2 (makeup or make-up or cosmetic* or skin colo?r* or skin tone* or stain* or dye* or pigment*)).tw. | 841 |
| 4 | micropigment*.tw. | 77 |
| 5 | dermal pigmentation.tw. | 87 |
| 6 | 1 or 2 or 3 or 4 or 5 | 1,022 |
| 7 | (co or si).fs. | 2,720,379 |
| 8 | (adverse effect* or adverse event* or adverse outcome* or adverse reaction* or complication* or harm* or risk* or hazard* or safe* or side effect* or tolera* or toxic* or undesirable effect* or undesirable event* or unexpected effect* or unexpected event*).tw. | 8,462,140 |
| 9 | (injur* or infect* or allerg* or anaphylax* or contra* or sepsis or septi* or inflam* or bacteria* or microb* or hygien* or granulom* or xanthom* or lichenoid* or streptococci or staphylococci or pseudomonas or mycobacteri* or hepatitis* or tuberculosis or syphilis or HIV or bleeding or scar* or keloid* or cicatri* or queloide* or fung* or viral*).tw. | 9,597,434 |
| 10 | exp risk assessment/ | 742,126 |
| 11 | exp adverse event/ | 976,040 |
| 12 | exp bacterial infection/ | 954,386 |
| 13 | exp sepsis/ | 334,964 |
| 14 | exp bloodstream infection/ | 18,706 |
| 15 | exp methicillin resistant Staphylococcus aureus/ | 56,649 |
| 16 | Mycobacterium/ or Lichenoid Eruption/ or Pseudomonas/ or Granuloma/ or Xanthomatosis/ | 92,430 |
| 17 | Streptococcus infection/ | 22,312 |
| 18 | Staphylococcus infection/ | 29,422 |
| 19 | exp hepatitis/ | 359,026 |
| 20 | tuberculosis/ | 129,803 |
| 21 | cutaneous syphilis/ or syphilis/ | 29,537 |
| 22 | exp Human immunodeficiency virus/ | 220,961 |
| 23 | exp scar/ | 99,346 |
| 24 | 7 or 8 or 9 or 10 or 11 or 12 or 13 or 14 or 15 or 16 or 17 or 18 or 19 or 20 or 21 or 22 or 23 | 16,892,555 |
| 25 | 6 and 24 | 729 |
| 26 | conference abstract.pt. | 4,871,325 |
| 27 | 25 not 26 | 595 |
| 28 | limit 27 to (english language and yr="2015 -Current") | **214** |

#### Ovid EMCARE 05.09.2023

| # | Query | Results from 5 Sep 2023 |
| --- | --- | --- |
| 1 | (microblading or microstroking).tw. | 7 |
| 2 | (eyebrow* adj2 (embroid* or feather* or enhance*)).tw. | 3 |
| 3 | ((permanent or semi-permanent) adj2 (makeup or make-up or cosmetic* or skin colo?r* or skin tone* or stain* or dye* or pigment*)).tw. | 111 |
| 4 | micropigment*.tw. | 12 |
| 5 | dermal pigmentation.tw. | 10 |
| 6 | 1 or 2 or 3 or 4 or 5 | 141 |
| 7 | (adverse effect* or adverse event* or adverse outcome* or adverse reaction* or complication* or harm* or risk* or hazard* or safe* or side effect* or tolera* or toxic* or undesirable effect* or undesirable event* or unexpected effect* or unexpected event*).tw. | 2,006,836 |
| 8 | (injur* or infect* or allerg* or anaphylax* or contra* or sepsis or septi* or inflam* or bacteria* or microb* or hygien* or granulom* or xanthom* or lichenoid* or streptococci or staphylococci or pseudomonas or mycobacteri* or hepatitis* or tuberculosis or syphilis or HIV or bleeding or scar* or keloid* or cicatri* or queloide* or fung* or viral*).tw. | 1,744,788 |
| 9 | exp risk assessment/ | 155,005 |
| 10 | exp adverse event/ | 143,308 |
| 11 | exp bacterial infection/ | 118,866 |
| 12 | exp sepsis/ | 53,602 |
| 13 | exp bloodstream infection/ | 2,796 |
| 14 | exp methicillin resistant Staphylococcus aureus/ | 9,798 |
| 15 | lichenoid eruption/ or pseudomonas/ or granuloma/ or xanthomatosis/ | 6,840 |
| 16 | exp Streptococcus infection/ | 8,294 |
| 17 | exp Staphylococcus infection/ | 7,490 |
| 18 | exp hepatitis/ | 34,728 |
| 19 | tuberculosis/ | 19,967 |
| 20 | syphilis/ | 4,622 |
| 21 | exp Human immunodeficiency virus/ | 55,306 |
| 22 | exp scar/ | 16,620 |
| 23 | exp keloid/ | 1,055 |
| 24 | 7 or 8 or 9 or 10 or 11 or 12 or 13 or 14 or 15 or 16 or 17 or 18 or 19 or 20 or 21 or 22 or 23 | 3,261,810 |
| 25 | 6 and 24 | 108 |
| 26 | limit 25 to (english language and yr="2015 -Current") | **51** |

#### AMED (Ovid) 05.09.2023

| # | Query | Results from 5 Sep 2023 |
| --- | --- | --- |
| 1 | (microblading or microstroking).tw. | 0 |
| 2 | (eyebrow* adj2 (embroid* or feather* or enhance*)).tw. | 0 |
| 3 | ((permanent or semi-permanent) adj2 (makeup or make-up or cosmetic* or skin colo?r* or skin tone* or stain* or dye* or pigment*)).tw. | 1 |
| 4 | micropigment*.tw. | 0 |
| 5 | dermal pigmentation.tw. | 0 |
| 6 | 1 or 2 or 3 or 4 or 5 | 1 |

#### CINAHL (EBSCO) 05.09.2023

| # | Query | Results from 5 Sep 2023 |
| --- | --- | --- |
| 1 | TI (microblading or microstroking) OR AB (microblading or microstroking) | 9 |
| 2 | TI ((eyebrow* N2 (embroid* or feather* or enhance*)) OR AB ((eyebrow* N2 (embroid* or feather* or enhance*)) | 1 |
| 3 | TI ((permanent or semi-permanent) N2 (makeup or make-up or cosmetic* or skin colo?r* or skin tone* or stain* or dye* or pigment*)) OR AB ((permanent or semi-permanent) N2 (makeup or make-up or cosmetic* or skin colo?r* or skin tone* or stain* or dye* or pigment*)) | 92 |
| 4 | TI micropigment* OR AB mocropigment* | 13 |
| 5 | TI "dermal pigmentation" OR AB "dermal pigmentation" | 6 |
| 6 | 1 or 2 or 3 or 4 or 5 | 119 |
| 7 | MW ae or mo or rf or bl or co or pc | 1,829,897 |
| 8 | TI ("adverse effect*" or "adverse event*" or "adverse outcome*" or "adverse reaction*" or complication* or harm* or risk* or hazard* or safe* or "side effect*" or tolera* or toxic* or "undesirable effect*" or "undesirable event*" or "unexpected effect*" or "unexpected event*") OR AB ("adverse effect*" or "adverse event*" or "adverse outcome*" or "adverse reaction*" or complication* or harm* or risk* or hazard* or safe* or "side effect*" or tolera* or toxic* or "undesirable effect*" or "undesirable event*" or "unexpected effect*" or "unexpected event*") | 1,567,613 |
| 9 | TI (injur* or infect* or allerg* or anaphylax* or contra* or sepsis or septi* or inflam* or bacteria* or microb* or hygien* or granulom* or xanthom* or lichenoid* or streptococci or staphylococci or pseudomonas or mycobacteri* or hepatitis* or tuberculosis or syphilis or HIV or bleeding or scar* or keloid* or cicatri* or queloide* or fung* or viral*) OR AB (injur* or infect* or allerg* or anaphylax* or contra* or sepsis or septi* or inflam* or bacteria* or microb* or hygien* or granulom* or xanthom* or lichenoid* or streptococci or staphylococci or pseudomonas or mycobacteri* or hepatitis* or tuberculosis or syphilis or HIV or bleeding or scar* or keloid* or cicatri* or queloide* or fung* or viral*) | 1,218,441 |
| 10 | (MH "Risk Assessment") | 152,447 |
| 11 | (MH "Bacterial Infections") | 10,920 |
| 12 | (MH "Sepsis") | 19,450 |
| 13 | (MH "Methicillin-Resistant Staphylococcus Aureus") | 5,884 |
| 14 | (MH "Lichenoid Eruptions") | 360 |
| 15 | (MH "Pseudomonas Infections") | 2,968 |
| 16 | (MH "Granuloma") | 3,393 |
| 17 | (MH "Lipid Metabolism, Inborn Errors") | 4,179 |
| 18 | (MH "Streptococcal Infections") | 5,550 |
| 19 | (MH "Hepatitis") | 3.037 |
| 20 | (MH "Tuberculosis") | 17,561 |
| 21 | (MH "Syphilis") | 4,181 |
| 22 | (MH "Human Immunodeficiency Virus") | 5,680 |
| 23 | (MH "Cicatrix+") | 6,924 |
| 24 | (MH "Keloid") | 643 |
| 25 | OR 7-24 | 3,318,333 |
| 26 | 6 AND 25 | 89 |
| 27 | 26 Limited to English Language and 2015 – 31082023 | **46** |

#### Total References Added to Endnote

| Database | References |
| --- | --- |
| Medline (Ovid) | 213 |
| EMBASE (Ovid) | 214 |
| Ovid EMCARE | 51 |
| AMED (Ovid) | 1 |
| CINAHL (EBSCO) | 46 |
| **TOTAL** | **525** |
| Duplicates Identified in Endnote | 251 |
| **New TOTAL** | **274** |

### Q2: Body Piercing

#### Medline (Ovid) 02.08.2023

Q2: Body Piercing Medline (Ovid) 02.08.2023
| # | Query | Results from 2 Aug 2023 |
| --- | --- | --- |
| 1 | exp Body Piercing/ | 670 |
| 2 | piercing*.tw. | 3,163 |
| 3 | 1 or 2 | 3,291 |
| 4 | (ae or co or in or to or bl or mo).fs. | 6,447,679 |
| 5 | (adverse effect* or adverse event* or adverse outcome* or adverse reaction* or complication* or harm* or risk* or hazard* or safe* or side effect* or tolera* or toxic* or undesirable effect* or undesirable event* or unexpected effect* or unexpected event*).tw. | 6,001,307 |
| 6 | (injur* or infect* or allerg* or anaphylax* or contra* or sepsis or septi* or inflam* or bacteria* or microb* or hygien* or granulom* or xanthom* or lichenoid* or streptococci or staphylococci or pseudomonas or mycobacteri* or hepatitis* or tuberculosis or syphilis or HIV or bleeding or scar* or keloid* or cicatri* or fung* or viral*).tw. | 7,604,837 |
| 7 | exp Risk Assessment/ | 310,881 |
| 8 | bacterial infections/ | 74,918 |
| 9 | exp Sepsis/ | 141,729 |
| 10 | exp Blood-Borne Infections/ | 427,206 |
| 11 | exp Methicillin-Resistant Staphylococcus aureus/ | 19,622 |
| 12 | Mycobacterium/ or Lichenoid Eruptions/ or Pseudomonas/ or Granuloma/ or Xanthomatosis/ | 65,705 |
| 13 | Streptococcal infections/ | 35,045 |
| 14 | Staphylococcal infections/ or Staphylococcal skin infections/ | 66,951 |
| 15 | Hepatitis/ or exp hepatitis, viral, human/ | 170,655 |
| 16 | Tuberculosis/ or tuberculosis, cutaneous/ | 115,504 |
| 17 | Syphilis/ or Syphilis, Cutaneous/ | 24,240 |
| 18 | exp HIV/ | 107,668 |
| 19 | cicatrix/ or keloid/ | 29,651 |
| 20 | 4 or 5 or 6 or 7 or 8 or 9 or 10 or 11 or 12 or 13 or 14 or 15 or 16 or 17 or 18 or 19 | 15,085,768 |
| 21 | exp "Systematic Review"/ | 234,624 |
| 22 | exp Meta-Analysis/ | 185,019 |
| 23 | exp Systematic Reviews as Topic/ | 11,055 |
| 24 | exp Meta-Analysis as Topic/ | 27,362 |
| 25 | (systematic review* or meta-analysis or review*).pt. | 3,356,535 |
| 26 | (systematic adj3 (review* or overview*)).ti,ab,kf. | 312,801 |
| 27 | (quantitative adj3 (review* or overview* or syntheses*)).ti,ab,kf. | 8,459 |
| 28 | (meta-analy* or metaanaly* or meta-synthes#s or metasyntes#s).tw. | 277,879 |
| 29 | rapid review*.ti,ab,kf. | 2,015 |
| 30 | 21 or 22 or 23 or 24 or 25 or 26 or 27 or 28 or 29 | 3,456,366 |
| 31 | 3 and 20 and 30 | 291 |
| 32 | limit 31 to (english language and yr="2015 -Current") | 89 |

#### EMBASE (Ovid) 02.08.2023

| # | Query | Results from 2 Aug 2023 |
| --- | --- | --- |
| 1 | exp Body Piercing/ | 1,247 |
| 2 | piercing*.tw. | 4,190 |
| 3 | 1 or 2 | 4,550 |
| 4 | (co or si).fs. | 2,855,329 |
| 5 | (adverse effect* or adverse event* or adverse outcome* or adverse reaction* or complication* or harm* or risk* or hazard* or safe* or side effect* or tolera* or toxic* or undesirable effect* or undesirable event* or unexpected effect* or unexpected event*).tw. | 8,641,323 |
| 6 | (injur* or infect* or allerg* or anaphylax* or contra* or sepsis or sepsi* or inflam* or bacteria* or microb* or hygien* or granulom* or xanthom* or lichenoid* or streptococci or staphylococci or pseudomonas or mycobacteri* or hepatitis* or tuberculosis or syphilis or HIV or bleeding or scar* or keloid* or cicatri* or fung* or viral*).tw. | 10,138,096 |
| 7 | exp risk assessment/ | 737,370 |
| 8 | exp adverse event/ | 997,723 |
| 9 | exp bacterial infection/ | 1,158,933 |
| 10 | exp sepsis/ | 347,774 |
| 11 | exp bloodstream infection/ | 18,590 |
| 12 | exp methicillin resistant Staphylococcus aureus/ | 56,403 |
| 13 | Mycobacterium/ or Lichenoid Eruption/ or Pseudomonas/ or Granuloma/ or Xanthomatosis/ | 115,398 |
| 14 | Streptococcus infection/ | 27,728 |
| 15 | Staphylococcus infection/ | 36,437 |
| 16 | exp hepatitis/ | 384,462 |
| 17 | tuberculosis/ | 174,733 |
| 18 | cutaneous syphilis/ or syphilis/ | 45,003 |
| 19 | exp Human immunodeficiency virus/ | 220,438 |
| 20 | exp scar/ | 112,238 |
| 21 | 4 or 5 or 6 or 7 or 8 or 9 or 10 or 11 or 12 or 13 or 14 or 15 or 16 or 17 or 18 or 19 or 20 | 17,797,748 |
| 22 | exp "systematic review"/ | 423,942 |
| 23 | exp meta analysis/ | 288,470 |
| 24 | (systematic review* or meta-analysis or review*).pt. | 3,149,857 |
| 25 | (systematic adj3 (review* or overview*)).ti,ab,kf. | 381,679 |
| 26 | (quantitative adj3 (review* or overview* or syntheses*)).ti,ab,kf. | 9,795 |
| 27 | (meta-analy* or metaanaly* or meta-synthes#s or metasyntes#s).tw. | 353,404 |
| 28 | rapid review*.ti,ab,kf. | 2,422 |
| 29 | 22 or 23 or 24 or 25 or 26 or 27 or 28 | 3,572,430 |
| 30 | 3 and 21 and 29 | 310 |
| 31 | limit 30 to (english and yr="2015 -Current") | 97 |
| 32 | conference abstract.pt. | 4,843,274 |
| 33 | 31 not 32 | **89** |

#### Ovid EMCARE 02.08.2023

| # | Query | Results from 2 Aug 2023 |
| --- | --- | --- |
| 1 | exp Body Piercing/ | 379 |
| 2 | piercing*.tw. | 1,073 |
| 3 | 1 or 2 | 1,138 |
| 4 | (adverse effect* or adverse event* or adverse outcome* or adverse reaction* or complication* or harm* or risk* or hazard* or safe* or side effect* or tolera* or toxic* or undesirable effect* or undesirable event* or unexpected effect* or unexpected event*).tw. | 1,990,510 |
| 5 | (injur* or infect* or allerg* or anaphylax* or contra* or sepsis or septi* or inflam* or bacteria* or microb* or hygien* or granulom* or xanthom* or lichenoid* or streptococci or staphylococci or pseudomonas or mycobacteri* or hepatitis* or tuberculosis or syphilis or HIV or bleeding or scar* or keloid* or cicatri* or fung* or viral*).tw. | 1,732,403 |
| 6 | exp risk assessment/ | 154,500 |
| 7 | exp adverse event/ | 143,131 |
| 8 | exp bacterial infection/ | 118,694 |
| 9 | exp sepsis/ | 53,567 |
| 10 | exp bloodstream infection/ | 2,787 |
| 11 | exp methicillin resistant Staphylococcus aureus/ | 9,790 |
| 12 | lichenoid eruption/ or pseudomonas/ or granuloma/ or xanthomatosis/ | 6,833 |
| 13 | exp Streptococcus infection/ | 8,292 |
| 14 | exp Staphylococcus infection/ | 7,487 |
| 15 | exp hepatitis/ | 34,679 |
| 16 | tuberculosis/ | 19,918 |
| 17 | syphilis/ | 4,607 |
| 18 | exp Human immunodeficiency virus/ | 55,245 |
| 19 | exp scar/ | 16,613 |
| 20 | 4 or 5 or 6 or 7 or 8 or 9 or 10 or 11 or 12 or 13 or 14 or 15 or 16 or 17 or 18 or 19 | 3,237,987 |
| 21 | exp "systematic review"/ | 120,894 |
| 22 | exp meta analysis/ | 58,543 |
| 23 | (systematic review* or meta-analysis or review*).pt. | 845,073 |
| 24 | (systematic adj3 (review* or overview*)).ti,ab,kf. | 159,122 |
| 25 | (quantitative adj3 (review* or overview* or synthes*)).ti,ab,kf. | 4,273 |
| 26 | (meta-analy* or metaanaly* or meta-synthes#s or metasyntes#s).tw. | 122,834 |
| 27 | rapid review*.ti,ab,kf. | 1,222 |
| 28 | 21 or 22 or 23 or 24 or 25 or 26 or 27 | 968,082 |
| 29 | 3 and 20 and 28 | 110 |
| 30 | limit 29 to (english language and yr="2015 -Current") | **31** |

#### AMED (Ovid) 02.08.2023

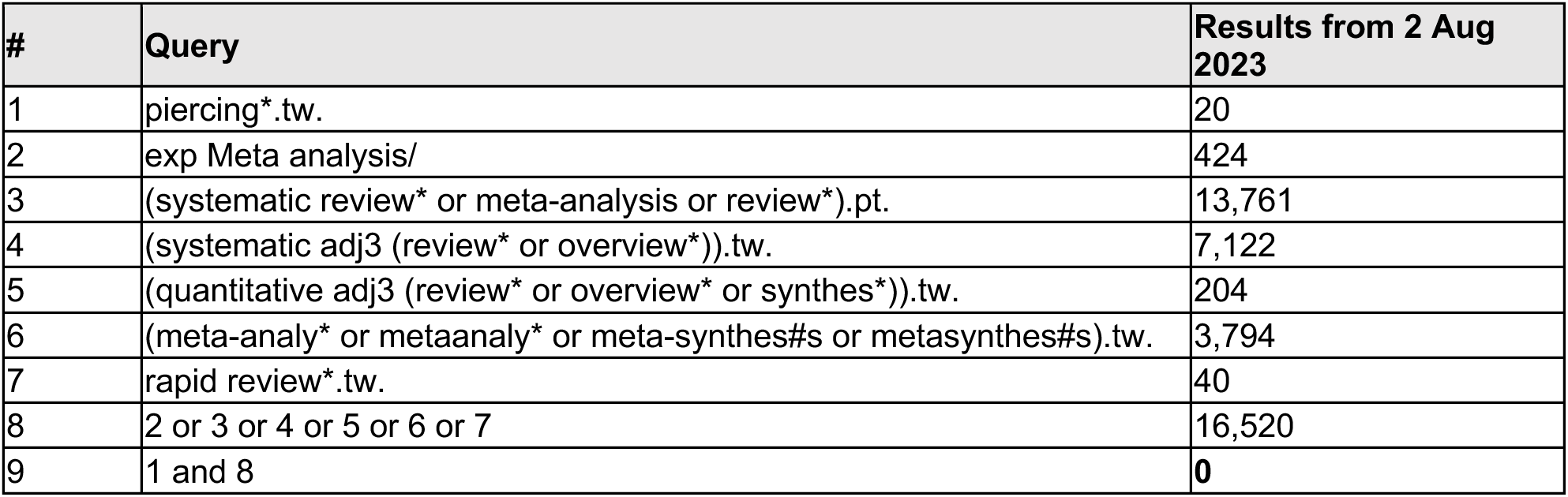

| # | Query | Results from 2 Aug 2023 |
| --- | --- | --- |
| 1 | piercing*.tw. | 20 |
| 2 | exp Meta analysis/ | 424 |
| 3 | (systematic review* or meta-analysis or review*).pt. | 13,761 |
| 4 | (systematic adj3 (review* or overview*).tw. | 7,122 |
| 5 | (quantitative adj3 (review* or overview* or syntheses*).tw. | 204 |
| 6 | (meta-analy* or metaanaly* or meta-synthes#s or metasynthes#s).tw. | 3,794 |
| 7 | rapid review*.tw. | 40 |
| 8 | 2 or 3 or 4 or 5 or 6 or 7 | 16,520 |
| 9 | 1 and 8 | 0 |

#### CINAHL (EBSCO) 02.08.2023

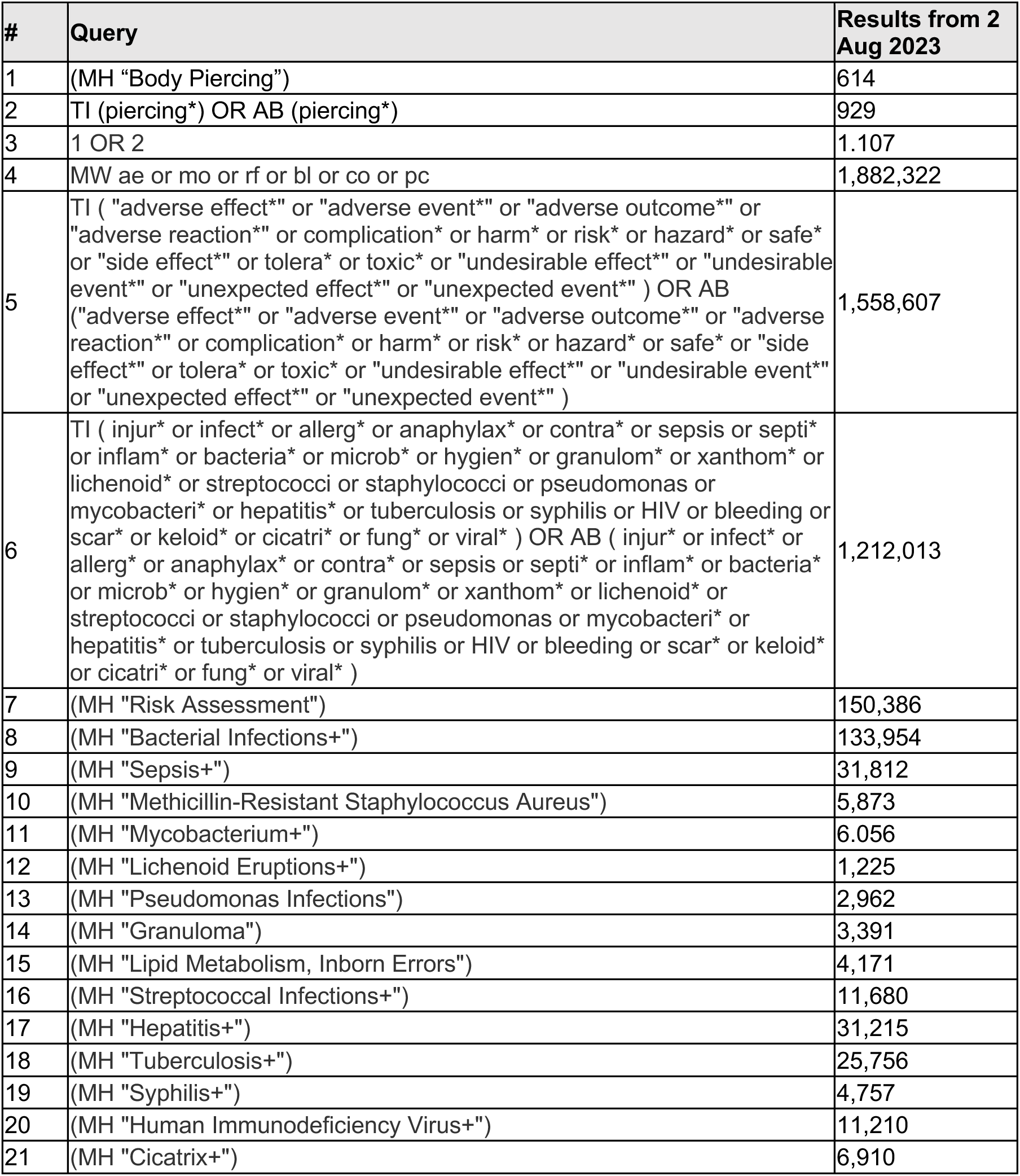
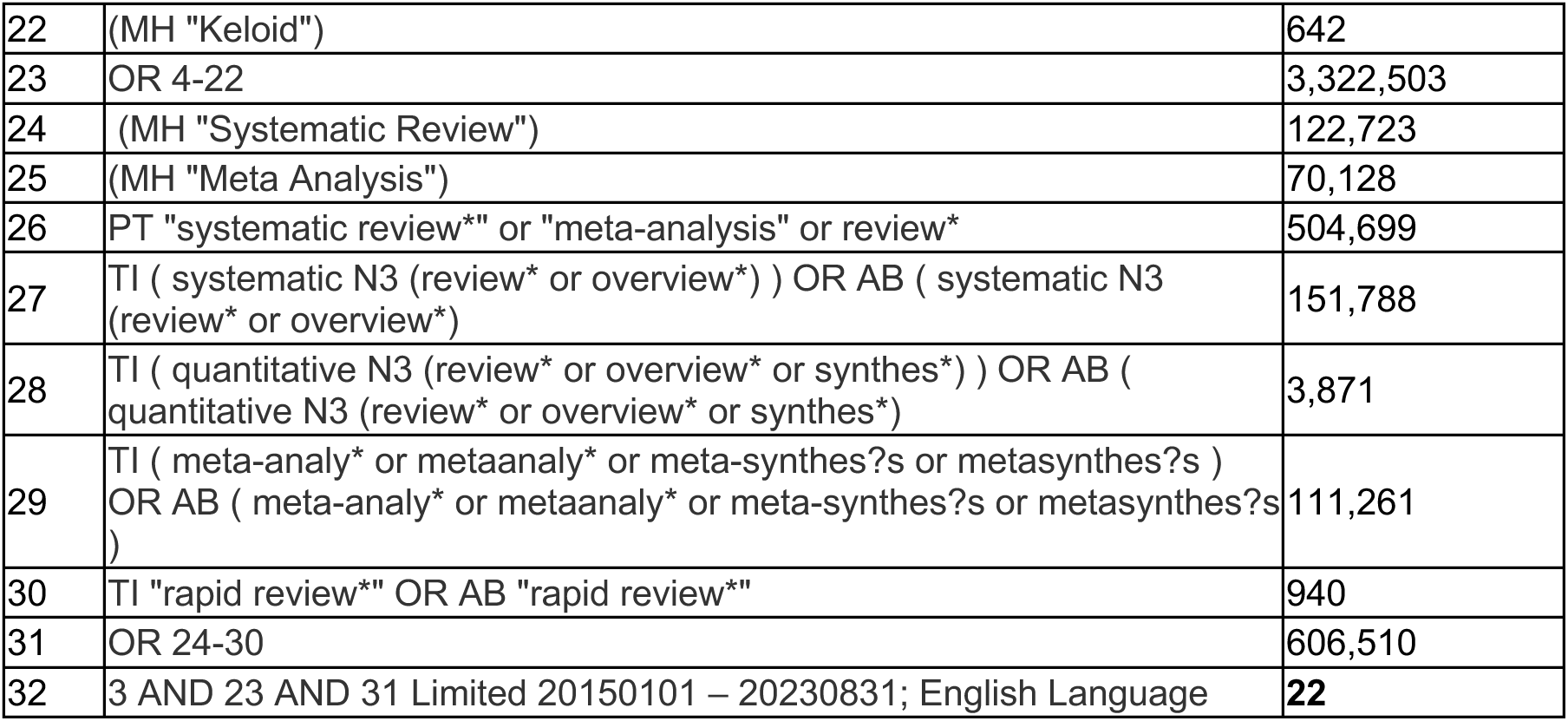

| # | Query | Results from 2 Aug 2023 |
| --- | --- | --- |
| 1 | (MH "Body Piercing") | 614 |
| 2 | TI (piercing*) OR AB (piercing*) | 929 |
| 3 | 1 OR 2 | 1,107 |
| 4 | MW ae or mo or rf or bl or co or pc | 1,882,322 |
| 5 | TI ( "adverse effect*" or "adverse event*" or "adverse outcome*" or "adverse reaction*" or complication* or harm* or risk* or hazard* or safe* or "side effect*" or tolera* or toxic* or "undesirable effect*" or "undesirable event*" or "unexpected effect*" or "unexpected event*" ) OR AB ("adverse effect*" or "adverse event*" or "adverse outcome*" or "adverse reaction*" or complication* or harm* or risk* or hazard* or safe* or "side effect*" or tolera* or toxic* or "undesirable effect*" or "undesirable event*" or "unexpected effect*" or "unexpected event*" ) | 1,558,607 |
| 6 | TI ( injur* or infect* or allerg* or anaphylax* or contra* or sepsis or septi* or inflam* or bacteria* or microb* or hygien* or granulom* or xanthom* or lichenoid* or streptococci or staphylococci or pseudomonas or mycobacteri* or hepatitis* or tuberculosis or syphilis or HIV or bleeding or scar* or keloid* or cicatri* or fung* or viral* ) OR AB ( injur* or infect* or allerg* or anaphylax* or contra* or sepsis or septi* or inflam* or bacteria* or microb* or hygien* or granulom* or xanthom* or lichenoid* or streptococci or staphylococci or pseudomonas or mycobacteri* or hepatitis* or tuberculosis or syphilis or HIV or bleeding or scar* or keloid* or cicatri* or fung* or viral* ) | 1,212,013 |
| 7 | (MH "Risk Assessment") | 150,386 |
| 8 | (MH "Bacterial Infections+") | 133,954 |
| 9 | (MH "Sepsis+") | 31,812 |
| 10 | (MH "Methicillin-Resistant Staphylococcus Aureus") | 5,873 |
| 11 | (MH "Mycobacterium+") | 6,056 |
| 12 | (MH "Lichenoid Eruptions+") | 1,225 |
| 13 | (MH "Pseudomonas Infections") | 2,962 |
| 14 | (MH "Granuloma") | 3,391 |
| 15 | (MH "Lipid Metabolism, Inborn Errors") | 4,171 |
| 16 | (MH "Streptococcal Infections+") | 11,680 |
| 17 | (MH "Hepatitis+") | 31,215 |
| 18 | (MH "Tuberculosis+") | 25,756 |
| 19 | (MH "Syphilis+") | 4,757 |
| 20 | (MH "Human Immunodeficiency Virus+") | 11,210 |
| 21 | (MH "Cicatrix+") | 6,910 |
| 22 | (MH "Keloid") | 642 |
| 23 | OR 4-22 | 3,322,503 |
| 24 | (MH "Systematic Review") | 122,723 |
| 25 | (MH "Meta Analysis") | 70,128 |
| 26 | PT "systematic review*" or "meta-analysis" or review* | 504,699 |
| 27 | TI ( systematic N3 (review* or overview* ) ) OR AB ( systematic N3 (review* or overview* ) ) | 151,788 |
| 28 | TI ( quantitative N3 (review* or overview* or syntheses* ) ) OR AB ( quantitative N3 (review* or overview* or syntheses* ) ) | 3,871 |
| 29 | TI ( meta-analy* or metaanaly* or meta-synthes?s or metasynthes?s ) OR AB ( meta-analy* or metaanaly* or meta-synthes?s or metasynthes?s ) | 111,261 |
| 30 | TI "rapid review*" OR AB "rapid review*" | 940 |
| 31 | OR 24-30 | 606,510 |
| 32 | 3 AND 23 AND 31 Limited 20150101 – 20230831; English Language | **22** |

#### COCHRANE LIBRARY 02.08.2023

| # | Query | Results from 2 Aug 2023 |
| --- | --- | --- |
| 1 | MeSH descriptor: [Body Piercing] explode all trees | 8 |
| 2 | (piercing*).ti,ab,kw | 144 |
| 3 | 1 OR 2 | (144 trials, **5 reviews**) |

#### Total References Added to Endnote

| Database | References |
| --- | --- |
| MEDLINE (Ovid) | 89 |
| EMBASE (Ovid) | 89 |
| Ovid EMCARE | 31 |
| AMED (Ovid) | 0 |
| CINAHL (EBSCO) | 22 |
| COCHRANE LIBRARY (Reviews) | 5 |
| Epistemonikos | 34 |
| **TOTAL** | **270** |
| Duplicates Identified in Endnote | 113 |
| **New TOTAL** | **157** |

### Q3: Acupuncture

#### MEDLINE (Ovid) 03.08.2023

Q3: Acupuncture MEDLINE (Ovid) 03.08.2023
| # | Query | Results from 3 Aug 2023 |
| --- | --- | --- |
| 1 | Acupuncture/ | 2,036 |
| 2 | exp Acupuncture Therapy/ | 29,161 |
| 3 | Dry Needling/ | 340 |
| 4 | (acupuncture or acupoint).tw. | 28,748 |
| 5 | (electroacupuncture or electro-acupuncture).tw. | 6,718 |
| 6 | moxibustion.tw. | 3,570 |
| 7 | (dry needling or intramuscular stimulation).tw. | 972 |
| 8 | or/1-7 | 39,017 |
| 9 | (ae or co or in or to or bl or mo).fs. | 6,446,639 |
| 10 | (adverse effect* or adverse event* or adverse outcome* or adverse reaction* or complication* or harm* or risk* or hazard* or safe* or side effect* or tolera* or toxic* or undesirable effect* or undesirable event* or unexpected effect* or unexpected event*).tw. | 5,999,798 |
| 11 | (injur* or infect* or allerg* or anaphylax* or contra* or sepsis or septi* or inflam* or bacteria* or microb* or hygien* or granulom* or xanthom* or lichenoid* or streptococci or staphylococci or pseudomonas or mycobacteri* or hepatitis* or tuberculosis or syphilis or HIV or bleeding or scar* or keloid* or cicatri* or fung* or viral*).tw. | 7,603,536 |
| 12 | exp Risk Assessment/ | 310,842 |
| 13 | bacterial infections/ | 74,912 |
| 14 | exp Sepsis/ | 141,692 |
| 15 | exp Blood-Borne Infections/ | 427,100 |
| 16 | exp Methicillin-Resistant Staphylococcus aureus/ | 19,627 |
| 17 | Mycobacterium/ or Lichenoid Eruptions/ or Pseudomonas/ or Granuloma/ or Xanthomatosis/ | 65,705 |
| 18 | streptococcal infections/ | 35,042 |
| 19 | staphylococcal infections/ or staphylococcal skin infections/ | 66,946 |
| 20 | hepatitis/ or exp hepatitis, viral, human/ | 170,622 |
| 21 | tuberculosis/ or tuberculosis, cutaneous/ | 115,494 |
| 22 | Syphilis/ or Syphilis, Cutaneous/ | 24,239 |
| 23 | exp HIV/ | 107,650 |
| 24 | cicatrix/ or keloid/ | 29,644 |
| 25 | or/9-24 | 15,083,129 |
| 26 | 8 and 25 | 17,461 |
| 27 | umbrella review.ti,ab,kf. | 1,472 |
| 28 | evidence map*.ti,ab,kf. | 545 |
| 29 | ((review or overview or synthesis) adj3 reviews).ti,ab,kf. | 5,254 |
| 30 | "review* of systematic review*".tw,pt,kf. | 2,183 |
| 31 | "overview* of systematic review*".tw,pt,kf. | 1,085 |
| 32 | (meta-analys#s adj3 overview*).ti. | 143 |
| 33 | or/27-32 | 7,974 |
| 34 | 26 and 33 | 143 |
| 35 | limit 34 to (english language and yr="2015 -Current") | **114** |

#### EMBASE (Ovid) 03.08.2023

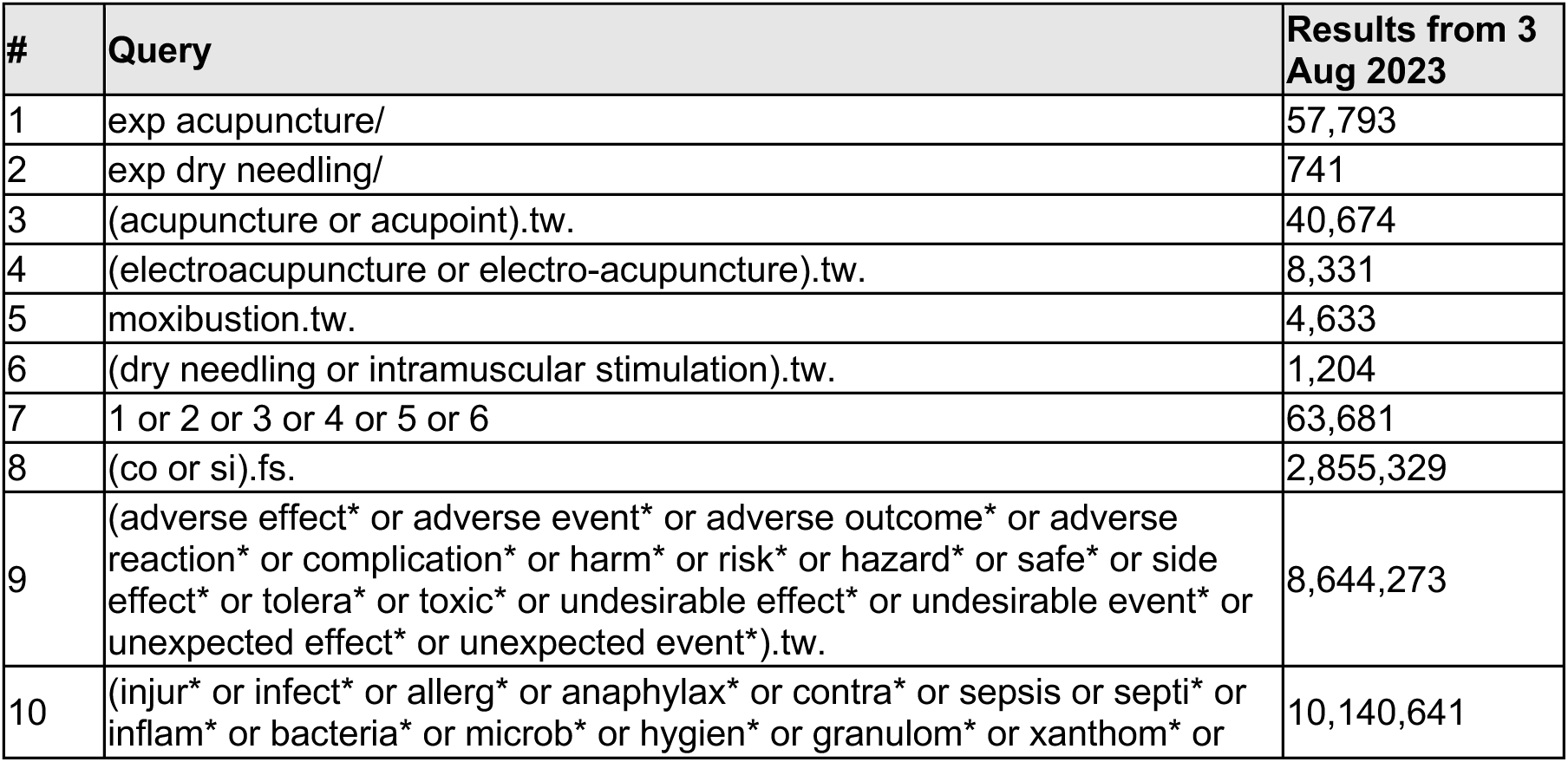
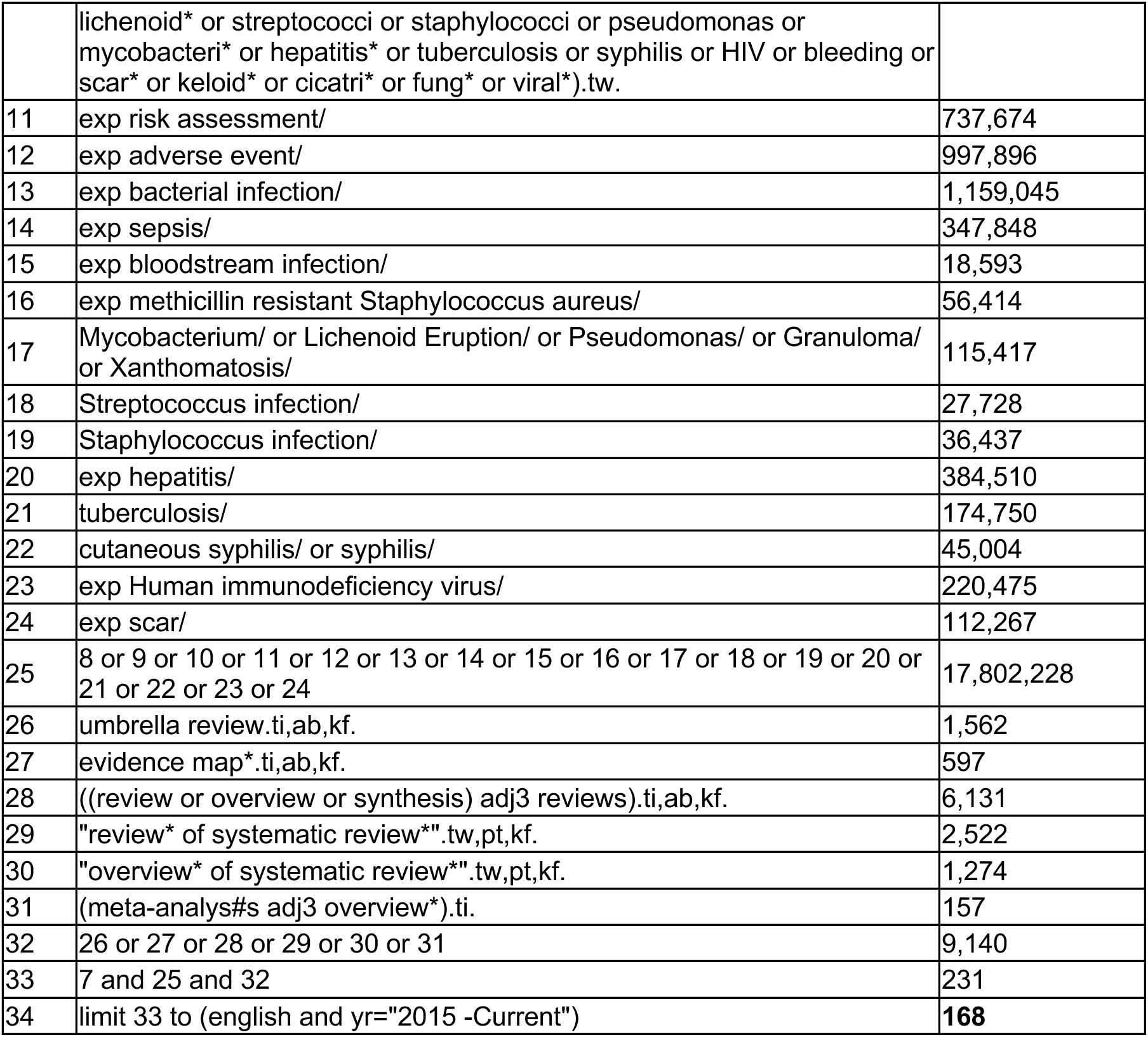

#### Ovid EMCARE 03.08.2023

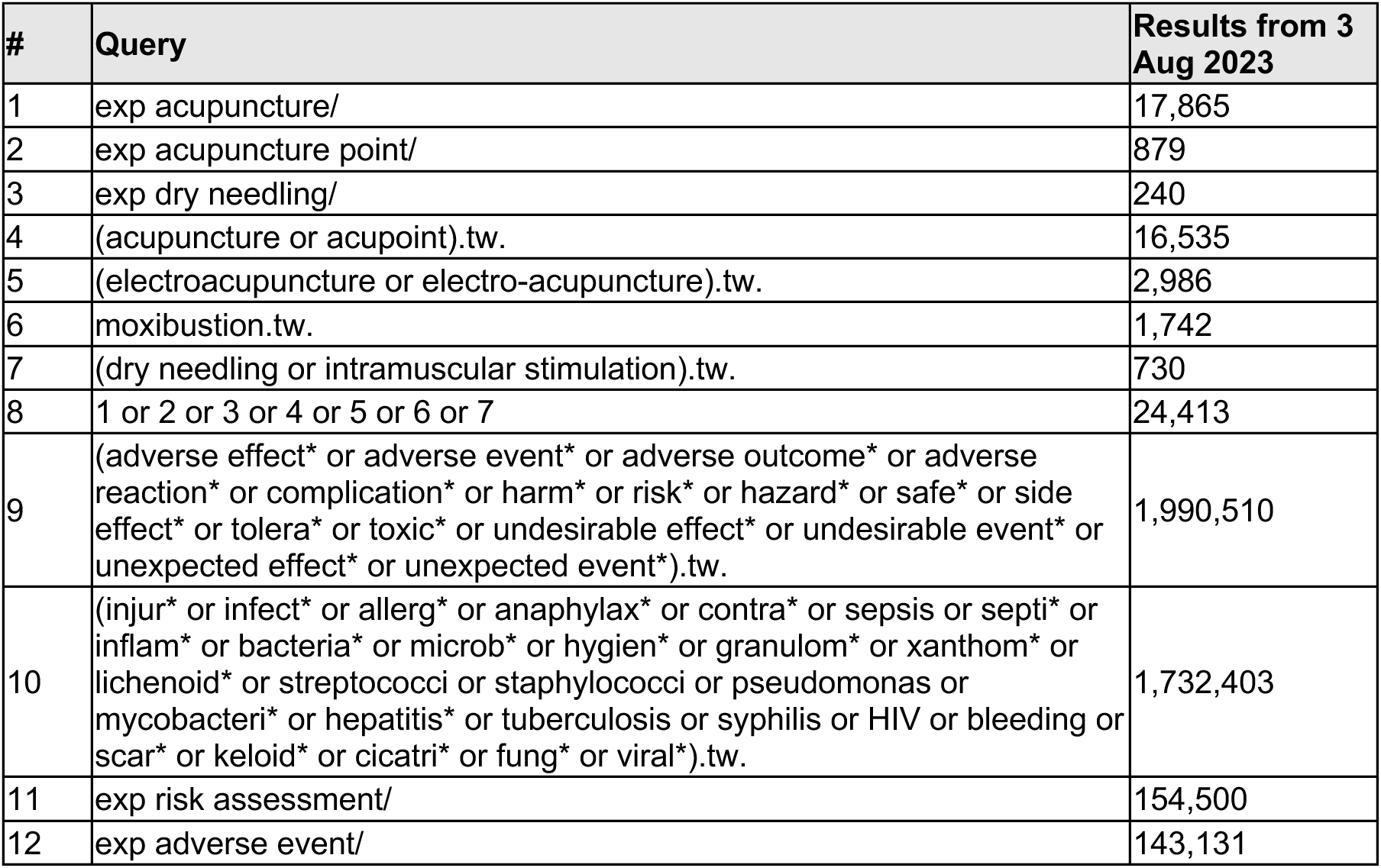
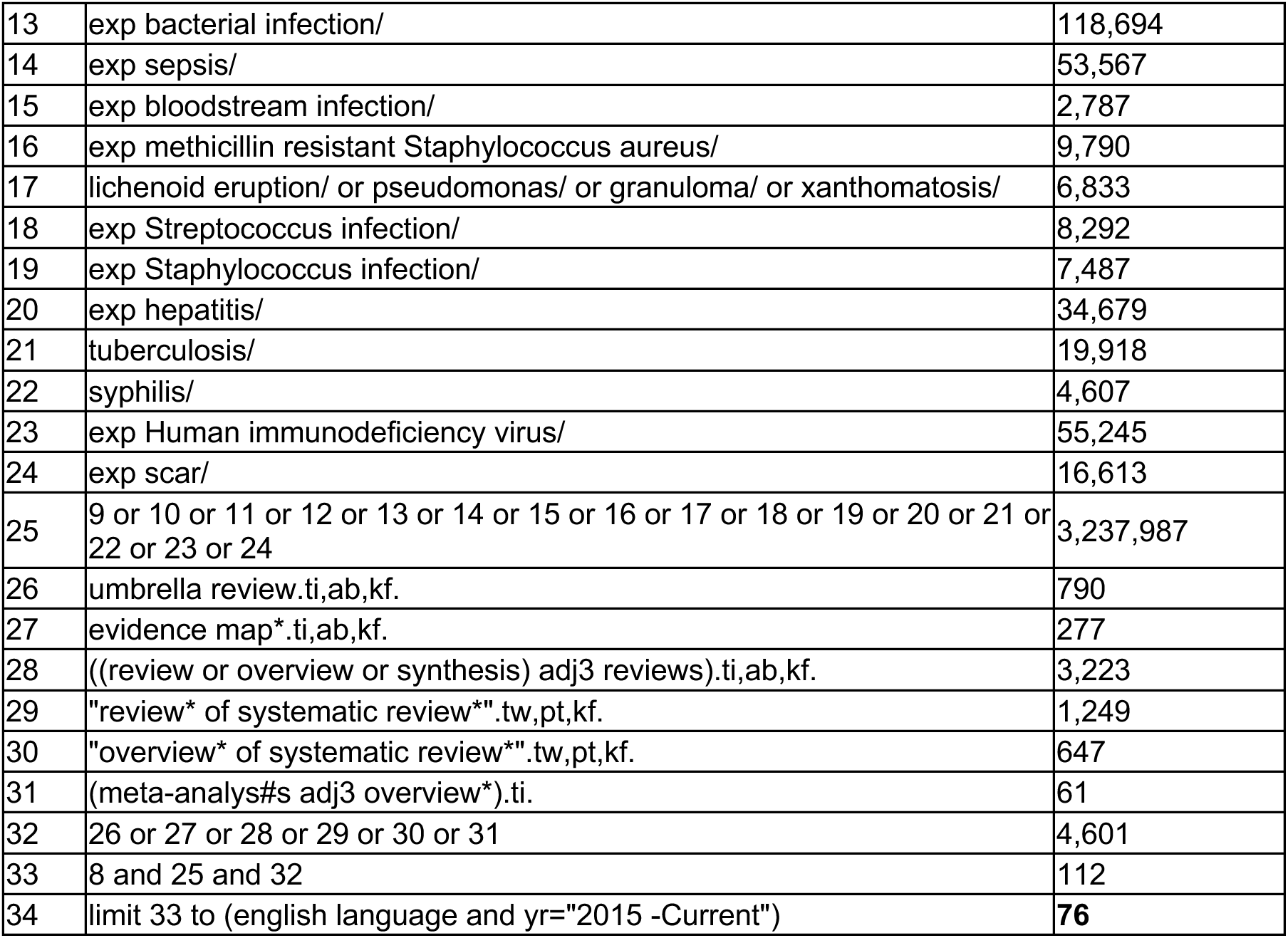

| # | Query | Results from 3 Aug 2023 |
| --- | --- | --- |
| 1 | exp acupuncture/ | 17,865 |
| 2 | exp acupuncture point/ | 879 |
| 3 | exp dry needling/ | 240 |
| 4 | (acupuncture or acupoint).tw. | 16,535 |
| 5 | (electroacupuncture or electro-acupuncture).tw. | 2,986 |
| 6 | moxibustion.tw. | 1,742 |
| 7 | (dry needling or intramuscular stimulation).tw. | 730 |
| 8 | 1 or 2 or 3 or 4 or 5 or 6 or 7 | 24,413 |
| 9 | (adverse effect* or adverse event* or adverse outcome* or adverse reaction* or complication* or harm* or risk* or hazard* or safe* or side effect* or tolera* or toxic* or undesirable effect* or undesirable event* or unexpected effect* or unexpected event*).tw. | 1,990,510 |
| 10 | (injur* or infect* or allerg* or anaphylax* or contra* or sepsis or septi* or inflam* or bacteria* or microb* or hygien* or granulom* or xanthom* or lichenoid* or streptococci or staphylococci or pseudomonas or mycobacteri* or hepatitis* or tuberculosis or syphilis or HIV or bleeding or scar* or keloid* or cicatri* or fung* or viral*).tw. | 1,732,403 |
| 11 | exp risk assessment/ | 154,500 |
| 12 | exp adverse event/ | 143,131 |
| 13 | exp bacterial infection/ | 118,694 |
| 14 | exp sepsis/ | 53,567 |
| 15 | exp bloodstream infection/ | 2,787 |
| 16 | exp methicillin resistant Staphylococcus aureus/ | 9,790 |
| 17 | lichenoid eruption/ or pseudomonas/ or granuloma/ or xanthomatosis/ | 6,833 |
| 18 | exp Streptococcus infection/ | 8,292 |
| 19 | exp Staphylococcus infection/ | 7,487 |
| 20 | exp hepatitis/ | 34,679 |
| 21 | tuberculosis/ | 19,918 |
| 22 | syphilis/ | 4,607 |
| 23 | exp Human immunodeficiency virus/ | 55,245 |
| 24 | exp scar/ | 16,613 |
| 25 | 9 or 10 or 11 or 12 or 13 or 14 or 15 or 16 or 17 or 18 or 19 or 20 or 21 or 22 or 23 or 24 | 3,237,987 |
| 26 | umbrella review.ti,ab,kf. | 790 |
| 27 | evidence map*.ti,ab,kf. | 277 |
| 28 | ((review or overview or synthesis) adj3 reviews).ti,ab,kf. | 3,223 |
| 29 | "review* of systematic review*".tw,pt,kf. | 1,249 |
| 30 | "overview* of systematic review*".tw,pt,kf. | 647 |
| 31 | (meta-analys#s adj3 overview*).ti. | 61 |
| 32 | 26 or 27 or 28 or 29 or 30 or 31 | 4,601 |
| 33 | 8 and 25 and 32 | 112 |
| 34 | limit 33 to (english language and yr="2015 -Current") | 76 |

#### AMED (Ovid) 03.08.23

| # | Query | Results from 3 Aug 2023 |
| --- | --- | --- |
| 1 | exp Acupuncture/ | 3,538 |
| 2 | exp Acupuncture therapy/ | 9,383 |
| 3 | (acupuncture or acupoint).tw. | 11,301 |
| 4 | (electroacupuncture or electro-acupuncture).tw. | 1,430 |
| 5 | moxibustion.tw. | 878 |
| 6 | (dry needling or intramuscular stimulation).tw. | 229 |
| 7 | 1 or 2 or 3 or 4 or 5 or 6 | 12,819 |
| 8 | (adverse effect* or adverse event* or adverse outcome* or adverse reaction* or complication* or harm* or risk* or hazard* or safe* or side effect* or tolera* or toxic* or undesirable effect* or undesirable event* or unexpected effect* or unexpected event*).tw. | 55,239 |
| 9 | (injur* or infect* or allerg* or anaphylax* or contra* or sepsis or septi* or inflam* or bacteria* or microb* or hygien* or granulom* or xanthom* or lichenoid* or streptococci or staphylococci or pseudomonas or mycobacteri* or hepatitis* or tuberculosis or syphilis or HIV or bleeding or scar* or keloid* or cicatri* or fung* or viral*).tw. | 73,313 |
| 10 | exp Adverse effects/ | 6,832 |
| 11 | exp Bacterial infections/ | 1,552 |
| 12 | exp Septicemia/ | 70 |
| 13 | exp Streptococcal infections/ | 40 |
| 14 | exp Hepatitis/ | 326 |
| 15 | exp Tuberculosis/ | 231 |
| 16 | exp Syphilis/ | 21 |
| 17 | exp Hiv/ | 61 |
| 18 | exp Cicatrix/ | 104 |
| 19 | 8 or 9 or 10 or 11 or 12 or 13 or 14 or 15 or 16 or 17 or 18 | 111,727 |
| 20 | 7 and 19 | 3,377 |
| 21 | umbrella review.tw. | 14 |
| 22 | evidence map*.tw. | 14 |
| 23 | ((review or overview or synthesis) adj3 reviews).tw. | 219 |
| 24 | "review* of systematic review*".tw.pt. | 43 |
| 25 | "overview* of systematic review*".tw.pt. | 70 |
| 26 | (meta-analys#s adj3 overview*).tw. | 25 |
| 27 | 21 or 22 or 23 or 24 or 25 or 26 | 260 |
| 28 | 20 and 27 | 30 |
| 29 | limit 28 to (english and yr="2015 -Current") | 21 |

#### CINAHL (EBSCO) 03.08.2023

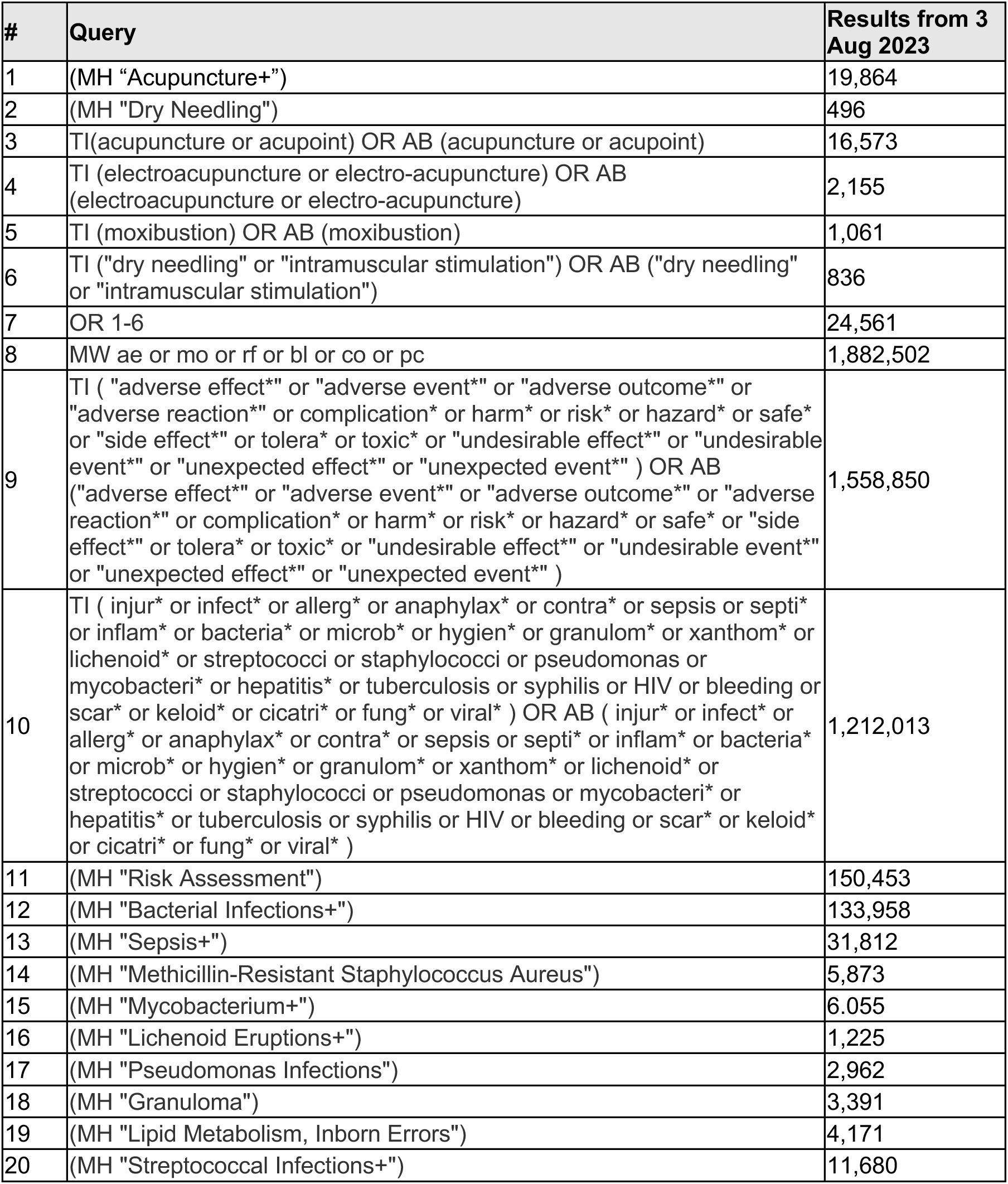
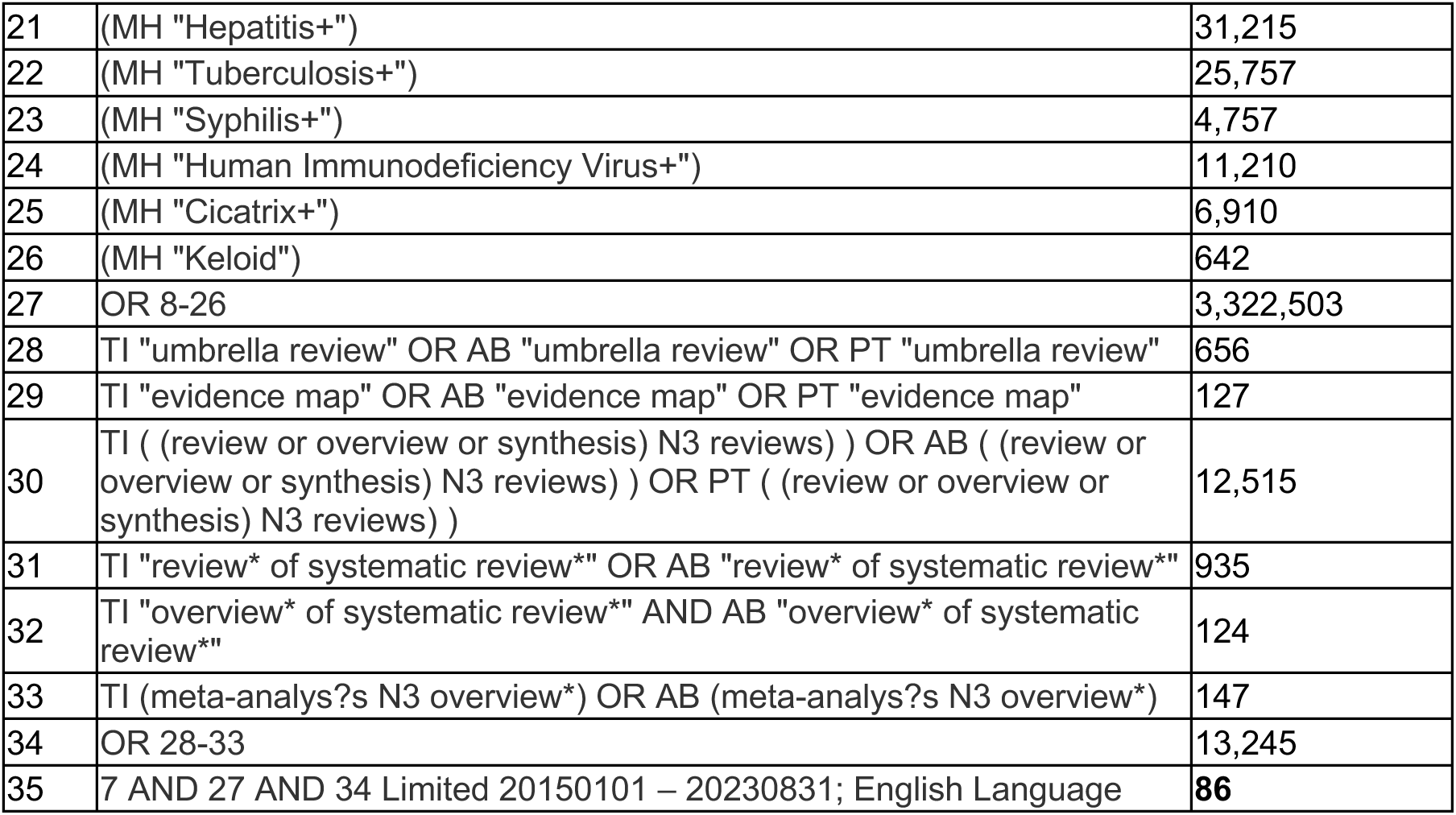

#### COCHRANE LIBRARY 03.08.2023

| # | Query | Results from 3 Aug 2023 |
| --- | --- | --- |
| 1 | MeSH descriptor: [Acupuncture] explode all trees | 713 |
| 2 | MeSH descriptor: [Dry Needling] explode all trees | 164 |
| 3 | (acupuncture or acupoint):ti,ab,kw (Word variations have been searched) | 20,929 |
| 4 | (electroacupuncture or electro-acupuncture):ti,ab,kw (Word variations have been searched) | 3,452 |
| 5 | (moxibustion):ti,ab,kw (Word variations have been searched) | 2,413 |
| 6 | ("dry needling" or "intramuscular stimulation"):ti,ab,kw (Word variations have been searched) | 1,115 |
| 7 | OR 1-6 | 23,624 |
| 8 | (adverse NEAR (effect* OR event* OR outcome* OR reaction*)):ti,ab,kw (Word variations have been searched) | 353,753 |
| 9 | (undesirable NEAR (event* OR effect*)):ti,ab,kw (Word variations have been searched) | 1,635 |
| 10 | (unexpected NEAR (event* OR effect*)):ti,ab,kw (Word variations have been searched) | 1,836 |
| 11 | (complication* or harm* or risk* or hazard* or safe* or tolera* or toxic*):ti,ab,kw (Word variations have been searched) | 807,238 |
| 12 | ((side NEAR effect*)):ti,ab,kw (Word variations have been searched) | 161,989 |
| 13 | (injur* or infect* or allerg* or anaphylax* or contra* or sepsis or septi* or inflam* or bacteria* or microb* or hygien* or granulom* or xanthom* or lichenoid* or streptococci or staphylococci or pseudomonas or mycobacteri* or hepatitis* or tuberculosis or syphilis or HIV or bleeding or scar* or keloid* or cicatri* or fung* or viral*):ti,ab,kw (Word variations have been searched) | 536,215 |
| 14 | MeSH descriptor: [Risk Assessment] explode all trees | 13,627 |
| 15 | MeSH descriptor: [Bacterial Infections] explode all trees | 21,585 |
| 16 | MeSH descriptor: [Sepsis] explode all trees | 6,783 |
| 17 | MeSH descriptor: [Blood-Borne Infections] explode all trees | 23,057 |
| 18 | MeSH descriptor: [Mycobacterium] explode all trees | 816 |
| 19 | MeSH descriptor: [Lichenoid Eruptions] explode all trees | 388 |
| 20 | MeSH descriptor: [Pseudomonas] explode all trees | 712 |
| 21 | MeSH descriptor: [Granuloma] explode all trees | 172 |
| 22 | MeSH descriptor: [Xanthomatosis] explode all trees | 19 |
| 23 | MeSH descriptor: [Streptococcal Infections] explode all trees | 2,031 |
| 24 | MeSH descriptor: [Staphylococcal Infections] explode all trees | 1,434 |
| 25 | MeSH descriptor: [Staphylococcal Skin Infections] explode all trees | 281 |
| 26 | MeSH descriptor: [Hepatitis] explode all trees | 8,403 |
| 27 | MeSH descriptor: [Tuberculosis] explode all trees | 3,311 |
| 28 | MeSH descriptor: [Syphilis] explode all trees | 228 |
| 29 | MeSH descriptor: [HIV] explode all trees | 3,755 |
| 30 | MeSH descriptor: [Cicatrix] explode all trees | 2,118 |
| 31 | MeSH descriptor: [Keloid] explode all trees | 224 |
| 32 | OR 8-31 | 1,159,972 |
| 33 | 7 AND 32 | 9,133<br>(142 Cochrane reviews, 10 Protocols) |
| 34 | Limited to 2015-2023 & Overviews | 2 |

\*Did look through the 142 reviews – no other relevant overviews missed by the overview filter
\*10 protocols not relevant

#### Epistemonikos 03.08.2023

| # | Query | Results from 3 Aug 2023 |
| --- | --- | --- |
| 1 | (title:(acupuncture OR acupoint OR electroacupuncture OR electro-acupuncture OR moxibustion OR "dry needling" OR "intramuscular stimulation") OR abstract:(acupuncture OR acupoint OR electroacupuncture OR electro-acupuncture OR moxibustion OR "dry needling" OR "intramuscular stimulation")) | 15,299 |
| 2 | (title:("adverse effect*" OR "adverse event*" OR "adverse outcome*" OR "adverse reaction*" OR complication* OR harm* OR risk* OR hazard* OR safe* OR "side effect*" OR tolera* OR toxic* OR "undesirable effect*" OR "undesirable event*" OR "unexpected effect*" OR "unexpected event*" OR injur* OR infect* OR allerg* OR anaphylax* OR contra* OR sepsis OR septi* OR inflam* OR bacteria* OR microb* OR hygien* OR granulom* OR xanthom* OR lichenoid* OR streptococci OR staphylococci OR pseudomonas OR mycobacteri* OR hepatitis* OR tuberculosis OR syphilis OR HIV OR bleeding OR scar* OR keloid* OR cicatri* OR fung* OR viral*) OR abstract:("adverse effect*" OR "adverse event*" OR "adverse outcome*" OR "adverse reaction*" OR complication* OR harm* OR risk* OR hazard* OR safe* OR "side effect*" OR tolera* OR toxic* OR "undesirable effect*" OR "undesirable event*" OR "unexpected effect*" OR "unexpected event*" OR injur* OR infect* OR allerg* OR anaphylax* OR contra* OR sepsis OR septi* OR inflam* OR bacteria* OR microb* OR hygien* OR granulom* OR xanthom* OR lichenoid* OR streptococci OR staphylococci OR pseudomonas OR mycobacteri* OR hepatitis* OR tuberculosis OR syphilis OR HIV OR bleeding OR scar* OR keloid* OR cicatri* OR fung* OR viral*)) | 1,812,067 |
| 3 | (title:("umbrella review" OR "evidence map" OR "review of systematic review" OR overview*) OR abstract:("umbrella review" OR "evidence map" OR "review of systematic review" OR overview*)) | 19,628 |
|  | 1 AND 2 = 5,311 (1,921 SR's 2015-2023) and (64 Broad Synthesis / Overview of SR's 2015-2023) |  |
| 4 | 1 AND 2 AND 3 | 111 |
| 5 | 4 Limited to 2015-2023 Systematic Reviews | 44 |
| 6 | 4 Limited to 2015-2023 Broad synthesis / Overview of systematic reviews | 42 |
|  | Exported to ENDNOTE | 86 |

\* Publication Type: Systematic review did contain umbrella reviews and evidence maps so included all. Of the 111 studies – 88 were published between 2015 and 2023. 2 of these were primary studies so the remainder (86) exported to Endnote.

#### Total References Added to Endnote

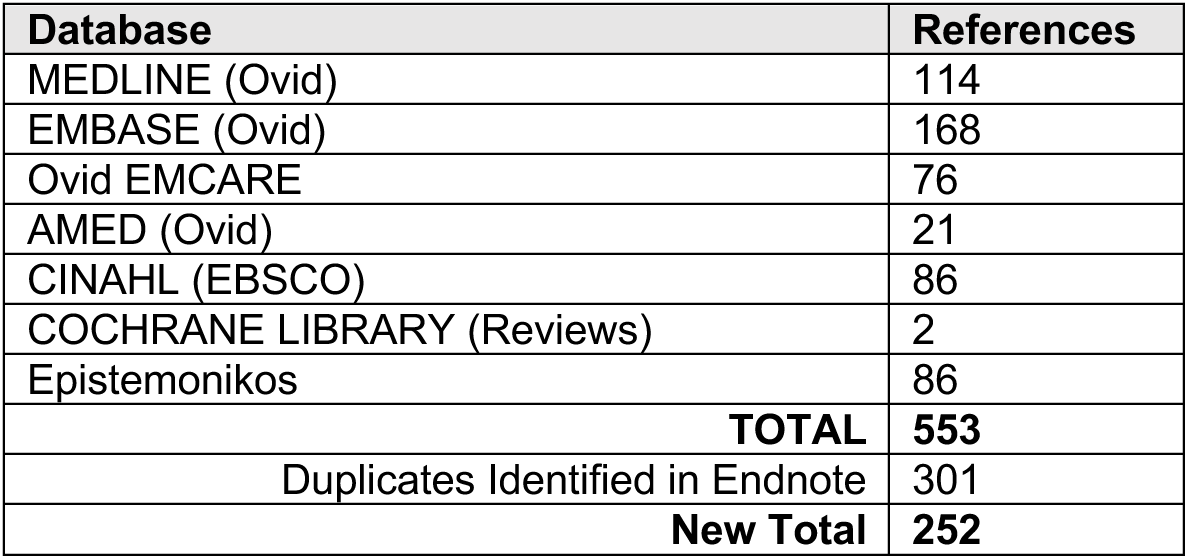

| Database | References |
| --- | --- |
| MEDLINE (Ovid) | 114 |
| EMBASE (Ovid) | 168 |
| Ovid EMCARE | 76 |
| AMED (Ovid) | 21 |
| CINAHL (EBSCO) | 86 |
| COCHRANE LIBRARY (Reviews) | 2 |
| Epistemonikos | 86 |
| **TOTAL** | **553** |
| Duplicates Identified in Endnote | 301 |
| **New Total** | **252** |

### Q4: Electrolysis

#### MEDLINE (Ovid) 07.08.2023

Q4: Electrolysis MEDLINE (Ovid) 07.08.2023
| # | Query | Results from 7 Aug 2023 |
| --- | --- | --- |
| 1 | exp Hair Removal/ | 2,057 |
| 2 | exp Electrolysis/ | 5,374 |
| 3 | (electrolysis or electroepilat* or electro-epilat* or electrolytic-epilat* or electrolytic-depilat* or needle-epilat* or thermolysis or electrologist* or electrology or epilatt* or depilat*).tw. | 10,915 |
| 4 | (hair* or cosmet*).tw. | 194,887 |
| 5 | 2 or 3 | 14,442 |
| 6 | 4 and 5 | 878 |
| 7 | 1 or 6 | 2,530 |
| 8 | (ae or co or in or to or bl or mo).fs. | 6,447,732 |
| 9 | (adverse effect* or adverse event* or adverse outcome* or adverse reaction* or complication* or harm* or risk* or hazard* or safe* or side effect* or tolera* or toxic* or undesirable effect* or undesirable event* or unexpected effect* or unexpected event*).tw. | 6,002,405 |
| 10 | (injur* or infect* or allerg* or anaphylax* or contra* or sepsis or septi* or inflam* or bacteria* or microb* or hygien* or granulom* or xanthom* or lichenoid* or streptococci or staphylococci or pseudomonas or mycobacteri* or hepatitis* or tuberculosis or syphilis or HIV or bleeding or scar* or keloid* or cicatri* or fung* or viral*).tw. | 7,605,996 |
| 11 | exp Risk Assessment/ | 310,875 |
| 12 | bacterial infections/ | 74,921 |
| 13 | exp Sepsis/ | 141,721 |
| 14 | exp Blood-Borne Infections/ | 427,191 |
| 15 | exp Methicillin-Resistant Staphylococcus aureus/ | 19,636 |
| 16 | Mycobacterium/ or Lichenoid Eruptions/ or Pseudomonas/ or Granuloma/ or Xanthomatosis/ | 65,710 |
| 17 | streptococcal infections/ | 35,045 |
| 18 | staphylococcal infections/ or staphylococcal skin infections/ | 66,955 |
| 19 | hepatitis/ or exp hepatitis, viral, human/ | 170,637 |
| 20 | tuberculosis/ or tuberculosis, cutaneous/ | 115,508 |
| 21 | Syphilis/ or Syphilis, Cutaneous/ | 24,242 |
| 22 | exp HIV/ | 107,674 |
| 23 | cicatrix/ or keloid/ | 29,646 |
| 24 | or/8-23 | 15,087,644 |
| 25 | 7 and 24 | 1,501 |
| 26 | limit 25 to (english language and yr="2015 -Current") | **412** |

EMBASE (Ovid) 07.08.2023
| # | Query | Results from 7 Aug 2023 |
| --- | --- | --- |
| 1 | exp Hair Removal/ | 1,367 |
| 2 | exp Electrolysis/ | 10,870 |
| 3 | (electrolysis or electroepilat* or electro-epilat* or electrolytic-epilat* or electrolytic-depilat* or needle-epilat* or thermolysis or electrologist* or electrology or epilatt* or depilat*).tw. | 12,711 |
| 4 | (hair* or cosmet*).tw. | 261,206 |
| 5 | 2 or 3 | 18,060 |
| 6 | 4 and 5 | 1,534 |
| 7 | 1 or 6 | 2,649 |
| 8 | (co or si).fs. | 2,857,996 |
| 9 | (adverse effect* or adverse event* or adverse outcome* or adverse reaction* or complication* or harm* or risk* or hazard* or safe* or side effect* or tolera* or toxic* or undesirable effect* or undesirable event* or unexpected effect* or unexpected event*).tw. | 8,649,158 |
| 10 | (injur* or infect* or allerg* or anaphylax* or contra* or sepsis or septi* or inflam* or bacteria* or microb* or hygien* or granulom* or xanthom* or lichenoid* or streptococci or staphylococci or pseudomonas or mycobacteri* or hepatitis* or tuberculosis or syphilis or HIV or bleeding or scar* or keloid* or cicatri* or fung* or viral*).tw. | 10,145,598 |
| 11 | exp risk assessment/ | 737,782 |
| 12 | exp adverse event/ | 998,469 |
| 13 | exp bacterial infection/ | 1,159,461 |
| 14 | exp sepsis/ | 348,072 |
| 15 | exp bloodstream infection/ | 18,600 |
| 16 | exp methicillin resistant Staphylococcus aureus/ | 56,465 |
| 17 | Mycobacterium/ or Lichenoid Eruption/ or Pseudomonas/ or Granuloma/ or Xanthomatosis/ | 115,462 |
| 18 | Streptococcus infection/ | 27,734 |
| 19 | Staphylococcus infection/ | 36,454 |
| 20 | exp hepatitis/ | 384,785 |
| 21 | tuberculosis/ | 174,787 |
| 22 | cutaneous syphilis/ or syphilis/ | 45,020 |
| 23 | exp Human immunodeficiency virus/ | 220,487 |
| 24 | exp scar/ | 112,326 |
| 25 | 8 or 9 or 10 or 11 or 12 or 13 or 14 or 15 or 16 or 17 or 18 or 19 or 20 or 21 or 22 or 23 or 24 | 17,811,493 |
| 26 | 7 and 25 | 1,649 |
| 27 | conference abstract.pt. | 4,847,411 |
| 28 | 26 not 27 | 1,303 |
| 29 | limit 28 to (english language and yr="2015 -Current") | 708 |

#### EMBASE (Ovid) 07.08.2023

Ovid EMCARE 07.08.2023
| # | Query | Results from 7 Aug 2023 |
| --- | --- | --- |
| 1 | exp Hair Removal/ | 136 |
| 2 | exp Electrolysis/ | 627 |
| 3 | (electrolysis or electroepilat* or electro-epilat* or electrolytic-epilat* or electrolytic-depilat* or needle-epilat* or thermolysis or electrologist* or electrology or epilatt* or depilat*).tw. | 767 |
| 4 | (hair* or cosmet*).tw. | 34,198 |
| 5 | 2 or 3 | 1,116 |
| 6 | 4 and 5 | 197 |
| 7 | 1 or 6 | 313 |
| 8 | (adverse effect* or adverse event* or adverse outcome* or adverse reaction* or complication* or harm* or risk* or hazard* or safe* or side effect* or tolera* or toxic* or undesirable effect* or undesirable event* or unexpected effect* or unexpected event*).tw. | 1,997,778 |
| 9 | (injur* or infect* or allerg* or anaphylax* or contra* or sepsis or septi* or inflam* or bacteria* or microb* or hygien* or granulom* or xanthom* or lichenoid* or streptococci or staphylococci or pseudomonas or mycobacteri* or hepatitis* or tuberculosis or syphilis or HIV or bleeding or scar* or keloid* or cicatri* or fung* or viral*).tw. | 1,737,739 |
| 10 | exp risk assessment/ | 154,771 |
| 11 | exp adverse event/ | 143,225 |
| 12 | exp bacterial infection/ | 118,785 |
| 13 | exp sepsis/ | 53,590 |
| 14 | exp bloodstream infection/ | 2,789 |
| 15 | exp methicillin resistant Staphylococcus aureus/ | 9,792 |
| 16 | lichenoid eruption/ or pseudomonas/ or granuloma/ or xanthomatosis/ | 6,835 |
| 17 | exp Streptococcus infection/ | 8,294 |
| 18 | exp Staphylococcus infection/ | 7,489 |
| 19 | exp hepatitis/ | 34,707 |
| 20 | tuberculosis/ | 19,943 |
| 21 | syphilis/ | 4,614 |
| 22 | exp Human immunodeficiency virus/ | 55,283 |
| 23 | exp scar/ | 16,619 |
| 24 | 10 or 11 or 12 or 13 or 14 or 15 or 16 or 17 or 18 or 19 or 20 or 21 or 22 or 23 | 528,450 |
| 25 | 7 and 24 | 42 |
| 26 | limit 25 to (english language and yr="2015 -Current") | 20 |

#### Ovid EMCARE 07.08.2023

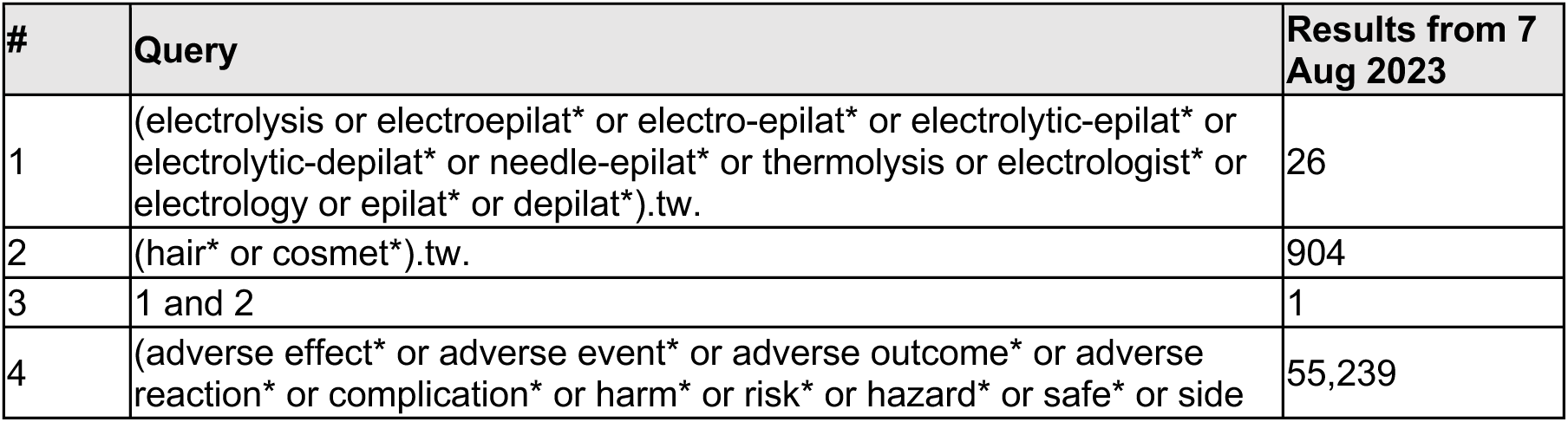
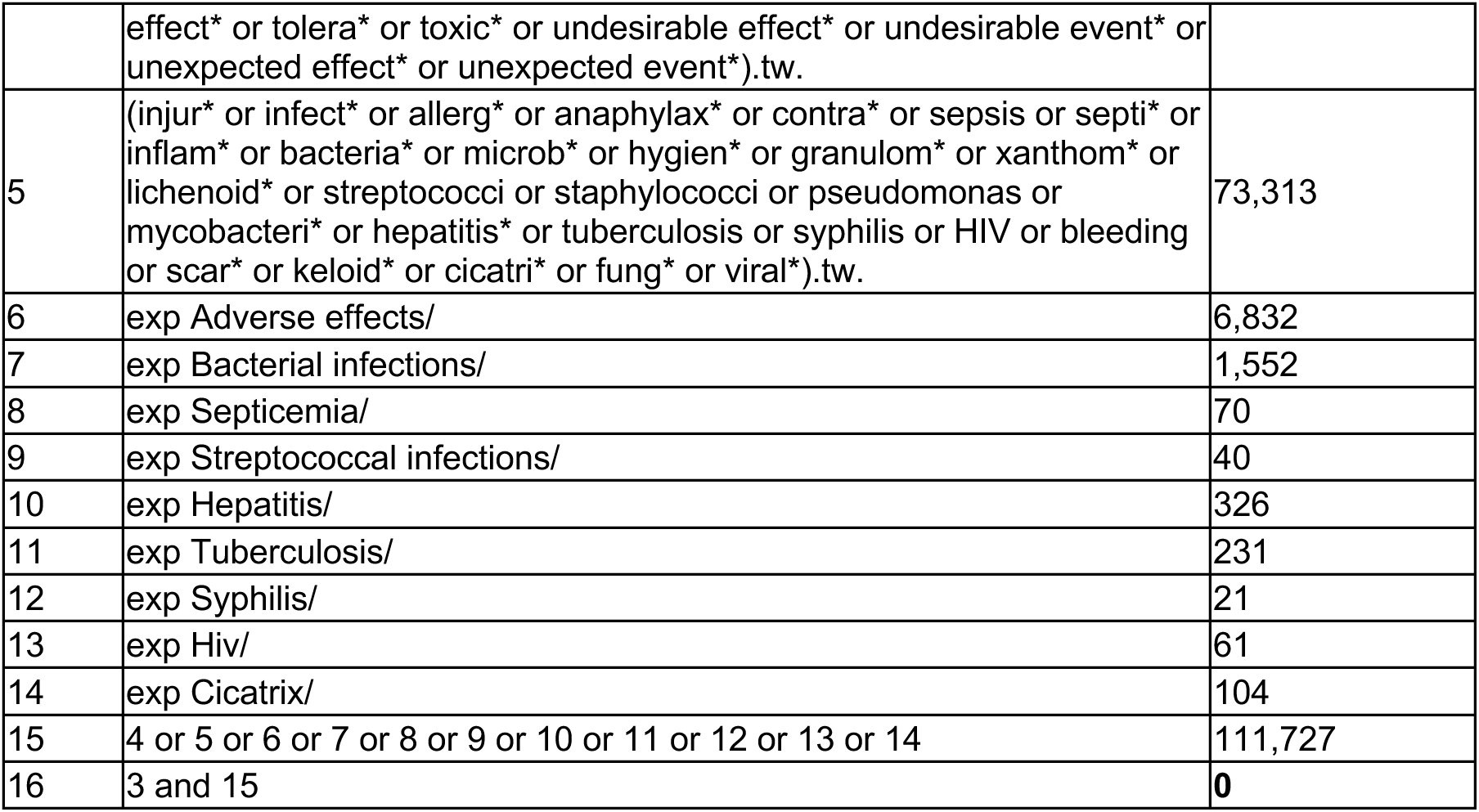

#### AMED (Ovid) 07.08.2023

CINAHL (EBSCO) 07.08.2023
| # | Query | Results from 3 Aug 2023 |
| --- | --- | --- |
| 1 | (MH "Hair Removal") | 739 |
| 2 | (MH "Electrolysis") | 136 |
| 3 | TI(electrolysis or electroepilat* or electro-epilat* or electrolytic-epilat* or electrolytic-depilat* or needle-epilat* or thermolysis or electrologist* or electrology or epil* or depilat*) OR AB (electrolysis or electroepilat* or electro-epilat* or electrolytic-epilat* or electrolytic-depilat* or needle-epilat* or thermolysis or electrologist* or electrology or epil* or depilat*) | 274 |
| 4 | TI (hair* or cosmet*) OR AB (hair* or cosmet*) | 22,687 |
| 5 | 2 or 3 | 364 |
| 6 | 4 and 5 | 120 |
| 7 | 1 or 6 | 798 |
| 8 | MW ae or mo or rf or bl or co or pc | 1,823,131 |
| 9 | TI ( "adverse effect*" or "adverse event*" or "adverse outcome*" or "adverse reaction*" or complication* or harm* or risk* or hazard* or safe* or "side effect*" or tolera* or toxic* or "undesirable effect*" or "undesirable event*" or "unexpected effect*" or "unexpected event*" ) OR AB ("adverse effect*" or "adverse event*" or "adverse outcome*" or "adverse reaction*" or complication* or harm* or risk* or hazard* or safe* or "side effect*" or tolera* or toxic* or "undesirable effect*" or "undesirable event*" or "unexpected effect*" or "unexpected event*" ) | 1,559,616 |
| 10 | TI ( injur* or infect* or allerg* or anaphylax* or contra* or sepsis or septi* or inflam* or bacteria* or microb* or hygien* or granulom* or xanthom* or lichenoid* or streptococci or staphylococci or pseudomonas or mycobacteri* or hepatitis* or tuberculosis or syphilis or HIV or bleeding or scar* or keloid* or cicatri* or fung* or viral* ) OR AB ( injur* or infect* or allerg* or anaphylax* or contra* or sepsis or septi* or inflam* or bacteria* or microb* or hygien* or granulom* or xanthom* or lichenoid* or streptococci or staphylococci or pseudomonas or mycobacteri* or hepatitis* or tuberculosis or syphilis or HIV or bleeding or scar* or keloid* or cicatri* or fung* or viral* ) | 1,212,586 |
| 11 | (MH "Risk Assessment") | 150,630 |
| 12 | (MH "Bacterial Infections+") | 133,984 |
| 13 | (MH "Sepsis+") | 31,818 |
| 14 | (MH "Methicillin-Resistant Staphylococcus Aureus") | 5,875 |
| 15 | (MH "Mycobacterium+") | 6,056 |
| 16 | (MH "Lichenoid Eruptions+") | 1,225 |
| 17 | (MH "Pseudomonas Infections") | 2,962 |
| 18 | (MH "Granuloma") | 3,391 |
| 19 | (MH "Lipid Metabolism, Inborn Errors") | 4,172 |
| 20 | (MH "Streptococcal Infections+") | 11,681 |
| 21 | (MH "Hepatitis+") | 31,223 |
| 22 | (MH "Tuberculosis+") | 25,760 |
| 23 | (MH "Syphilis+") | 4,762 |
| 24 | (MH "Human Immunodeficiency Virus+") | 11,212 |
| 25 | (MH "Cicatrix+") | 6,911 |
| 26 | (MH "Keloid") | 642 |
| 27 | OR 8-26 | 3,324,304 |
| 28 | 7 AND 27 Limited 20150101 – 20230831; English Language | **158** |

#### CINAHL (EBSCO) 07.08.2023

#### Total References Added to Endnote

| Database | References |
| --- | --- |
| Medline (Ovid) | 412 |
| EMBASE (Ovid) | 708 |
| Ovid EMCARE | 20 |
| AMED (Ovid) | 0 |
| CINAHL (EBSCO) | 158 |
| **TOTAL** | **1298** |
| Duplicates Identified in Endnote | 412 |
| **New TOTAL** | **886** |

## APPENDIX 3: Websites searched for grey literature

| Websites |
| --- |
| Academy of Medical Sciences<br>https://acmedsci.ac.uk/ |
| American Academy of Allergy, Asthma & Immunology<br>https://www.aaaai.org/ |
| American Academy of Dermatology<br>https://www.aad.org/ |
| American College of Allergy, Asthma & Immunology<br>https://acaai.org/ |
| Association of Medical Microbiology and Infectious Disease Canada<br>https://ammi.ca/en/ |
| Australasian College of Dermatologists<br>https://www.dermcoll.edu.au/ |
| Australasian Society of Clinical Immunology and Allergy<br>https://www.allergy.org.au/ |
| Australian Medical Association<br>https://www.ama.com.au/ |
| British Association of Dermatologists<br>https://www.bad.org.uk/ |
| British HIV Association<br>https://www.bhiva.org/ |
| British Society for Haematology<br>https://b-s-h.org.uk/ |
| British Society for Immunology<br>https://www.immunology.org/ |
| Canadian Medical Association<br>https://www.cma.ca/ |
| Centers for Disease Control and Prevention<br>https://www.cdc.gov/ |
| European Academy of Allergy and Clinical Immunology<br>https://eaaci.org/ |
| European Centre for Disease Prevention and Control<br>https://www.ecdc.europa.eu/ |
| European Hematology Association<br>https://ehaweb.org/ |
| Health Education and Improvement Wales<br>https://heiw.nhs.wales/ |
| Health Education England<br>https://www.hee.nhs.uk/ |
| Infectious Diseases Society of America<br>https://www.idsociety.org/ |
| Medical Council of New Zealand<br>https://www.mcnz.org.nz/ |
| National Institute for Health and Care Excellence<br>https://www.nice.org.uk/ |
| New Zealand Dermatological Society<br>https://www.nzdsi.org/ |
| Royal Australasian College of Physicians<br>https://www.racp.edu.au/ |
| Royal College of General Practitioners<br>https://www.rcgp.org.uk/ |
| Royal College of Physicians<br>https://www.rcplondon.ac.uk/ |
| Royal College of Physicians and Surgeons of Canada<br>https://www.royalcollege.ca/ |
| Royal College of Physicians and Surgeons of Glasgow<br>https://rcpsg.ac.uk/ |
| Royal College of Physicians of Edinburgh<br>https://www.rcpe.ac.uk/ |
| Royal New Zealand College of General Practitioners<br>https://www.rnzcp.org.nz/ |
| Royal Society for Public Health (RSPH)<br>https://www.rsph.org.uk/ |
| Royal Society of Medicine<br>https://www.rsm.ac.uk |
| World Health Organisation<br>www.who.int |
| Public Health Wales<br>https://phw.nhs.wales/ |
| UK Health Security Agency<br>https://researchportal.ukhsa.gov.uk/ |
| Office for Health Improvement & Disparities<br>https://www.gov.uk/government/organisations/office-for-health-improvement-and-disparities<br>Filtered for research |
| Public Health Scotland<br>https://publichealthscotland.scot/ |
| Public Health Northern Ireland<br>https://www.publichealth.hscni.net/ |
| US Food and Drug Administration (FDA)<br>https://www.fda.gov/ |
| Google Advanced Search<br>https://www.google.co.uk/advanced_search |
| Association of Professional Piercers<br>https://safepiercing.org/ |
| The United Kingdom's Association Of Professional Piercers<br>https://www.ukapp.org.uk/ |
| Accreditation Commission for Acupuncture and Herbal Medicine<br>https://acahm.org/ |
| American Association of Acupuncture and Oriental Medicine<br>https://www.aaaomonline.org/ |
| British Acupuncture Council<br>https://acupuncture.org.uk/ |
| International Academy of Medical Acupuncture, Inc.<br>https://iama.edu/ |
| National Certification Commission for Acupuncture and Oriental Medicine<br>https://www.nccaom.org/ |
| Society for Acupuncture Research<br>https://www.acupunctureresearch.org/ |

## APPENDIX 4: Excluded studies

**Table 18.** Studies excluded at full-text screening.

| Full citation | Exclusion reason(s) |
| --- | --- |
| **Q1a. Tattooing (n=21)** |  |
| Dieckmann R, Boone I, Brockmann SO, et al. (2016). The risk of bacterial infection after tattooing – A systematic review of the literature. *Deutsches Arzteblatt International*. 113(40): 665-71.<br>https://dx.doi.org/10.3238/arztebl.2016.0665 | No critical appraisal |
| Huisman S, van der Bent SAS, Maijer KI, et al. (2020). Cutaneous non-allergic complications in tattoos: An overview of the literature. *Presse Medicale*. 49(4): 104049.<br>https://dx.doi.org/10.1016/j.lpm.2020.104049 | Wrong study design (narrative review), <2 databases searched |
| Ibraheim MK, Desai M, Tawfik M, et al. (2023). microblading-induced granulomatous reaction: case report and review of the literature. *American Journal of Dermatopathology*. 45(7): 487-91.<br>https://dx.doi.org/10.1097/DAD.0000000000002449 | Wrong study design (narrative review), <2 databases searched |
| Khodadost M, Maajani K, Arabsalmani M, et al. (2017). Is tattooing a risk factor for hepatitis C transmission?: An updated systematic review and meta-analysis. *Hepatitis Monthly*. 17.<br>https://dx.doi.org/10.5812/hepatmon.14308 | No critical appraisal |
| Kiszla BM, Elmets CA, Mayo TT. (2023). Quantitative analysis of restricted metals and metalloids in tattoo inks: A systematic review and meta-analysis. *Chemosphere*. 313: 137291.<br>https://dx.doi.org/10.1016/j.chemosphere.2022.137291 | No critical appraisal |
| Kluger N, Andraud M, Lartigau-Roussin C, et al. (2021). The Koebner phenomenon on tattoos and piercings in a patient with cutaneous lupus: a case report and review of the literature. *Acta Dermatovenereologica Alpina, Panonica et Adriatica*. 30(1): 43-6. | Wrong study design (narrative review), <2 databases searched |
| Laux P, Tralau T, Tentschert J, et al. (2016). A medical-toxicological view of tattooing. *Lancet*. 387(10016): 395-402.<br>[https://dx.doi.org/10.1016/S0140-6736(15)60215-X](https://dx.doi.org/10.1016/S0140-6736%2815%2960215-X) | Wrong study design (narrative review), <2 databases searched |
| Lim SH, Lee S, Lee YB, et al. (2022). Increased prevalence of transfusion-transmitted diseases among people with tattoos: A systematic review and metaanalysis. *PLoS ONE*. 17.<br>https://dx.doi.org/10.1371/journal.pone.0262990 | Wrong outcome |
| Mudedla S, Avendano EE, Raman G. (2015). Non-tuberculous mycobacterium skin infections after tattooing in healthy individuals: a systematic review of case reports. *Dermatology Online Journal*. 21(6): 16. | No critical appraisal |
| Munoz-Ortiz J, Gomez-Lopez MT, Echeverry-Hernandez P, et al. (2021). Dermatological and ophthalmological inflammatory, infectious, and tumoral tattoo-related reactions: a systematic review. *Permanente Journal*. 25(5): 26.<br>https://dx.doi.org/10.7812/TPP/20.225 | No critical appraisal |
| Negi S, Bala L, Shukla S, et al. (2022). Tattoo inks are toxicological risks to human health: A systematic review of their ingredients, fate inside skin, toxicity due to polycyclic aromatic hydrocarbons, primary aromatic amines, metals, and overview of regulatory frameworks. *Toxicology & Industrial Health*. 38(7): 417-34.<br>https://dx.doi.org/10.1177/07482337221100870 | Wrong study design (narrative review) |
| Patel M, Cobbs CG. (2015). Infections from Body Piercing and Tattoos. *Microbiology Spectrum*. 3(6): 12.<br>https://dx.doi.org/10.1128/microbiolspec.IOL5-0016-2015 | Wrong study design (narrative review), <2 databases searched |
| Rahbarinejad Y, Guio-Aguilar P, Vu AN, et al. (2023). Pathogenesis, diagnosis and management of squamous cell carcinoma and pseudoepithelial hyperplasia secondary to red ink tattoo: a case series and Review. *Journal of Clinical Medicine*. 12(6): 21.<br>https://dx.doi.org/10.3390/jcm12062424 | Wrong study design (narrative review), <2 databases searched |
| Rello J, Tejada S, Campogiani L, et al. (2022). Systemic infections associated with tattoos or permanent makeup: A systematic review. *Medicina Clinica*. 158(4): 159-66.<br>https://dx.doi.org/10.1016/j.medcli.2021.01.014 | <2 databases searched, no critical appraisal |
| Ricci F, Fania L, Antonelli F, et al. (2022). Melanoma on tattoos: a case study and review of the literature. *European Journal of Dermatology*. 32(6): 703-8.<br>https://dx.doi.org/10.1684/ejd.2022.4353 | Wrong study design (narrative review), <2 databases searched |
| Thomas S, Gouk C, Jayasakeera N, et al. (2016). The Sequelae of Metallosis Resulting in Skin Pigmentation and Tattooing: A Case Presentation and Literature Review. *The Surgery Journal*. 2(4): e143-e6.<br>https://dx.doi.org/10.1055/s-0036-1596060 | Wrong study design (narrative review), no critical appraisal |
| Thum CK, Biswas A. (2015). Inflammatory complications related to tattooing: a histopathological approach based on pattern analysis. *American Journal of Dermatopathology*. 37(1): 54-66.<br>https://dx.doi.org/10.1097/DAD.0b013e3182974558 | Wrong study design (narrative review), <2 databases searched |
| Tran NT, Dubost C, Baggio S, et al. (2018). Safer tattooing interventions in prisons: a systematic review and call to action. *BMC Public Health*. 18(1): 1015.<br>https://dx.doi.org/10.1186/s12889-018-5867-x | Wrong outcome |
| Van Bresse MF, Van Waerebeek K, Duignan PJ. (2022). tattoo skin disease in cetacea: a review, with new cases for the Northeast Pacific. *Animals* [Electronic Resource]. 12(24): 18.<br>https://dx.doi.org/10.3390/ani12243581 | Wrong population, wrong study design |
| Van Remoortel H, Moorkens D, Avau B, et al. (2019). Is there a risk of transfusion-transmissible infections after percutaneous needle treatments in blood donors? A systematic review and meta-analysis. *Vox Sanguinis*. 114(4): 297-309.<br>https://dx.doi.org/10.1111/vox.12780 | Wrong outcome |
| Weis KT, Schreiver I, Siewert K, et al. (2021). Tattoos – more than just colored skin? Searching for tattoo allergens. *Journal der Deutschen Dermatologischen Gesellschaft*. 19(5): 657-69. https://dx.doi.org/10.1111/ddg.14436 | Wrong study design (narrative review), <2 databases searched |
| **Q1b. Permanent make-up (n=22)** |  |
| Bruhn C. (2016). Ornaments with risks: Health risks due to tattoos, permanent make-up and tattoo removal. *Deutsche Apotheker Zeitung*. 156: 395-402 | Foreign language |
| Daruwalla SB, Dhurat RS, Hamid SAT. (2022). All that a dermatotrichologist needs to know about hair camouflage: a comprehensive review. *International Journal of Trichology*. 14(3): 77-83.<br>https://dx.doi.org/10.4103/ijt.ijt_6_21 | Wrong outcome, wrong population |
| De Cuyper C. (2015). Complications of cosmetic tattoos. *Current Problems in Dermatology*. 48: 61-70.<br>https://dx.doi.org/10.1159/000369188 | Wrong publication type |
| De Cuyper C. (2020a). How to advise a patient who wants a tattoo? *Presse Medicale*. 49(4): 104048.<br>https://dx.doi.org/10.1016/j.lpm.2020.104048 | Wrong publication type |
| De Cuyper C. (2020b). Tattoo allergy. Can we identify the allergen? *Presse Medicale*. 49(4): 104047. https://dx.doi.org/10.1016/j.lpm.2020.104047 | Wrong publication type |
| Dirks M. (2022). Formulation and production of tattoo ink stock products for cosmetic and medical applications: pigment raw materials, ingredients, auxiliary substances and contaminants. *Current Problems in Dermatology*. 56: 23-9.<br>https://dx.doi.org/10.1159/000521863 | Wrong publication type, not available |
| Giulbudagian M, Schreiver I, Singh AV, et al. (2020). Safety of tattoos and permanent make-up: a regulatory view. *Archives of Toxicology*. 94(2): 357-69.<br>https://dx.doi.org/10.1007/s00204-020-02655-z | Wrong publication type |
| Hvas D, Serup J. (2022). Microblading technique for tattooing of "hairstrokes" that simulate natural hair: eyebrow tattooing and correction of medical conditions. *Current Problems in Dermatology*. 56: 141-54.<br>https://dx.doi.org/10.1159/000529810 | Wrong publication type, not available |
| Kluger N. (2019). An update on cutaneous complications of permanent tattooing. *Expert Review of Clinical Immunology*. 15(11): 1135-43.<br>https://dx.doi.org/10.1080/1744666X.2020.1676732 | Wrong intervention |
| Kluger N. (2022). delayed granulomatous reaction after eyebrow microblading. *Dermatologic Surgery*. 48(4): 472-3.<br>https://dx.doi.org/10.1097/DSS.0000000000003361 | Wrong publication type |
| Marwah MK, Kerure AS, Marwah GS. (2021). Microblading and the science behind it. *Indian Dermatology Online Journal*. 12(1): 6-11.<br>https://dx.doi.org/10.4103/idoj.IDOJ_230_20 | Wrong study design (unpicked narrative review) |
| Park JH, Ho YH, Manonukul K. (2022). Natural results of scalp micropigmentation: a review. *Journal of Cosmetic Dermatology*. 21(11): 5509-13.<br>https://dx.doi.org/10.1111/jocd.15297 | Wrong outcome, wrong study design |
| Portilla Maya N, Kempf W, Perez Munoz N, et al. (2021). Histopathologic spectrum of findings associated with tattoos: multicenter study series of 230 cases. *American Journal of Dermatopathology*. 43(8): 543-53.<br>https://dx.doi.org/10.1097/DAD.0000000000001695 | Wrong intervention |
| Rello J, Tejada S, Campogiani L, et al. (2022). Systemic infections associated with tattoos or permanent makeup: a systematic review. *Medicina Clinica*. 158(4): 159-66.<br>https://dx.doi.org/10.1016/j.medcli.2021.01.014 | Wrong study design (unpicked narrative review) |
| Schubert S, Kluger N, Schreiber I. (2023). Hypersensitivity to permanent tattoos: Literature summary and comprehensive review of patch tested tattoo patients 1997-2022. *Contact Dermatitis*. 88(5): 331-50.<br>https://dx.doi.org/10.1111/cod.14291 | Wrong study design (unpicked narrative review) |
| Serup J. (2022). Technical and Clinical Complications of Cosmetic Tattooing. *Current Problems in Dermatology*. 56: 225-44.<br>https://dx.doi.org/10.1159/000526048 | Wrong study design (unpicked narrative review) |
| Shaji M, Merin KA, Kameswaran R. (2022). A Review on Cosmetics Causing Contact Urticaria. *Indian Journal of Dermatology*. 67(4): 392-8.<br>https://dx.doi.org/10.4103/ijd.ijd_1090_20 | Wrong intervention, wrong study design |
| Spurr A, Hanna N, Colantonio S. (2022). Cutaneous sarcoidosis in eyebrows cosmetically pigmented with microblading method: A case report and review of the literature. *SAGE Open Medical Case Reports*. 10: 2050313X221117720.<br>https://dx.doi.org/10.1177/2050313X221117720 | Wrong study design (unpicked narrative review) |
| Tomita S, Mori K, Yamazaki H, et al. (2021). Complications of permanent makeup procedures for the eyebrow and eyeliner. *Medicine*. 100(18): e25755.<br>https://dx.doi.org/10.1097/MD.00000000000025755 | Wrong setting |
| van der Bent SAS, Rauwerdink D, Oyen EMM, et al. (2021). Complications of tattoos and permanent makeup: overview and analysis of 308 cases. *Journal of Cosmetic Dermatology*. 20(11): 3630-41. https://dx.doi.org/10.1111/jocd.14498 | Data pertaining to SPMU cannot be extracted |
| Verdier C. (2015). Surveillance of tattoo-related adverse events by the EU RAPEX system and by national monitoring. *Current Problems in Dermatology*. 48: 210-7.<br>https://dx.doi.org/10.1159/000369230 | Wrong publication type, wrong study design, |
| Verhulst L, Goossens A. (2016). Cosmetic components causing contact urticaria: a review and update. *Contact Dermatitis*. 75(6): 333-44.<br>https://dx.doi.org/10.1111/cod.12679 | Wrong intervention, wrong study design |
| **Q2. Body piercing (n=11)** |  |
| Domian N, Kasacka I. (2017). Local and systemic disorders caused by the presence of foreign bodies in the oral cavity. *Dental and Medical Problems*. 54: 267-72.<br>https://dx.doi.org/10.17219/dmp/75553 | Wrong study design (narrative review), <2 databases searched, no critical appraisal |
| Kluger N, Andraud M, Lartigau-Roussin C, et al. (2021). The Koebner phenomenon on tattoos and piercings in a patient with cutaneous lupus: a case report and review of the literature. *Acta Dermatovenereologica Alpina, Panonica et Adriatica*. 30(1): 43-6. | Wrong study design (narrative review), <2 databases searched, no critical appraisal |
| Lee B, Vangipuram R, Peterson E, et al. (2018). Complications associated with intimate body piercings. *Dermatology Online Journal*. 24.<br>https://pubmed.ncbi.nlm.nih.gov/30261561/ | Wrong study design (narrative review), <2 databases searched, no critical appraisal |
| Mclsaac KE. (2016). Incorrect application of epidemiologic concepts in The Incidence of Complications associated with Lip and/or Tongue piercings: a systematic review. *International Journal of Dental Hygiene*. 14(1): 74-5.<br>https://dx.doi.org/10.1111/ijdh.12180 | Wrong publication type (letter to the editor) |
| Offen E, Allison JR. (2022). Do oral piercings cause problems in the mouth? Evidence-Based Dentistry. 23(3): 126-7.<br>https://dx.doi.org/10.1038/s41432-022-0816-z | Wrong publication type (commentary) |
| Patel M, Cobbs CG. (2015). Infections from Body Piercing and Tattoos. *Microbiology Spectrum*. 3(6): 12.<br>https://dx.doi.org/10.1128/microbiolspec.IOL5-0016-2015 | Wrong study design (narrative review), <2 databases searched, no critical appraisal |
| Saccomanno S, Ieria I, Manenti RJ, et al. (2021). Complications of oral piercing: a review of the literature and two case reports. *Journal of Biological Regulators & Homeostatic Agents*. 35(3): 167-78.<br>https://dx.doi.org/10.23812/21-3supp1-19 | Wrong study design (narrative review), <2 databases searched, no critical appraisal |
| Sosin M, Weissler JM, Pulcrano M, et al. (2015). Transcartilaginous ear piercing and infectious complications: a systematic review and critical analysis of outcomes. *Laryngoscope*. 125(8): 1827-34.<br>https://dx.doi.org/10.1002/lary.25238 | <2 databases searched, no critical appraisal |
| Van Remoortel H, Moorkens D, Avau B, et al. (2019). Is there a risk of transfusion-transmissible infections after percutaneous needle treatments in blood donors? A systematic review and meta-analysis. *Vox Sanguinis*. 114(4): 297-309.<br>https://dx.doi.org/10.1111/vox.12780 | Wrong outcome |
| Vane SCS. (2016). Oral and perioral piercings: An analysis of complications, management and use of oral hygiene techniques. *Research Journal of Pharmacy and Technology*. 8: 1007-10.<br>https://dx.doi.org/10.5958/0974-360X.2015.00170.5 | Wrong study design (narrative review), <2 databases searched, no critical appraisal |
| Yang S, Wang D, Zhang Y, et al. (2015). Transmission of hepatitis B and C virus infection through body piercing: a systematic review and meta-analysis. *Medicine*. 94(47): e1893.<br>https://dx.doi.org/10.1097/MD.0000000000001893 | Wrong outcome |
| **Q3. Acupuncture (n=2)** |  |
| Nielsen A, Gereau S, Tick H. (2020). Risks and safety of extended auricular therapy: a review of reviews and case reports of adverse events. *Pain Medicine*. 21(6): 1276-93.<br>https://dx.doi.org/10.1093/pm/pnz379 | No critical appraisal |
| Paley CA, Johnson MI. (2019). Acupuncture for the relief of chronic pain: a synthesis of systematic reviews. *Medicina*. 56(1): 24.<br>https://dx.doi.org/10.3390/medicina56010006 | No critical appraisal, wrong outcome |
| **Q4. Electrolysis (n=20)** |  |
| Butler SM, Smith NK, Collazo E, et al. (2015). Pubic hair preferences, reasons for removal, and associated genital symptoms: comparisons between men and women. *Journal of Sexual Medicine*. 12(1): 48-58.<br>https://dx.doi.org/10.1111/jsm.12763 | Wrong outcome |
| Cohen PR. (2020). Hypertrichosis and genital rejuvenation: cutaneous adverse events associated with pubic hair grooming. *SKINmed*. 18(5): 297-9. | Wrong intervention, wrong study design |
| Gaither TW, Fergus K, Sutcliffe S, et al. (2020). Pubic hair grooming and sexually transmitted infections: a clinic-based cross-sectional survey. *Sexually Transmitted Diseases*. 47(6): 419-25. | Wrong outcome |
| Gaither TW, Truesdale M, Harris CR, et al. (2015). The influence of sexual orientation and sexual role on male grooming-related injuries and infections. *Journal of Sexual Medicine*. 12(3): 631-40.<br>https://dx.doi.org/10.1111/jsm.12780 | Wrong outcome |
| Hodges AL, Holland AC. (2017). Prevention and treatment of injuries and infections related to pubic hair removal. *Nursing for women's health*. 21: 313-7.<br>https://dx.doi.org/10.1016/j.nwh.2017.06.001 | Wrong intervention, wrong study design |
| Kang CN, Shah M, Lynde C, et al. (2021). hair removal practices: a literature review. *Skin Therapy Letter*. 26(5): 6-11 | Wrong study design (unpicked narrative review) |
| Luster J, Turner AN, Henry JP, Jr., et al. (2019). Association between pubic hair grooming and prevalent sexually transmitted infection among female university students. *PLoS ONE* [Electronic Resource]. 14(9): e0221303. https://dx.doi.org/10.1371/journal.pone.0221303 | Wrong intervention |
| Matheson E, Bain J. (2019). Hirsutism in women. *American Family Physician*. 100(3): 168-75. | Wrong study design (unpicked narrative review) |
| Mezin-Sarbu E, Wohlrab J. (2023). Epilation and depilation in the genital area – motivation, methods, risks and recommendations from a dermatological point of view. *Journal der Deutschen Dermatologischen Gesellschaft*. 21(5): 455-62 https://dx.doi.org/10.1111/ddg.14993 | Wrong study design (unpicked systematic review), not focused on electrolysis |
| Molgo M, Moll-Manzur C, Downey C. (2020). Follicular traction urticaria induced by electric epilation. *Cutis*. 105(5): E23-E4. | Wrong intervention |
| Osterberg EC, Gaither TW, Awad MA, et al. (2017). Correlation between pubic hair grooming and STIs: results from a nationally representative probability sample. *Sexually Transmitted Infections*. 93(3): 162-6. | Wrong outcome |
| Pany A, Klang V, Brunner M, et al. (2019). Effect of physical and chemical hair removal methods on skin barrier function in vitro: consequences for a hydrophilic model permanent. *Skin Pharmacology & Physiology*. 32(1): 8-21. https://dx.doi.org/10.1159/000493168 | Wrong outcome, wrong population |
| Shukalek C, Parsons L, Somayaji R. (2017). Delving into skin and soft tissue infections (SSTI) Part II: Focus on superficial infections. *World Council of Enterostomal Therapists Journal*. 37(3): 20-4. | Full text not available |
| Staszczuk M. (2020). Just Zap It: Electrolysis is a permanent alternative to waxing and sugaring. *Skin Deep*. 32-3. | Wrong publication type |
| Swain TA, Tully AS, Redford T, et al. (2016). Hair removal-related injuries in the United States, 1991-2014. *Journal of Cosmetic Dermatology*. 15(4): 444-51. https://dx.doi.org/10.1111/jocd.12283 | Wrong intervention |
| Trivedi MK, Kroumpouzou G, Murase JE. (2017). A review of the safety of cosmetic procedures during pregnancy and lactation. *International Journal of Women's Dermatology*. 3(1): 6-10. https://dx.doi.org/10.1016/j.ijwd.2017.01.005 | Wrong outcome, wrong study design |
| Truesdale MD, Osterberg EC, Gaither TW, et al. (2017). prevalence of pubic hair grooming-related injuries and identification of high-risk individuals in the United States. *JAMA Dermatology*. 153(11): 1114-21. https://dx.doi.org/10.1001/jamadermatol.2017.2815 | No breakdown of outcomes by type of hair removal |
| Williamson H. (2015). Social pressures and health consequences associated with body hair removal. *Journal of Aesthetic Nursing*. 4(3): 131-3. | Wrong study design |
| Wu Y, Dai Y, Wang T, et al. (2022). The application of electrolysis of depigmented hair using a trichiasis electrolyzer combined with single hair follicle transplantation for the treatment of vitiligo-associated leukotrichia. *Dermatologic Therapy*. 35(5): e15400. https://dx.doi.org/10.1111/dth.15400 | Wrong setting, wrong intervention |
| Yuan N, Feldman AT, Chin P, et al. (2022). Comparison of Permanent Hair Removal Procedures before Gender-Affirming Vaginoplasty: Why We Should Consider Laser Hair Removal as a First-Line Treatment for Patients Who Meet Criteria. *Sexual Medicine*. 10(5): 100545. https://dx.doi.org/10.1016/j.esxm.2022.100545 | Wrong setting, wrong intervention |

## APPENDIX 5: Table of unique primary studies of body piercing

**Table 19.**
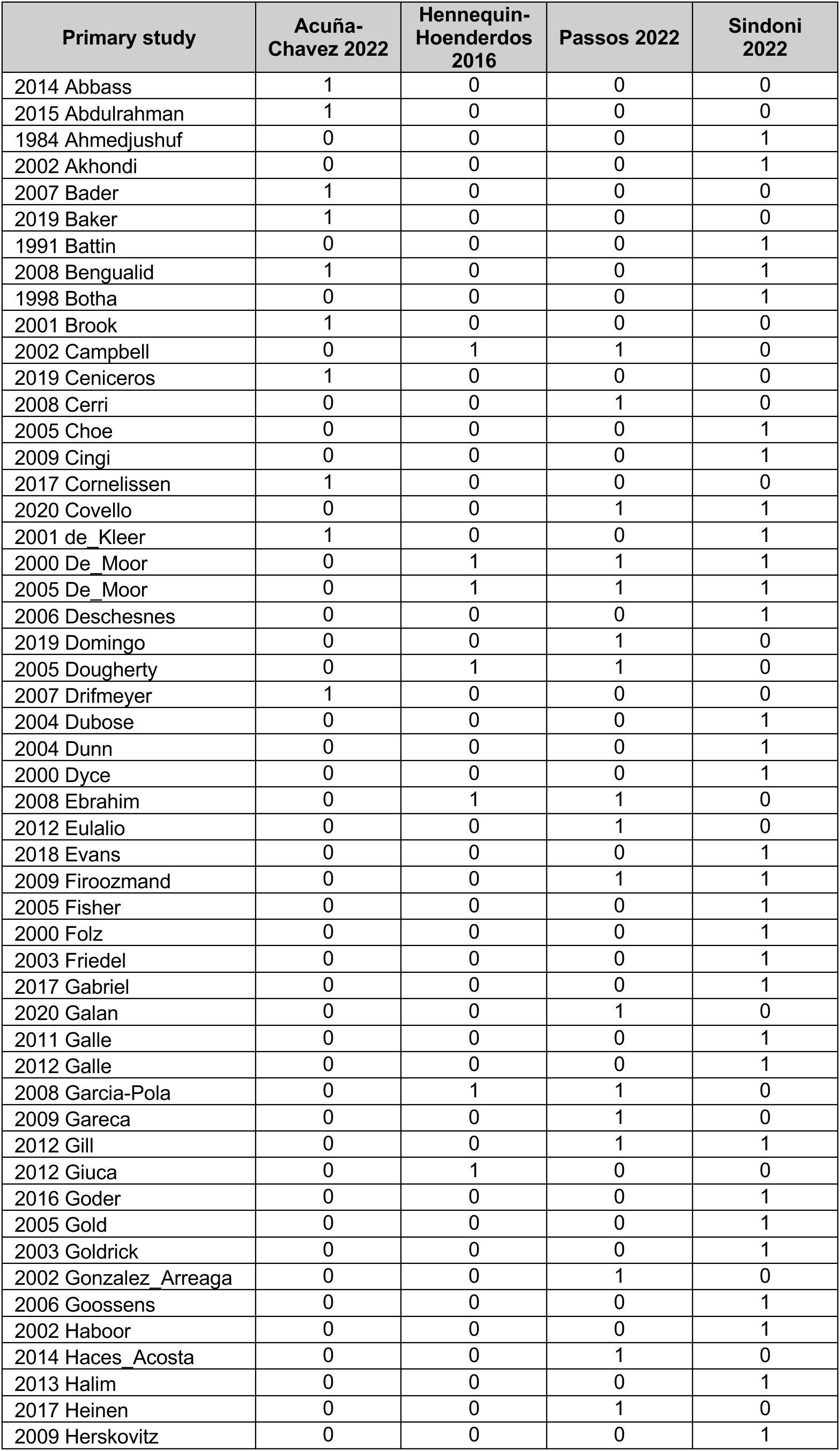
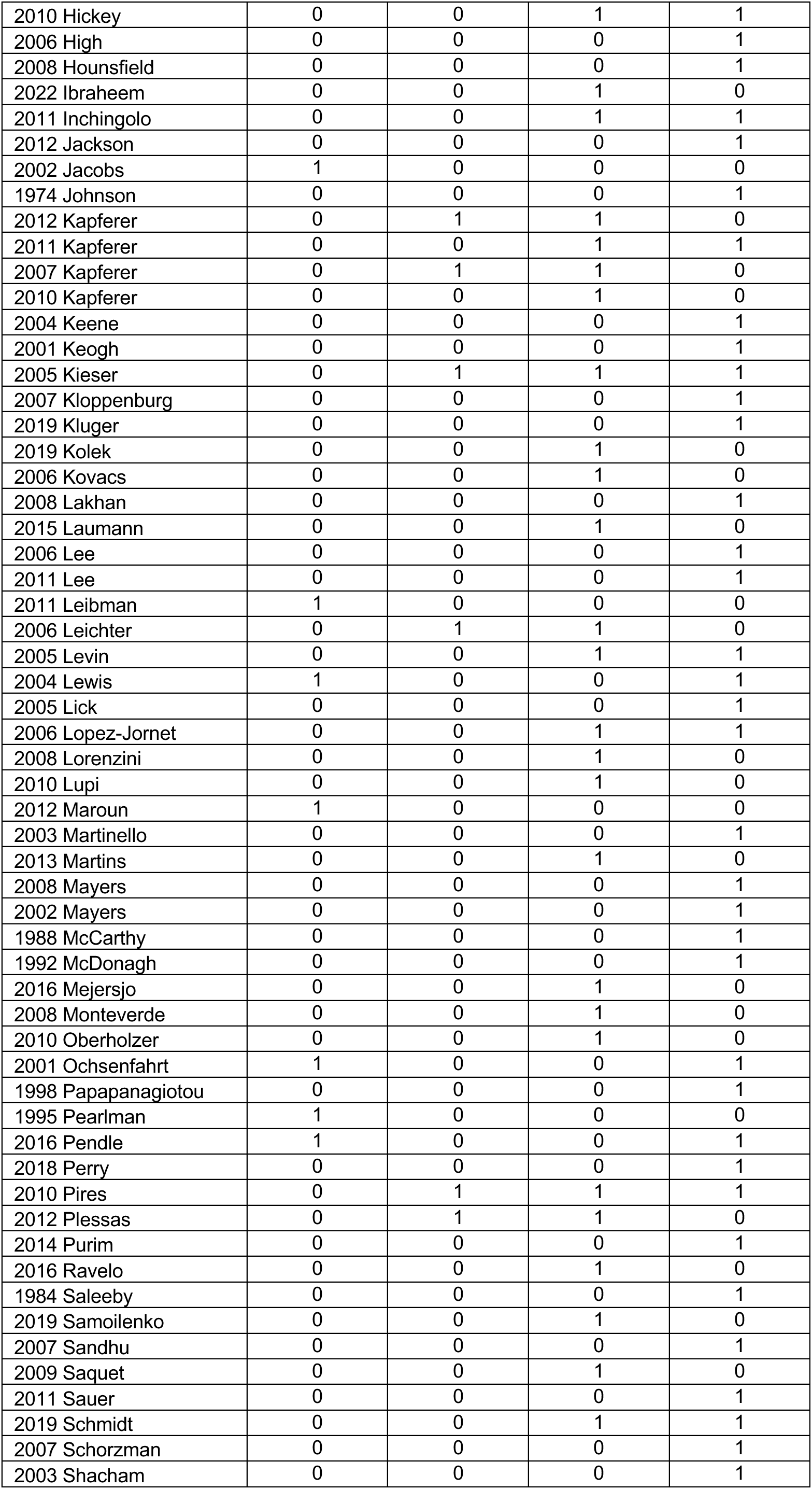
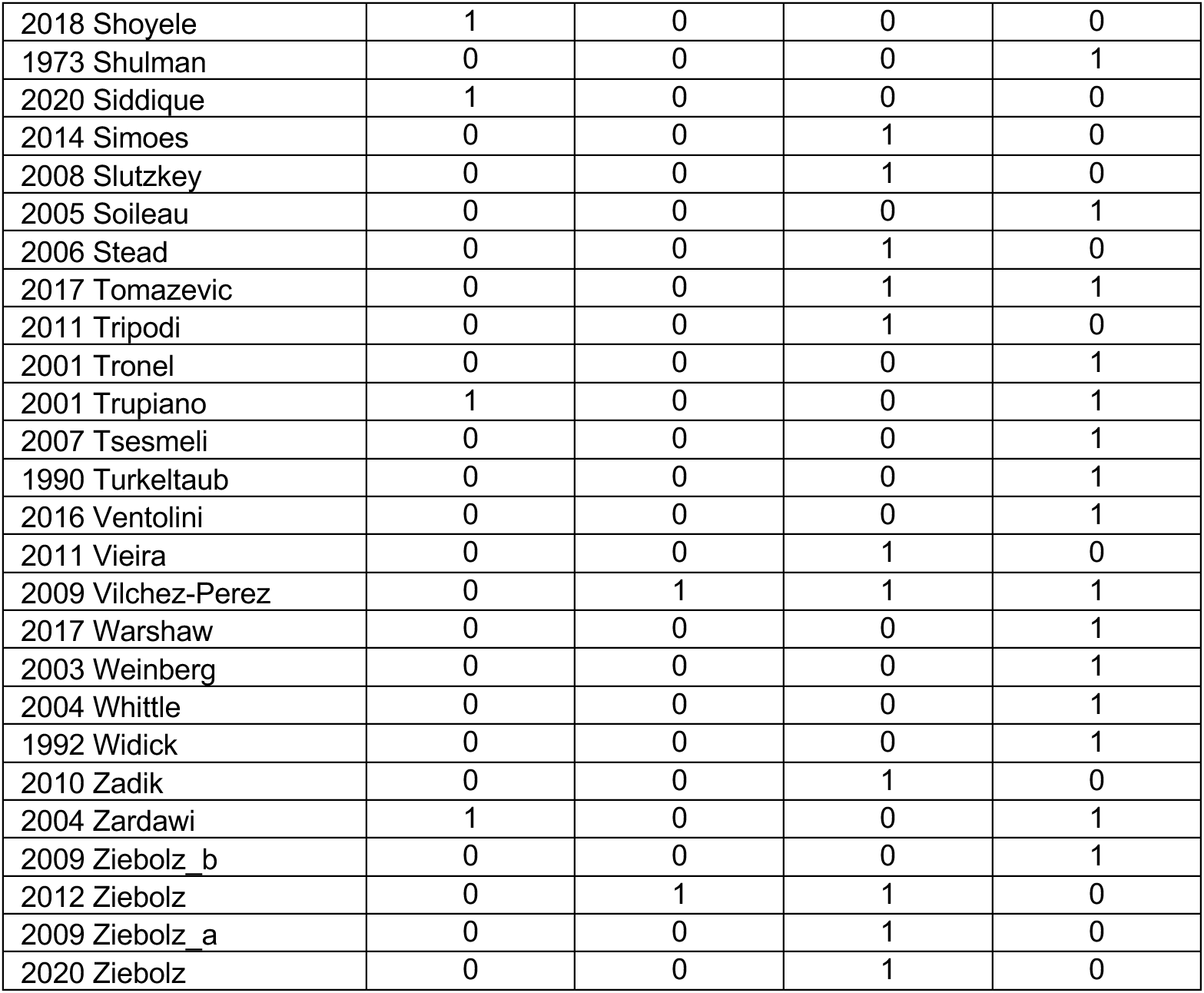
Unique primary studies from included SRs on body piercing.

| Primary study | Acuña-Chavez 2022 | Hennequin-Hoenderdos 2016 | Passos 2022 | Sindoni 2022 |
| --- | --- | --- | --- | --- |
| 2014 Abbass | 1 | 0 | 0 | 0 |
| 2015 Abdulrahman | 1 | 0 | 0 | 0 |
| 1984 Ahmedjushuf | 0 | 0 | 0 | 1 |
| 2002 Akhondi | 0 | 0 | 0 | 1 |
| 2007 Bader | 1 | 0 | 0 | 0 |
| 2019 Baker | 1 | 0 | 0 | 0 |
| 1991 Battin | 0 | 0 | 0 | 1 |
| 2008 Bengualid | 1 | 0 | 0 | 1 |
| 1998 Botha | 0 | 0 | 0 | 1 |
| 2001 Brook | 1 | 0 | 0 | 0 |
| 2002 Campbell | 0 | 1 | 1 | 0 |
| 2019 Cenicerós | 1 | 0 | 0 | 0 |
| 2008 Cerri | 0 | 0 | 1 | 0 |
| 2005 Choe | 0 | 0 | 0 | 1 |
| 2009 Cingi | 0 | 0 | 0 | 1 |
| 2017 Cornelissen | 1 | 0 | 0 | 0 |
| 2020 Covello | 0 | 0 | 1 | 1 |
| 2001 de Kleer | 1 | 0 | 0 | 1 |
| 2000 De Moor | 0 | 1 | 1 | 1 |
| 2005 De Moor | 0 | 1 | 1 | 1 |
| 2006 Deschesnes | 0 | 0 | 0 | 1 |
| 2019 Domingo | 0 | 0 | 1 | 0 |
| 2005 Dougherty | 0 | 1 | 1 | 0 |
| 2007 Drifmeyer | 1 | 0 | 0 | 0 |
| 2004 Dubose | 0 | 0 | 0 | 1 |
| 2004 Dunn | 0 | 0 | 0 | 1 |
| 2000 Dyce | 0 | 0 | 0 | 1 |
| 2008 Ebrahim | 0 | 1 | 1 | 0 |
| 2012 Eulalio | 0 | 0 | 1 | 0 |
| 2018 Evans | 0 | 0 | 0 | 1 |
| 2009 Firoozmand | 0 | 0 | 1 | 1 |
| 2005 Fisher | 0 | 0 | 0 | 1 |
| 2000 Folz | 0 | 0 | 0 | 1 |
| 2003 Friedel | 0 | 0 | 0 | 1 |
| 2017 Gabriel | 0 | 0 | 0 | 1 |
| 2020 Galan | 0 | 0 | 1 | 0 |
| 2011 Galle | 0 | 0 | 0 | 1 |
| 2012 Galle | 0 | 0 | 0 | 1 |
| 2008 Garcia-Pola | 0 | 1 | 1 | 0 |
| 2009 Gareca | 0 | 0 | 1 | 0 |
| 2012 Gill | 0 | 0 | 1 | 1 |
| 2012 Giuca | 0 | 1 | 0 | 0 |
| 2016 Goder | 0 | 0 | 0 | 1 |
| 2005 Gold | 0 | 0 | 0 | 1 |
| 2003 Goldrick | 0 | 0 | 0 | 1 |
| 2002 Gonzalez_Arreaga | 0 | 0 | 1 | 0 |
| 2006 Goossens | 0 | 0 | 0 | 1 |
| 2002 Haboor | 0 | 0 | 0 | 1 |
| 2014 Haces_Acosta | 0 | 0 | 1 | 0 |
| 2013 Halim | 0 | 0 | 0 | 1 |
| 2017 Heinen | 0 | 0 | 1 | 0 |
| 2009 Herskovitz | 0 | 0 | 0 | 1 |
| 2010 Hickey | 0 | 0 | 1 | 1 |
| 2006 High | 0 | 0 | 0 | 1 |
| 2008 Hounsfield | 0 | 0 | 0 | 1 |
| 2022 Ibraheem | 0 | 0 | 1 | 0 |
| 2011 Inchingolo | 0 | 0 | 1 | 1 |
| 2012 Jackson | 0 | 0 | 0 | 1 |
| 2002 Jacobs | 1 | 0 | 0 | 0 |
| 1974 Johnson | 0 | 0 | 0 | 1 |
| 2012 Kapferer | 0 | 1 | 1 | 0 |
| 2011 Kapferer | 0 | 0 | 1 | 1 |
| 2007 Kapferer | 0 | 1 | 1 | 0 |
| 2010 Kapferer | 0 | 0 | 1 | 0 |
| 2004 Keene | 0 | 0 | 0 | 1 |
| 2001 Keogh | 0 | 0 | 0 | 1 |
| 2005 Kieser | 0 | 1 | 1 | 1 |
| 2007 Kloppenburg | 0 | 0 | 0 | 1 |
| 2019 Kluger | 0 | 0 | 0 | 1 |
| 2019 Kolek | 0 | 0 | 1 | 0 |
| 2006 Kovacs | 0 | 0 | 1 | 0 |
| 2008 Lakhan | 0 | 0 | 0 | 1 |
| 2015 Laumann | 0 | 0 | 1 | 0 |
| 2006 Lee | 0 | 0 | 0 | 1 |
| 2011 Lee | 0 | 0 | 0 | 1 |
| 2011 Leibman | 1 | 0 | 0 | 0 |
| 2006 Leichter | 0 | 1 | 1 | 0 |
| 2005 Levin | 0 | 0 | 1 | 1 |
| 2004 Lewis | 1 | 0 | 0 | 1 |
| 2005 Lick | 0 | 0 | 0 | 1 |
| 2006 Lopez-Jornet | 0 | 0 | 1 | 1 |
| 2008 Lorenzini | 0 | 0 | 1 | 0 |
| 2010 Lupi | 0 | 0 | 1 | 0 |
| 2012 Maroun | 1 | 0 | 0 | 0 |
| 2003 Martinello | 0 | 0 | 0 | 1 |
| 2013 Martins | 0 | 0 | 1 | 0 |
| 2008 Mayers | 0 | 0 | 0 | 1 |
| 2002 Mayers | 0 | 0 | 0 | 1 |
| 1988 McCarthy | 0 | 0 | 0 | 1 |
| 1992 McDonagh | 0 | 0 | 0 | 1 |
| 2016 Mejersjo | 0 | 0 | 1 | 0 |
| 2008 Monteverde | 0 | 0 | 1 | 0 |
| 2010 Oberholzer | 0 | 0 | 1 | 0 |
| 2001 Ochsenfahrt | 1 | 0 | 0 | 1 |
| 1998 Papapanagiotou | 0 | 0 | 0 | 1 |
| 1995 Pearlman | 1 | 0 | 0 | 0 |
| 2016 Pendle | 1 | 0 | 0 | 1 |
| 2018 Perry | 0 | 0 | 0 | 1 |
| 2010 Pires | 0 | 1 | 1 | 1 |
| 2012 Plessas | 0 | 1 | 1 | 0 |
| 2014 Purim | 0 | 0 | 0 | 1 |
| 2016 Ravelo | 0 | 0 | 1 | 0 |
| 1984 Saleeby | 0 | 0 | 0 | 1 |
| 2019 Samoilenko | 0 | 0 | 1 | 0 |
| 2007 Sandhu | 0 | 0 | 0 | 1 |
| 2009 Saquet | 0 | 0 | 1 | 0 |
| 2011 Sauer | 0 | 0 | 0 | 1 |
| 2019 Schmidt | 0 | 0 | 1 | 1 |
| 2007 Schorzman | 0 | 0 | 0 | 1 |
| 2003 Shacham | 0 | 0 | 0 | 1 |
| 2018 Shoyele | 1 | 0 | 0 | 0 |
| 1973 Shulman | 0 | 0 | 0 | 1 |
| 2020 Siddique | 1 | 0 | 0 | 0 |
| 2014 Simoes | 0 | 0 | 1 | 0 |
| 2008 Slutzkey | 0 | 0 | 1 | 0 |
| 2005 Soileau | 0 | 0 | 0 | 1 |
| 2006 Stead | 0 | 0 | 1 | 0 |
| 2017 Tomazevic | 0 | 0 | 1 | 1 |
| 2011 Tripodi | 0 | 0 | 1 | 0 |
| 2001 Tronel | 0 | 0 | 0 | 1 |
| 2001 Trupiano | 1 | 0 | 0 | 1 |
| 2007 Tsesmeli | 0 | 0 | 0 | 1 |
| 1990 Turkeltaub | 0 | 0 | 0 | 1 |
| 2016 Ventolini | 0 | 0 | 0 | 1 |
| 2011 Vieira | 0 | 0 | 1 | 0 |
| 2009 Vilchez-Perez | 0 | 1 | 1 | 1 |
| 2017 Warshaw | 0 | 0 | 0 | 1 |
| 2003 Weinberg | 0 | 0 | 0 | 1 |
| 2004 Whittle | 0 | 0 | 0 | 1 |
| 1992 Widick | 0 | 0 | 0 | 1 |
| 2010 Zadik | 0 | 0 | 1 | 0 |
| 2004 Zardawi | 1 | 0 | 0 | 1 |
| 2009 Ziebolz_b | 0 | 0 | 0 | 1 |
| 2012 Ziebolz | 0 | 1 | 1 | 0 |
| 2009 Ziebolz_a | 0 | 0 | 1 | 0 |
| 2020 Ziebolz | 0 | 0 | 1 | 0 |

## The Health and Care Research Wales Evidence Centre

Our dedicated team works together with Welsh Government, the NHS, social care, research institutions and the public to deliver vital research to tackle health and social care challenges facing Wales.

Funded by Welsh Government, through Health and Care Research Wales, the Evidence Centre answers key questions to improve health and social care policy and provision across Wales.

Along with our collaborating partners, we conduct reviews of existing evidence and new research, to inform policy and practice needs, with a focus on ensuring real-world impact and public benefit that reaches everyone.

**Director:** Professor Adrian Edwards

**Associate Directors:** Dr Alison Cooper, Dr Natalie Joseph-Williams, Dr Ruth Lewis

## Abbreviations

CI: Confidence Interval
ER: Event Rate
GRADE: Grading of Recommendations, Assessment, Development and Evaluation
HIV: Human Immunodeficiency Virus
OR: Odds Ratio
RR: Relative Risk
SR: Systematic Review
UK: United Kingdom
USA: United States of America

## Footnotes

1 There is a discrepancy relating to the number of SRs reporting infections in the text of the Xu et al. (2023) paper and in Table S1 (characteristics of the included studies) provided in supplementary materials to the paper. According to the paper, the number is 19 SRs. However, only 11 SRs reporting infections were identified in Table S1.

2 Multiple non-critical weaknesses may diminish confidence in the review and it may be appropriate to move the overall appraisal down from moderate to low quality.

3 Searched for reviews of both tattooing and semi-permanent make-up

4 See Table 18 in Appendix 4 for detail

5 1 NR identified and unpicked

6 Overall quality was assessed on four critical domains which relate to Q3, Q4, Q5 and Q8 (see Section 5.5) which have been highlighted in red above.

